# Spatial transcriptomics and machine learning define exhaustion-like bone marrow T-cell islands associated with myeloma progression and clinical risk

**DOI:** 10.64898/2026.06.30.26356926

**Authors:** Xinye Li, Xiaolong Jiang, Qiuyan Dong, Jie Wu, Yangqiu Li, Yikai Zhang, Liye Zhong

**Affiliations:** State Key Laboratory of Bioactive Molecules and Druggability Assessment, Key Laboratory of Regenerative Medicine, Ministry of Education, Institute of Hematology, School of Medicine, Jinan University, Guangzhou 510632, China; Department of Hematology, First Affiliated Hospital of Jinan University, Guangzhou 510632, China; Sixth Affiliated Hospital of Sun Yat-sen University, Guangzhou 510655, China

**Keywords:** Multiple myeloma, Spatial transcriptomics, Machine learning, Bone marrow microenvironment, T-cell islands, T cell exhaustion

## Abstract

**Background:** Multiple myeloma (MM) progression is accompanied by remodeling of the bone marrow immune microenvironment. Local interactions among malignant plasma cells, stromal cells, myeloid cells, and immune cells not only support tumor cell survival, expansion, and immune escape, but are also closely associated with disease progression, therapeutic response, and clinical prognosis. Moreover, T cell exhaustion is a common T cells dysfunction in MM and limited efficacy of T cell-targeting therapies. However, the *in situ* organization and clinical significance of exhausted T cells in MM patients bone marrow remain insufficiently understood.

**Methods:** In this study, we analyzed bone marrow Xenium 5K spatial transcriptomics data from control (Ctrl), monoclonal gammopathy of undetermined significance (MGUS), smoldering myeloma (SM), and MM samples. After canonical multi-sample integration and celltype annotation, we used Gaussian mixture model (GMM)-based spatial partitioning, and multilayer perceptron (MLP) machine learning for systematic characterization the T cell microenvironment in MM bone marrow.

**Results:** Our results showed that exhaustion-like T cells increased during MM progression and formed spatially discrete T cell-enriched regions in the bone marrow, which we defined as exhaustion-like bone marrow T cell islands (eBM-TIs). These niches were mainly characterized by enhanced T cell-plasma cell communication associated with upregulated Galectin signaling. Pseudobulk analysis further showed enhanced IFN-related signaling in eBM-TIs, accompanied by upregulation of CXCR3 ligands such as CXCL9 and CXCL10, suggesting that the IFN-CXCL9/10 axis may contribute to T cell chemotaxis, maintenance of chronic inflammation, and formation of exhaustion-like states. By transferring spatial niche labels to scRNA-seq cohorts with available clinical staging information using MLP, we further found that the proportion of eBM-TI-like T cells was associated with higher disease risk and unfavorable prognostic outcomes.

**Conclusions:** In summary, this study identifies eBM-TIs as a spatial niche in the MM bone marrow. These niches represent an important immune unit linking chronic inflammation, T cell exhaustion, and clinical risk, and may serve as a potential biomarker of MM disease progression.

## 1 Background

Multiple myeloma (MM) is a plasma cell malignancy that arises and continuously evolves within the bone marrow microenvironment (BME). In addition to tumor cell-intrinsic genetic and epigenetic abnormalities, the BME plays a decisive role in supporting MM cell survival, expansion, immune escape, and therapeutic resistance ^[1]^. In recent years, single-cell RNA sequencing (scRNA-seq) studies have further shown that, during the progression from control (Ctrl), monoclonal gammopathy of undetermined significance (MGUS) and smoldering myeloma (SM) to active MM, the BME undergoes continuous remodeling, particularly within T/NK cell populations ^[2]^. Patient-specific T cell characteristics, including abundance, cytotoxicity, and exhaustion levels, are emerging as critical determinants of clinical efficacy and drug resistance, particularly with the clinical introduction of B cell maturation antigen (BCMA)- and G protein-coupled receptor class C group 5 member D (GPRC5D)-targeted antibody therapies that engage these cells ^[3-8]^.

It is worth notice that the bone marrow is not a homogeneous tissue, and local cellular composition and signaling outputs are highly spatially dependent ^[9]^. Previous studies with multi-region sequencing have confirmed substantial heterogeneity across different bone marrow sites and local lesions within the same patient ^[10]^. Recent studies based on bone marrow trephine biopsies have also shown that tumor cells, CD8^+^ T cells, and regulatory T cells form mosaic-like local tissue structures ^[11]^. Despite previous bulk and scRNA-seq analysis of bone marrow aspirates that addressed issues related to cellular composition and functional states, such approaches inevitably disrupted the native tissue architecture and are therefore limited in answering questions such as where tumor cells and immune cells encounter each other *in situ*, in which neighborhoods T cell exhaustion occurs, and how local immunosuppressive barriers are formed.

Spatial transcriptomics provides a direct approach to address these questions ^[12]^. Spatial studies of extramedullary myeloma lesions have revealed that functional and exhausted T cells can be spatially segregated and accompanied by marked subclonal heterogeneity ^[13]^. Nevertheless, current imaging-based studies of myeloma bone marrow remain largely focused on methodological establishment and feasibility validation, leaving room for more systematic analysis of the T cell microenvironment. In the study by Yip et al., the authors applied a cell-filtering threshold that removed cells with *<60 transcripts* or *<50 detected genes*, performed unsupervised clustering within each individual sample, and then conducted integrated analysis on the extracted cell populations, leading to several important findings ^[14]^. This workflow is valuable for establishing an analytical framework. However, it remains limited for questions requiring cross-sample comparison and characterization of continuous state transitions. Meanwhile, recent methodological studies in spatial transcriptomics have suggested that multi-sample integration and batch correction can help recover shared spatial structures across samples and improve the reliability of spatial gene-pattern detection and cellular neighborhood comparisons ^[15, 16]^.

Based on these considerations, this study reanalyzed the bone marrow T cell microenvironment across the progression from Ctrl, MGUS, and SM to MM while preserving the original spatial information as much as possible. We first optimized the workflow in terms of cell inclusion quality control, within-group multi-sample integration, and spatial quantitative analysis. We then focused on characterizing the spatial composition, functional states, neighborhood organization, and relationship of T cells with malignant plasma cells (MPCs), aiming to provide a more systematic understanding of immune microenvironment remodeling in MM bone marrow and its potential clinical significance.

## 2 Methods

### 2.1 Preprocessing and quality control of spatial transcriptomics datasets

The data used in this study were downloaded from the Gene Expression Omnibus (GEO) database under accession number GSE299207. According to the original study ^[14]^, raw data processing was performed using Xenium Explorer (v3.0, 10X Genomics). Xenium outputs was first loaded into SpatialData zarr format and quality control was subsequently performed on each sample using Scanpy v1.12.0. For this dataset, we started with utmost preservation of spatial architecture and integrity. Following official 10X Genomics analysis guide (Supplementary Files 1-2), cells with <*20 total transcripts* or <*10 detected genes* were considered as low-quality and therefore removed. The procedures for data merging, data import, and quality-control logic are described in Supplementary Methods S1.

### 2.2 Within-group multi-sample integration and cell annotation

Hoping to retain potential differences in BME composition and disease-associated transcriptional features among Ctrl, MGUS, SM, and MM clinical stages, instead of performing unified integration across all samples from different clinical groups, only samples within the same disease stage was integrated. For each clinical group, highly variable genes were first selected using the Seurat v3 method, followed by data normalization, log-transformation, and principal component analysis. Harmony ^[17]^ was then applied with sample as the batch variable for within-group batch correction, after which uniform manifold approximation and projection (UMAP) embedding and Leiden clustering were performed.

Based on the clustering results, we performed preliminary annotation of each cluster by integrating differentially expressed genes and predefined biomarker genes. Clusters with clear biomarker features were directly assigned to the corresponding cell-type labels. For clusters with insufficient expression information or ambiguous annotation, robust cell type decomposition (RCTD) ^[18]^ and building aggregates with a neighborhood kernel and spatial yardstick (BANKSY) ^[19]^ spatial clustering results were further incorporated for integrated interpretation. Detailed procedures are provided in Supplementary Methods S2.

### 2.3 Advanced spatial analyses and functional-state assessment

Based on the final cell annotations, we performed further spatial analyses. First, using cell centroid coordinates, a fixed-radius neighborhood framework was calculated within each independent sample to evaluate local cellular composition and enrichment relative to the sample-level background, thereby characterizing the neighborhood relationships between T cells and other cells in the BME. Next, local T-cell and plasma-cell densities and their within-sample normalized values were used to construct a two-dimensional colocalization representation. A Gaussian mixture model (GMM) was then applied for unsupervised partitioning of local spatial states ^[20]^, defining T-dominant, plasma-dominant, and mixed spatial regions. Based on these spatial regions, the plasma B-cell malignancy (PBM) score ^[21]^ was mapped to different spatial compartments to compare local differences in plasma cell malignancy. Meanwhile, AUCell scoring was performed for T cells to assess cytotoxicity- and exhaustion-related functional states. We also performed spatial cell-cell communication analysis ^[22]^, pseudobulk-based differential gene expression analysis ^[23]^, and Multilayer perceptron (MLP)-based spatial information mapping of scRNA-seq data ^[24]^. Detailed procedures are described in Supplementary Methods S3.

### 2.4 Statistical analysis

Statistical analyses were performed, including t-test, Wilcoxon rank-sum test, and Kruskal-Wallis test. All significance levels were indicated by p-values, with p < 0.05 considered statistically significant (**** p < 0.0001, *** p < 0.001, ** p < 0.01, * p < 0.05, n.s. not significant).

## 3 Results

### 3.1 T cell-enriched niches emerge in the bone marrow during MM progression

To systematically characterize T cell changes in the BME during MM progression, we first performed spatial single-cell annotation of Xenium bone marrow sections from Ctrl, MGUS, SM, and MM samples. Through multi-sample integration within each clinical group, major bone marrow cell populations were consistently identified across different disease stages (Figure 1, Supplementary Figures 1-3). At the sample level, T cell abundance generally increased with plasma cell disease progression. The proportion of T cells was relatively low in Ctrl samples, whereas SM and MM samples showed higher overall T cell proportions. Several MM samples exhibited marked T cell expansion, with some showing an increase of more than 80% compared with the mean Ctrl level (Figure 1B-E). Further analysis of T cell composition revealed that the CD4^+^/CD8^+^ T cell ratio reached a relative peak at the SM stage and then markedly decreased in active MM, indicating that T cell expansion during progression to MM was accompanied by a gradual shift toward CD8^+^ T cell predominance (Supplementary Figure 4).

**Figure 1.**
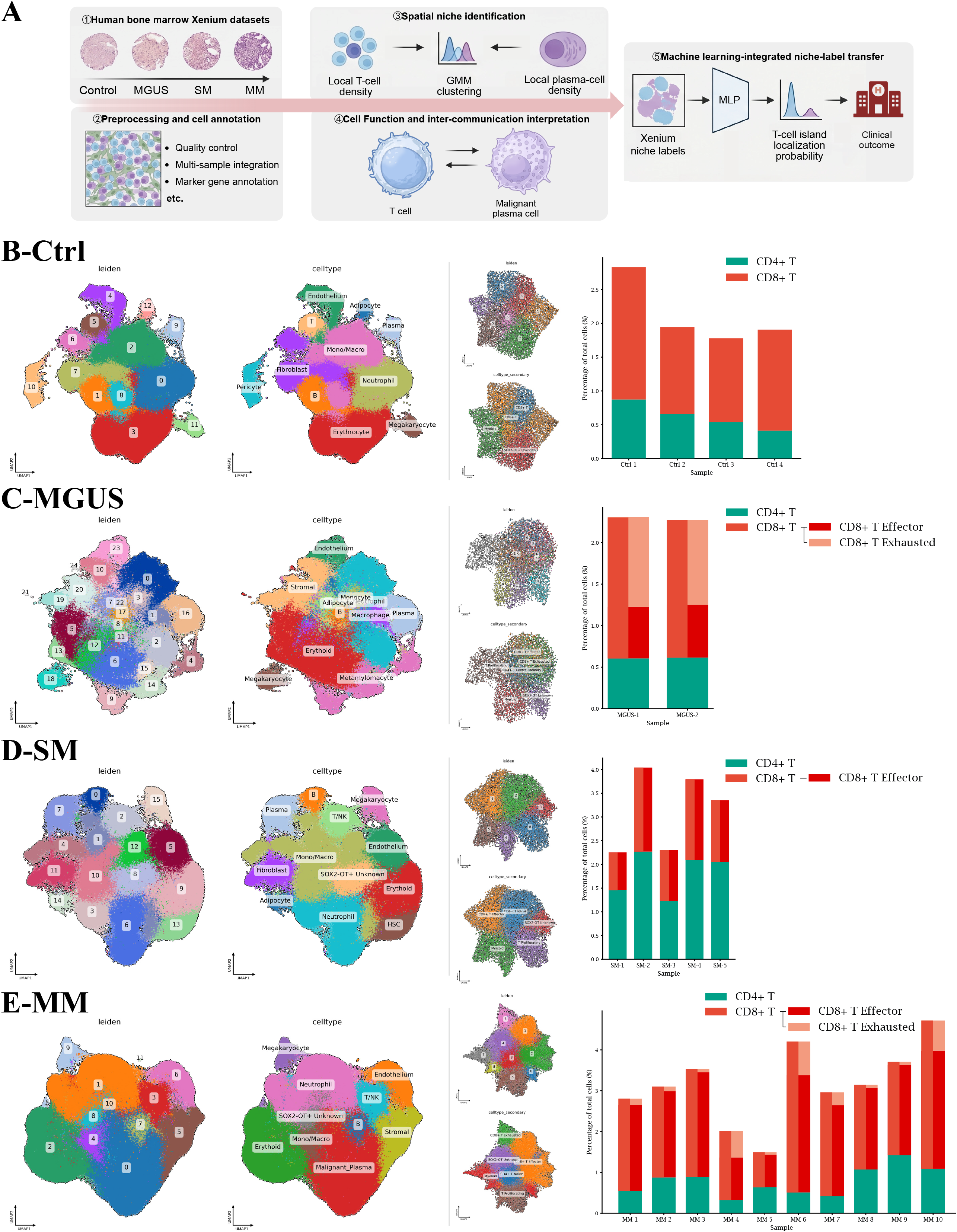
Spatial transcriptomics analysis workflow, bone marrow cell reannotation, and changes in T cell composition (A) Schematic overview of the analytical workflow used in this study. (B-E) UMAP clustering, major cell type annotation, T cell subpopulation annotation, and T cell subpopulation proportions across samples in Ctrl, MGUS, SM, and MM groups, shown in columns 1-4, respectively.

We observed that T cells and plasma cells were unevenly distributed within the tissue. In these regions, T cell-enriched niches could be adjacent to, partially overlapping with, or spatially separated from plasma cell-enriched niches (Figure 2A, Supplementary Figures 5-8). To define these local immune niches, we applied GMM-based unsupervised clustering. This approach consistently identified region that was dominated by T cell, dominated by plasma cell, or lacking clear dominance of either cell type in most samples. After interpretive annotation, these regions were defined as T_Dominant, Plasma_Dominant, and Mixed spatial niches, respectively. In some samples, regions with concomitantly high T cell and plasma cell densities were further identified as Engaging (In SM and MM samples) or Infiltrating (In Ctrl and MGUS samples) regions (Supplementary Figures 9-12).

**Figure 2.**
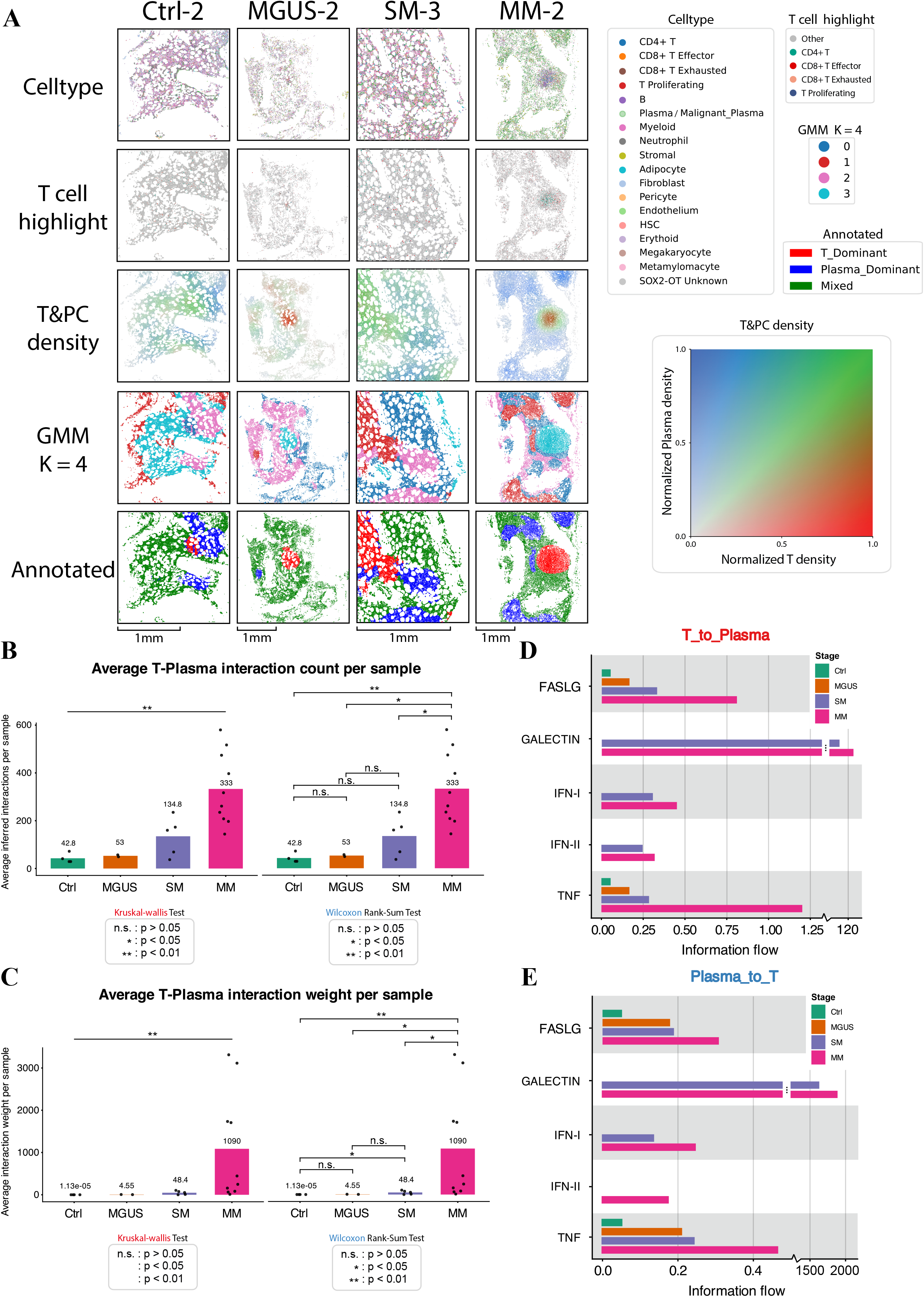
GMM identifies T cell-enriched niches in MM bone marrow and their functional features (A) Spatial maps of representative Ctrl, MGUS, SM, and MM samples. From top to bottom, the panels show the spatial distribution of major cell types, highlighted T cells, local T cell and plasma cell density, GMM-based unsupervised spatial partitioning, and annotated spatial niches. (B-C) Comparison of the average number of ligand-receptor interactions and average communication weights between T cells and plasma cells across Ctrl, MGUS, SM, and MM stages. (D-E) Information-flow analysis in the T_to_Plasma and Plasma_to_T directions, comparing the overall contribution of different immunoregulatory pathways across disease stages.

Among these niches, T_Dominant regions exhibited spatially discrete structures with marked local T cell enrichment, suggesting that relatively independent T cell-enriched niches emerge in the MM bone marrow. Consistently, ligand-receptor interaction analysis showed that both the average number and strength of T cell-plasma cell communications increased with disease progression (Figure 2B-C). Directional communication analysis further showed that information flow in both T-to-Plasma and Plasma-to-T directions changed during disease progression. Interferon (IFN)-related signaling from T cells to plasma cells was enhanced, while GALECTIN-related signaling from plasma cells to T cells was increased, suggesting that local T cells retained inflammatory activation features, whereas MPCs may reciprocally influence T cell functional states through GALECTIN-related pathways (Figure 2D-E). At pathway level, enhanced T cell-plasma cell communication was mainly concentrated in immune regulatory pathways including GALECTIN, IFN-I, IFN-II, tumor necrosis factor (TNF), and FAS ligand (FASLG), among which the GALECTIN pathway was particularly prominent in MM (Figure 3A-C).

**Figure 3.**
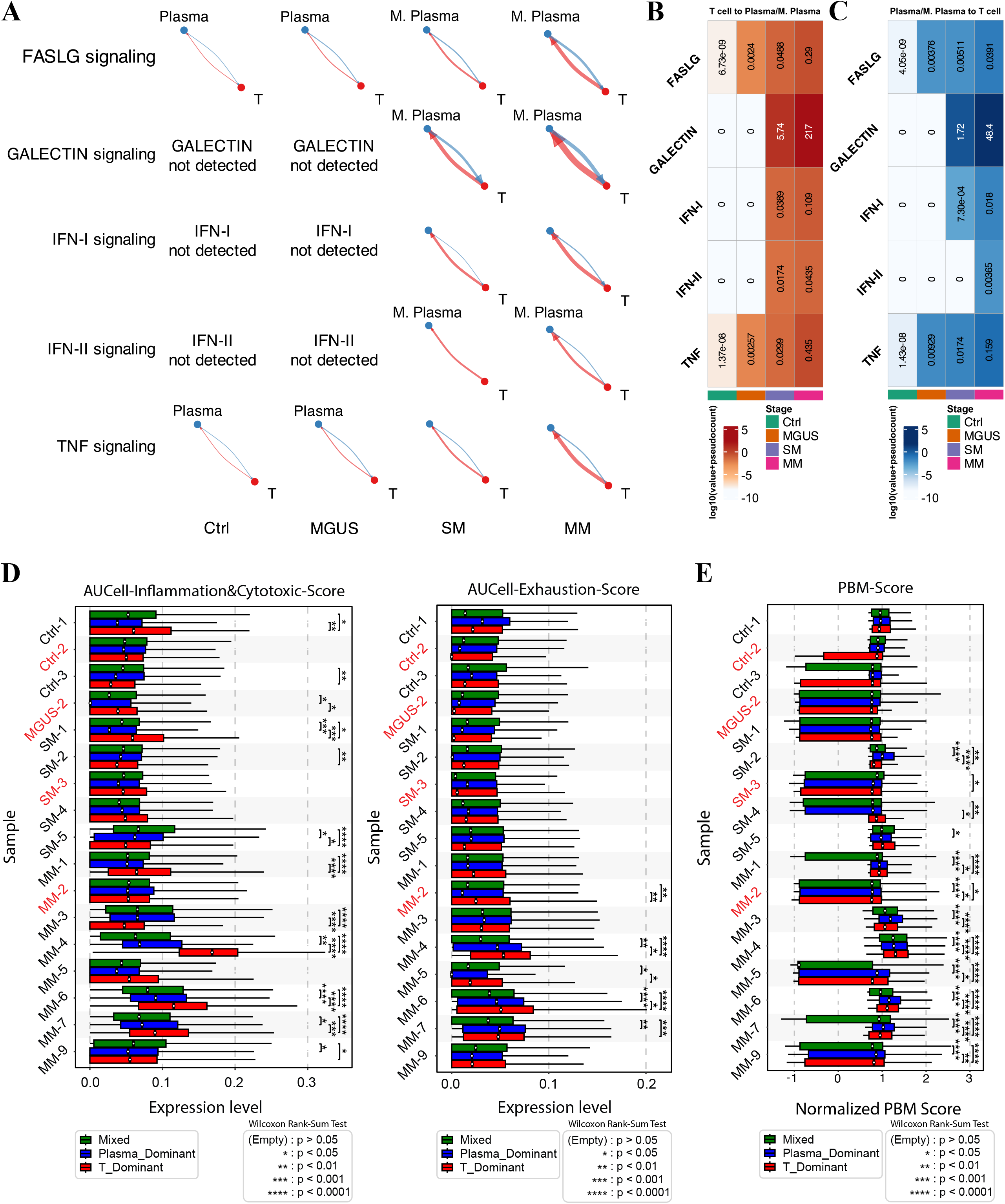
T cell-plasma cell communication is enhanced during MM progression and is characterized by prominent Galectin-related signaling (A) Network diagrams of T cell-plasma cell communication mediated by FASLG, GALECTIN, IFN-I, IFN-II, and TNF pathways across different disease stages. (B-C) Pathway-level communication heatmaps for T cell-to-plasma cell/MPC signaling and plasma cell/MPC-to-T cell signaling. (D) Comparison of AUCell inflammation/cytotoxicity scores and exhaustion scores of T cells across different samples and GMM-defined spatial niches. (E) Comparison of PBM scores in plasma cell/MPC lineages across different samples and GMM-defined spatial niches.

These findings indicate that the T_Dominant niche represents a T cell-enriched spatial niche with distinct communication features in the MM bone marrow.

### 3.2 T_Dominant niches can be defined as exhaustion-like bone marrow T-cell islands

After identifying T_Dominant spatial niches in the MM bone marrow, we further evaluated the functional states of T cells within these T cell-enriched regions. AUCell functional scoring showed that T cells within T_Dominant regions exhibited higher exhaustion-like features, while some samples retained certain inflammatory and cytotoxic signals. These findings suggested that these T cells may exist in an inflammation-associated exhaustion-like state under chronic antigen stimulation (Figure 3D, Supplementary Figure 14). PBM score revealed differences in plasma cell malignancy across GMM-defined spatial regions. Plasma cells located in Plasma_Dominant or T/plasma cell co-enriched regions showed higher PBM-related perturbation (Figure 3E). These results suggested that T cells may be continuously recruited and activated within local plasma cell-associated niches and gradually acquire exhaustion-like phenotypes after prolonged exposure to MPCs and their immunoregulatory signals.

Based on three core features of T_Dominant regions, namely spatially discrete local T cell enrichment, higher T cell exhaustion scores, and enhanced T cell-plasma cell immunoregulatory communication during disease progression, we defined these regions as exhaustion-like bone marrow T-cell islands (eBM-TIs). This term specifically refers to T cell-enriched niches in the MM BME characterized by spatial boundaries, functional bias, and exhaustion-associated communication features.

As a complementary validation, NMF-based whole-celltype neighborhood composition analysis also identified T cell-enriched, plasma cell-enriched, or mixed T/plasma cell neighborhood components across multiple samples, confirming existence of eBM-TIs rather than algorithm bias (Supplementary Figure 13). We therefore proposed that eBM-TIs formed during MM progression may fail to translate into effective anti-myeloma immune responses and instead represent spatial immune niches associated with chronic inflammation and T cell exhaustion.

### 3.3 eBM-TIs are accompanied by enhanced T cell-plasma cell communication centered on Galectin and IFNG-CXCL9/10

We next focused on IFN-associated chemokine signaling. Pseudobulk differential expression analysis showed enhanced IFN-related signals in eBM-TIs, accompanied by prominent upregulation of C-X-C motif chemokine receptor 3 (CXCR3) ligands including C-X-C motif chemokine ligand 9 (CXCL9) and C-X-C motif chemokine ligand 10 (CXCL10) (Figure 4A). Together with previous studies ^[25-27]^, these findings suggested that the interferon gamma (IFNG)-CXCL9/10 axis may contribute to formation and maintenance of eBM-TIs. Activated or inflammatory T/NK cells may produce IFNG, inducing BME cells or plasma cell-associated niches to express CXCL9/10, thereby recruiting or retaining T cells. Under sustained inflammation and tumor antigen exposure, these T cells may further acquire exhaustion-like phenotypes through GALECTIN-related T cell-plasma cell communication. Thus, the IFNG-CXCL9/10 axis and the GALECTIN pathway may jointly constitute a potential mechanism linking T cell chemotaxis, chronic inflammation, and immune exhaustion.

**Figure 4.**
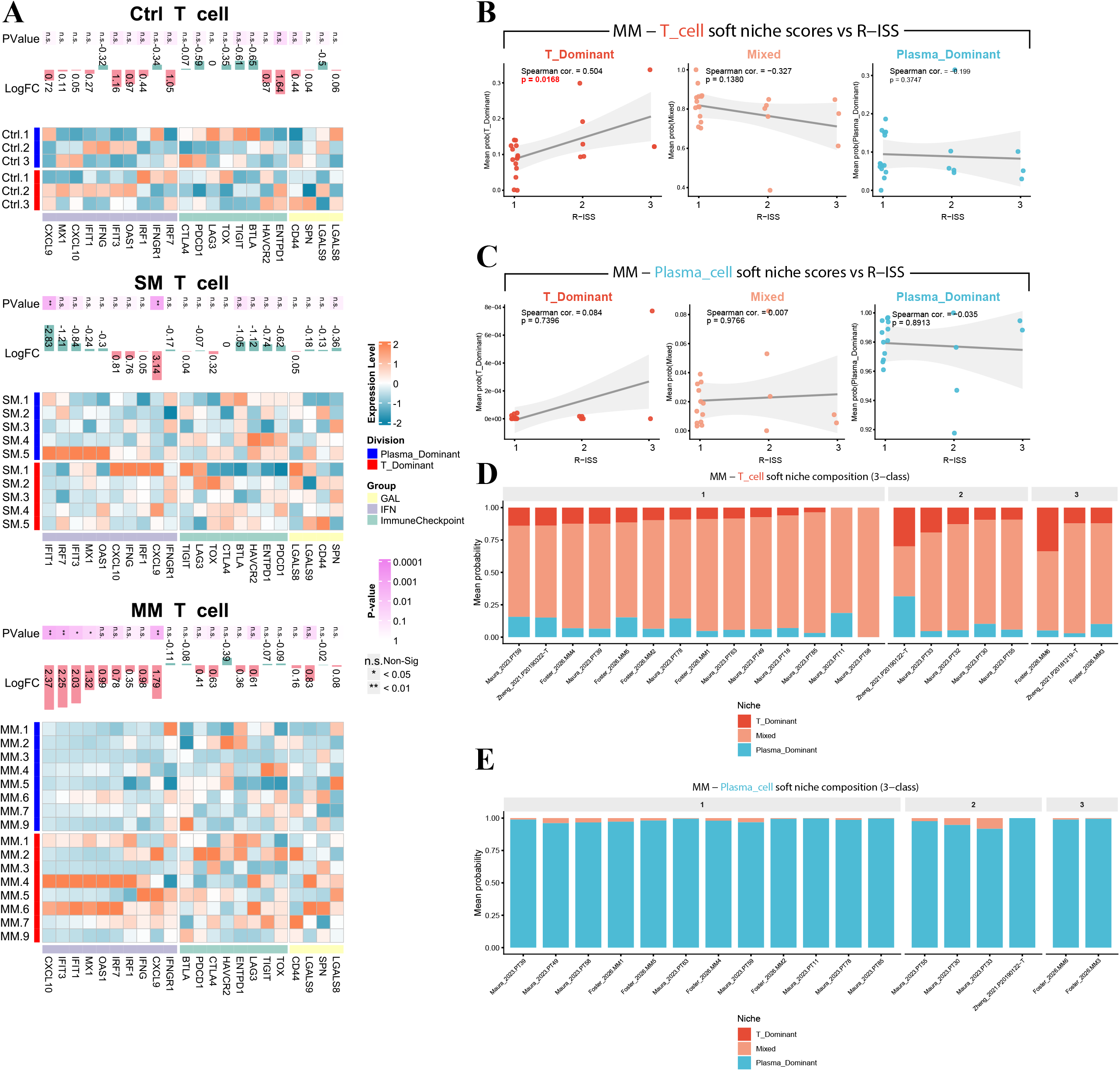
Molecular features of T_Dominant niches and MLP-assisted spatial niche transfer analysis (A) Pseudobulk differential expression analysis of T cells between T_Dominant and Plasma_Dominant spatial niches in Ctrl, SM, and MM samples. (B-C) Correlation analysis between MLP-inferred soft niche scores of T cells and plasma cells and R-ISS stage in an external MM scRNA-seq cohort. (D-E) Proportions of T_Dominant-like, Mixed-like, and Plasma_Dominant-like soft niche probabilities in T cell and plasma cell populations from each MM patient, ordered by R-ISS stage.

So far, our studies have confirmed that T cells localized in eBM-TIs may undergo unique transcriptomic alterations and carry corresponding biological significance. Simultaneous capture of both transcriptomic and geographic information of such T cells made possible by Xenium *in situ* platform, combined with readout similarity with 10X Chromium single-cell platform inspired us to develop an MLP-based machine learning model for prediction of T cell location signature utilizing its transcriptomic features. The results showed that the proportion of eBM-TI-like T cells was positively associated with revised international staging system (R-ISS) stage (Figure 4B-E). In patients with higher R-ISS stages ^[28, 29]^, T cells were more likely to acquire T_Dominant-like spatial niche features, whereas this feature was relatively weaker in lower-risk patients. These findings suggested that presence of eBM-TIs may reflect an immunosuppressive state associated with higher R-ISS stage. The combined effects of the IFNG-CXCL9/10 axis and GALECTIN pathway may represent an important underlying mechanism, providing a basis for future mechanistic studies and therapeutic exploration targeting T cell exhaustion, GALECTIN signaling, and the CXCR3 chemokine axis.

## 4 Discussion

Previous studies have suggested that the BME plays a critical regulatory role during the progression of plasma cell disorders from MGUS to SM and eventually to active MM, with continuous remodeling particularly in T/NK cell populations ^[2]^. However, the specific alterations in BME T cells that may influence disease progression remain incompletely understood, which may involve shifts in T cell lineage composition, spatial distribution, and functional state. Moreover, compared with the patterns of T cell immunosuppression observed in leukemia, it remains unclear whether T cells in the myeloma BME acquire disease-specific alterations in chemotaxis, aggregation, or migratory behavior. Therefore, this study advances the understanding of the MM bone marrow T cell microenvironment from previous cell-proportion level to single-cell spatial resolution. Recent sequencing-based bone marrow studies have suggested that MM plasma cell-infiltrated regions are accompanied by T cell functional gradients, neutrophil extracellular traps (NETs), and interleukin-17 (IL-17)-associated inflammatory changes ^[30, 31]^. Another study of bone marrow breakout lesions further showed that local MM lesions can serve as hotspots of tumor-immune co-evolution ^[32]^. In this context, we define eBM-TIs as a spatial immune unit. Importantly, eBM-TIs should not be directly equated with canonical tertiary lymphoid structures. Before sufficient evidence is obtained for follicular dendritic cells, high endothelial venules, and B cell follicle organization, eBM-TIs are more appropriately defined as T cell-enriched spatial niches in the BME.

scRNA-seq studies by Zavidij et al., Schinke et al., Boiarsky et al., and others have shown that immune microenvironment remodeling occurs early during the progression from MGUS to MM ^[2, 33, 34]^. Larger atlas studies have further linked bone marrow immune states to patient outcomes ^[35]^. Shasha et al. reported that typical terminally exhausted T cells are not broadly present in newly diagnosed multiple myeloma (NDMM) ^[36]^. Our findings suggested that exhausted T cells in the BME mainly exist as eBM-TIs, therefore bone marrow aspiration biopsy may be a better way to detect them. The exhaustion-like state identified in eBM-TIs is spatially restricted and does not represent uniform terminal exhaustion across the entire bone marrow T cell compartment. In light of current understanding of T cell exhaustion, including chronic antigen stimulation, thymocyte selection-associated high mobility group box protein (TOX)-dependent epigenetic fixation, T cell factor-1 (TCF1)^+^ precursor exhausted cells, and heterogeneity of terminal exhaustion ^[37-40]^, eBM-TIs in MM may represent an intermediate adaptive state induced by local chronic stimulation. In this state, T cells may still retain traces of inflammatory or cytotoxic activity, but their function is continuously shaped by immunosuppressive communication.

Goodyear et al. reported that neoplastic plasma cells and their associated stromal components can generate an inflammatory chemokine environment within the bone marrow and alter the distribution of T cells across lymphoid compartments ^[25]^. Bolomsky et al. identified CXCL9 as an independent prognostic factor in NDMM ^[26]^, while Giuliani et al. demonstrated that MM cells express CXCR3 and its ligand system, suggesting that this axis may act not only on T cells but also participate in the survival and migration of plasma cells themselves ^[27]^. These findings are consistent with our results (Figure 4, Supplementary Figure 15). Meanwhile, the CXCL9/10-CXCR3 axis has 2-faced functions in cancer. On the one hand, it can promote the recruitment of Th1/CD8^+^ T/NK cells; on the other hand, it may also maintain chronic inflammation, impair NK cell homing to the bone marrow, or promote immune imbalance ^[41-44]^. Therefore, in this study, the upregulation of CXCL9/10 marks an IFN-driven chronic inflammatory chemotactic program, which may represent one of the initiating factors of T cell exhaustion. These also suggests that the IFNG-CXCL9/10 axis may be one potential mechanism for the formation of eBM-TIs in MM patients’ bone marrow.

The Galectin family is recognized as an important node in glyco-immune regulation and can influence immune cell fate through extracellular glycan recognition as well as non-canonical intracellular mechanisms ^[45, 46]^. In MM, the T-cell immunoglobulin and mucin-domain containing-3 (TIM-3)/Galectin-9 (LGALS9) axis has been proposed as a potential therapeutic target. Its function is context-dependent and may involve immune homeostasis, tumor survival, and immunosuppression ^[45, 47]^. It has also been reported to participate in MM progression by negatively regulating CD4^+^ T cells ^[48]^. Combined with the enhanced Plasma-to-T Galectin communication and differential expression trend of the LGALS9 gene (p = 0.057) observed in this study, these findings suggested that MPCs may restrict T cell effector function through Galectin-associated glyco-immune signaling, thereby transforming eBM-TIs into exhaustion-like immune niches. Therefore, the marked upregulation of the Galectin pathway may be one of the reasons that eBM-TIs fail to exert effective anti-myeloma immune activity.

Finally, using MLP, we projected T cells from scRNA-seq samples with available R-ISS staging information onto the spatial niche framework defined in our spatial transcriptomics analysis ^[2, 49-52]^. Previous studies have demonstrated that the efficacy of T cell-redirecting therapies is closely associated with pretreatment T cell composition, the T cell/MM cell ratio, the immunosuppressive microenvironment, and functional decline induced by persistent stimulation ^[3, 5, 6, 30, 53-56]^. In this study, we found that eBM-TI-like features were significantly associated with higher R-ISS stage in corresponding patients. This suggests that bone marrow spatial immune architecture may represent an important risk-related indicator in MM. Such spatial immune features have likely been underappreciated in previous studies due to technical limitations. Our analytical strategy also provides an exploratory framework for linking discoveries from spatial transcriptomics to conventional scRNA-seq clinical cohorts.

This study also has several limitations. The sample size remains limited, and the Xenium 5K panel cannot fully resolve TCR clonality, metabolic states, or glycosylation-related regulation without customized probes designed with paired-sample single-cell data for reference. The role of CXCL9/10 and Galectin axes in the formation of eBM-TIs still needs to be verified by confirmatory experiments. In addition, T cell location label provided by MLP-based prediction algorithm may only serve predictive purposes at present, and its clinical diagnostical value needs to be verified by more clinical data.

## 5 Conclusions

In this study, we reported scattered T_Dominant niche formation in BME during MM disease progression. This T_Dominant niche is characterized by T cell aggregation, IFNG-CXCL9/CXCL10 chemotactic signaling and galectin-centered T cell-MPC interactions, in which chronic inflammatory recruitment is coupled to progressive T cell exhaustion. Therefore, we defined it as eBM-TIs. Moreover, eBM-TIs is associated with high clinical risk in MM patients. Further elucidation of biological characteristics of eBM-TIs in the development of MM may provide laboratory evidence of eBM-TIs as a biomarker for assessing disease progression/prognosis of MM patients, as well as theraputic target to reshape BME and improve outcome of MM patients.

## Data Availability

Xenium 5K data and scRNA-seq data from published studies were acquired and combined. Specifically, data shared through the GEO can be accessed for Yip et al. under the accession GSE299207, Maura et al. GSE161195, and Zheng et al. GSE156728. Data shared through the Sequence Read Archive (SRA) can be accessed for Foster et al. under the accession PRJNA1401834.
All code for this study is FULLY open source, please refer to our GitHub repository at https://github.com/Flynn2059/Code-concerning-MM-Xenium-Data-analysis-paper.

## Abbreviations

BANKSY: Building aggregates with a neighborhood kernel and spatial yardstick
BCMA: B cell maturation antigen
BME: Bone marrow microenvironment
Ctrl: Control
CXCR3: C-X-C motif chemokine receptor 3
CXCL9: C-X-C motif chemokine ligand 9
CXCL10: C-X-C motif chemokine ligand 10
eBM-TIs: exhaustion-like bone marrow T-cell islands
FASLG: FAS ligand
GEO: Gene Expression Omnibus
GMM: Gaussian mixture model
GPRC5D: G protein-coupled receptor class C group 5 member D
IFN: Interferon
IFNG: Interferon gamma
IL-17: Interleukin-17
LGALS9: Galectin-9
MGUS: Monoclonal gammopathy of undetermined significance
MLP: Multilayer perceptron
MM: Multiple myeloma MPCs - Malignant plasma cells
NDMM: Newly diagnosed multiple myeloma
NETs: Neutrophil extracellular traps
PBM: Plasma B-cell malignancy
RCTD: Robust cell type decomposition
R-ISS: Revised international staging system
scRNA-seq: Single-cell RNA sequencing
SM: Smoldering myeloma
SRA: Sequence Read Archive
TCF1: T cell factor-1
TIM-3: T-cell immunoglobulin and mucin-domain containing-3
TOX: Thymocyte selection-associated high mobility group box protein
UMAP: Uniform manifold approximation and projection

## Funding

This study was supported by grants from the Huahsia Foundation (No. JK-2025-910), the Guangdong Natural Medical Research Association (No. GSNMRA-2026-JC-AL-001).

## Acknowledgments

We thank Professor Raymond K. H. Yip and his team at the Walter and Eliza Hall Institute of Medical Research (WEHI) in Australia for making this data publicly available in the spirit of open science. We also appreciate the support of all doctors and patients involved in this study.

## Author contributions

Y.Q.L., L.Y.Z., Y.K.Z., X.Y.L., and X.L.J. conceptualized and designed the study.

X.L.J. resolved the vast majority of code logic, visualization, and server resource issues, while X.Y.L. was responsible for result interpretation and further analysis.

X.Y.L., and X.L.J. wrote the original manuscript.

Y.Q.L., L.Y.Z., Y.K.Z., Q.Y.D., and J.W. supervised and edited the original manuscript.

## Conflict of interest

The authors declare no competing interests.

## Data availability statement

Xenium 5K data and scRNA-seq data from published studies were acquired and combined. Specifically, data shared through the GEO can be accessed for Yip et al. under the accession GSE299207, Maura et al. GSE161195, and Zheng et al. GSE156728. Data shared through the Sequence Read Archive (SRA) can be accessed for Foster et al. under the accession PRJNA1401834.

All code for this study is FULLY open source, please refer to our GitHub repository at https://github.com/Flynn2059/Code-concerning-MM-Xenium-Data-analysis-paper.

---

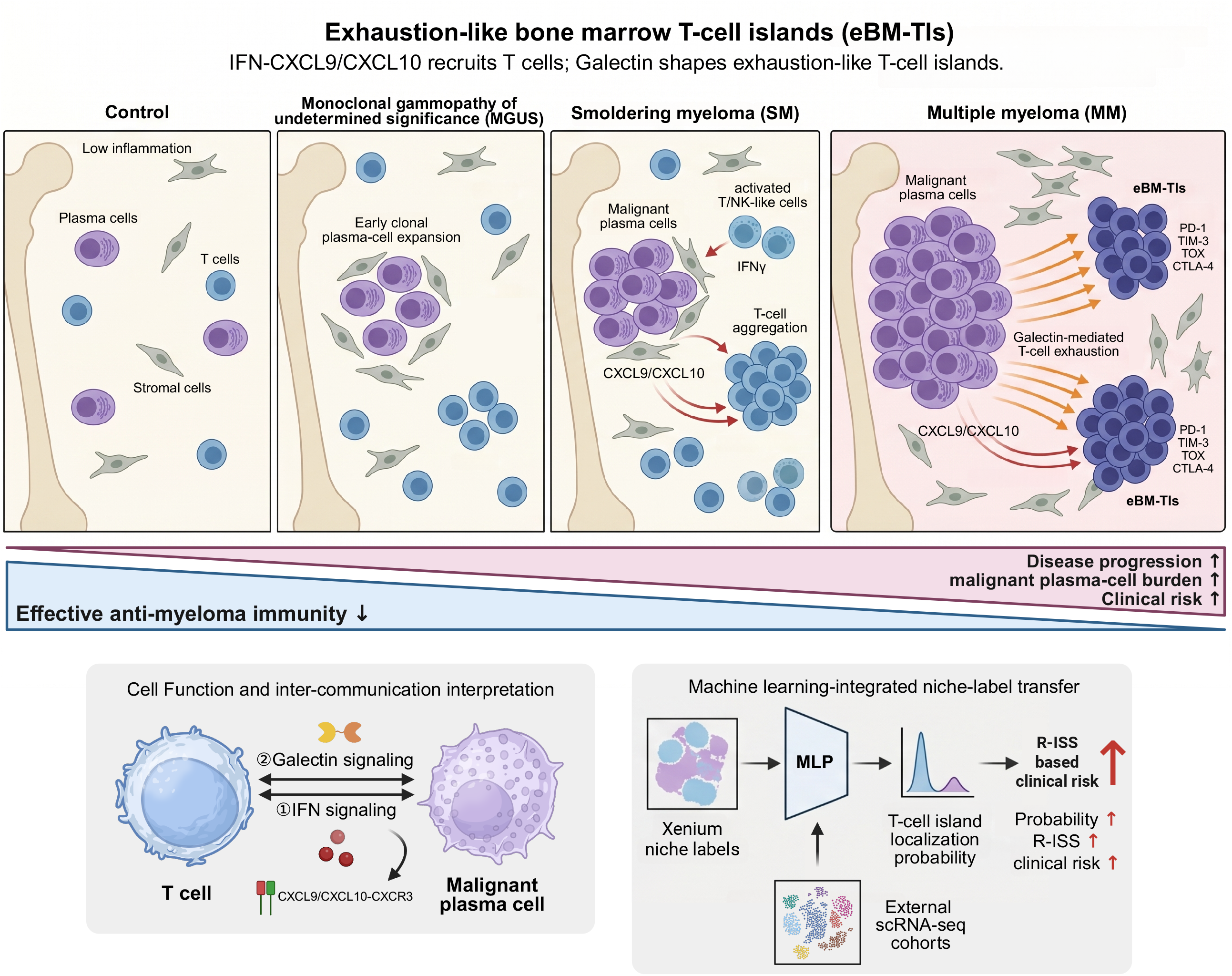

## Notes

### Competing Interest Statement

The authors have declared no competing interest.

### Author Declarations

GSE299207 GSE161195 GSE156728 PRJNA1401834

## References

[1] Wang L, Shi M, Sung AY, Yin CC, Bai Y, Chen M. Role of the bone marrow microenvironment in multiple myeloma: Impact of niches on drug resistance mechanisms.Semin Diagn Pathol. 2025;42(4):150916.

[2] Zavidij O, Haradhvala NJ, Mouhieddine TH, Sklavenitis-Pistofidis R, Cai S, Reidy M, et al. Single-cell RNA sequencing reveals compromised immune microenvironment in precursor stages of multiple myeloma. Nat Cancer. 2020;1(5):493–506.

[3] Verkleij CPM, O’Neill CA, Broekmans MEC, Frerichs KA, Bruins WSC, Duetz C, et al. T-Cell Characteristics Impact Response and Resistance to T-Cell-Redirecting Bispecific Antibodies in Multiple Myeloma. Clin Cancer Res. 2024;30(14):3006–22.

[4] Lin P, Acharya S, Reyes-Silva F, Basar R, Uprety N, Moreno Rueda LY, et al. CD70-Targeting CAR NK Cells Overcome BCMA Downregulation and Improve Survival in High-risk Multiple Myeloma Models. Blood Cancer Discov. 2026;7(2):234–49.

[5] Lee H, Durante M, Skerget S, Vishwamitra D, Benaoudia S, Ahn S, et al. Impact of soluble BCMA and non-T-cell factors on refractoriness to BCMA-targeting T-cell engagers in multiple myeloma. Blood. 2024;144(25):2637–51.

[6] Tedder B, Bhutani M. Resistance Mechanisms to BCMA Targeting Bispecific Antibodies and CAR T-Cell Therapies in Multiple Myeloma. Cells. 2025;14(14):1077.

[7] Rodriguez-Otero P, van de Donk N, Pillarisetti K, Cornax I, Vishwamitra D, Gray K, et al. GPRC5D as a novel target for the treatment of multiple myeloma: a narrative review. Blood Cancer J. 2024;14(1):24.

[8] Van der Vreken A, Meeus F, Tu C, van den Broecke L, Raimondi V, Vescovini R, et al. B7-H3 nanobody-based CAR T cells control multiple myeloma growth, while dual BCMA/B7-H3 CAR T cells overcome antigen escape. J Hematol Oncol. 2025;18(1):103.

[9] Baccin C, Al-Sabah J, Velten L, Helbling PM, Grunschlager F, Hernandez-Malmierca P, et al. Combined single-cell and spatial transcriptomics reveal the molecular, cellular and spatial bone marrow niche organization. Nat Cell Biol. 2020;22(1):38–48.

[10] Rasche L, Chavan SS, Stephens OW, Patel PH, Tytarenko R, Ashby C, et al. Spatial genomic heterogeneity in multiple myeloma revealed by multi-region sequencing. Nat Commun. 2017;8(1):268.

[11] Hagos YB, Lecat CSY, Patel D, Mikolajczak A, Castillo SP, Lyon EJ, et al. Deep Learning Enables Spatial Mapping of the Mosaic Microenvironment of Myeloma Bone Marrow Trephine Biopsies. Cancer Res. 2024;84(3):493–508.

[12] Cheng X, Peng T, Chu T, Yang Y, Liu J, Gao Q, et al. Application of single-cell and spatial omics in deciphering cellular hallmarks of cancer drug response and resistance. J Hematol Oncol. 2025;18(1):70.

[13] John M, Helal M, Duell J, Mattavelli G, Stanojkovska E, Afrin N, et al. Spatial transcriptomics reveals profound subclonal heterogeneity and T-cell dysfunction in extramedullary myeloma. Blood. 2024;144(20):2121–35.

[14] Yip RKH, Er J, Qin L, Nguyen QH, Motyer A, Rimes JS, et al. Profiling the spatial architecture of multiple myeloma in human bone marrow trephine biopsy specimens with spatial transcriptomics. Blood. 2025;146(15):1837–49.

[15] Long Y, Ang KS, Li M, Chong KLK, Sethi R, Zhong C, et al. Spatially informed clustering, integration, and deconvolution of spatial transcriptomics with GraphST. Nat Commun. 2023;14(1):1155.

[16] Millard N, Chen JH, Palshikar MG, Pelka K, Spurrell M, Price C, et al. Batch correcting single-cell spatial transcriptomics count data with Crescendo improves visualization and detection of spatial genepatterns. Genome Biol. 2025;26(1):36.

[17] Korsunsky I, Millard N, Fan J, Slowikowski K, Zhang F, Wei K, et al. Fast, sensitive and accurate integration of single-cell data with Harmony. Nat Methods. 2019;16(12):1289–96.

[18] Cable DM, Murray E, Zou LS, Goeva A, Macosko EZ, Chen F, et al. Robust decomposition of cell type mixtures in spatial transcriptomics. Nat Biotechnol. 2022;40(4):517–26.

[19] Singhal V, Chou N, Lee J, Yue Y, Liu J, Chock WK, et al. BANKSY unifies cell typing and tissue domain segmentation for scalable spatial omics data analysis. Nat Genet. 2024;56(3):431–41.

[20] Pedregosa F, Varoquaux G, Gramfort A, Michel V, Thirion B, Grisel O, et al. Scikit-learn: Machine Learning in Python. J Mach Learn Res. 2011;12:2825–30.

[21] Li JR, Arsang-Jang S, Cheng Y, Sun F, D’Souza A, Dhakal B, et al. Enhancing prognostic power in multiple myeloma using a plasma cell signature derived from single-cell RNA sequencing. Blood Cancer J. 2024;14(1):38.

[22] Jin S, Plikus MV, Nie Q. CellChat for systematic analysis of cell-cell communication from single-cell transcriptomics. Nat Protoc. 2025;20(1):180–219.

[23] Li X, Chen Z, Chen J, Zhang X, Zheng J, Liu Z, et al. Impact of G-CSF on Donor TCR Clonal Diversity and T Cell Function During Donor HSC Mobilisation. Cell Prolif. 2026:e70213.

[24] Guo M-H, Liu Z-N, Mu T-J, Hu S-M. Beyond Self-Attention: External Attention Using Two Linear Layers for Visual Tasks. IEEE Trans Pattern Anal Mach Intell. 2023;45(5):5436–47.

[25] Goodyear OC, Essex S, Seetharam A, Basu S, Moss P, Pratt G. Neoplastic plasma cells generate an inflammatory environment within bone marrow and markedly alter the distribution of T cells between lymphoid compartments. Oncotarget. 2017;8(18):30383–94.

[26] Bolomsky A, Schreder M, Hubl W, Zojer N, Hilbe W, Ludwig H. Monokine induced by interferon gamma (MIG/CXCL9) is an independent prognostic factor in newly diagnosed myeloma. Leuk Lymphoma. 2016;57(11):2516–25.

[27] Giuliani N, Bonomini S, Romagnani P, Lazzaretti M, Morandi F, Colla S, et al. CXCR3 and its binding chemokines in myeloma cells: expression of isoforms and potential relationships with myeloma cell proliferation and survival. Haematologica. 2006;91(11):1489–97.

[28] Rajkumar SV, Dimopoulos MA, Palumbo A, Blade J, Merlini G, Mateos MV, et al. International Myeloma Working Group updated criteria for the diagnosis of multiple myeloma. Lancet Oncol. 2014;15(12):E538–E48.

[29] Wu SQ, Li XF, Qiu ZJ, Zhu ZJ, Chen XL, Chen P, et al. Comparison of tandem and single autologous stem cell transplantation in multiple myeloma: a retrospective propensity score-matching study. Blood Sci. 2025;7(2):e00235.

[30] Muinos-Lopez E, Lopez-Perez AR, Sudupe L, Vilas-Zornoza A, Sarvide S, Ripalda-Cemborain P, et al. Characterization of the bone marrow architecture of multiple myeloma using spatial transcriptomics. Commun Biol. 2025;8(1):1620.

[31] Zhang S, Guo R, Liu Y, Wu Z, Song Y. Basic and applied research progress of TRAIL in hematologic malignancies. Blood Sci. 2025;7(2):e00221.

[32] Lutz R, Poos AM, Sole-Boldo L, John L, Wagner J, Prokoph N, et al. Bone marrow breakout lesions act as key sites for tumor-immune cell diversification in multiple myeloma. Sci Immunol. 2025;10(104):eadp6667.

[33] Schinke C, Poos AM, Bauer M, John L, Johnson S, Deshpande S, et al. Characterizing the role of the immune microenvironment in multiple myeloma progression at a single-cell level. Blood Adv. 2022;6(22):5873–83.

[34] Boiarsky R, Haradhvala NJ, Alberge JB, Sklavenitis-Pistofidis R, Mouhieddine TH, Zavidij O, et al. Single cell characterization of myeloma and its precursor conditions reveals transcriptional signatures of early tumorigenesis. Nat Commun. 2022;13(1):7040.

[35] Pilcher WC, Yao L, Gonzalez-Kozlova E, Pita-Juarez Y, Karagkouni D, Acharya CR, et al. A single-cell atlas characterizes dysregulation of the bone marrow immune microenvironment associated with outcomes in multiple myeloma. Nat Cancer. 2026;7(1):224–46.

[36] Shasha C, Glass DR, Moelhman E, Islas L, Tian Y, Chour T, et al. Hallmarks of T-cell exhaustion and antigen experience are absent in multiple myeloma from diagnosis to maintenance therapy. Blood. 2025;145(26):3113–23.

[37] Wherry EJ, Kurachi M. Molecular and cellular insights into T cell exhaustion. Nat Rev Immunol. 2015;15(8):486–99.

[38] Blank CU, Haining WN, Held W, Hogan PG, Kallies A, Lugli E, et al. Defining ‘T cell exhaustion’. Nat Rev Immunol. 2019;19(11):665–74.

[39] Beltra JC, Manne S, Abdel-Hakeem MS, Kurachi M, Giles JR, Chen Z, et al. Developmental Relationships of Four Exhausted CD8(+) T Cell Subsets Reveals Underlying Transcriptional and Epigenetic Landscape Control Mechanisms. Immunity. 2020;52(5):825–41 e8.

[40] Khan O, Giles JR, McDonald S, Manne S, Ngiow SF, Patel KP, et al. TOX transcriptionally and epigenetically programs CD8(+) T cell exhaustion. Nature. 2019;571(7764):211–8.

[41] Bonanni V, Antonangeli F, Santoni A, Bernardini G. Targeting of CXCR3 improves anti-myeloma efficacy of adoptively transferred activated natural killer cells. J Immunother Cancer. 2019;7(1):290.

[42] Tokunaga R, Zhang W, Naseem M, Puccini A, Berger MD, Soni S, et al. CXCL9, CXCL10, CXCL11/CXCR3 axis for immune activation -A target for novel cancer therapy. Cancer Treat Rev. 2018;63:40–7.

[43] Wang X, Zhang Y, Wang S, Ni H, Zhao P, Chen G, et al. The role of CXCR3 and its ligands in cancer. Front Oncol. 2022;12:1022688.

[44] Korbecki J, Barczak K, Bosiacka B, Surowka A, Duchnik E, Skarbinski M, et al. The Importance of Chemokines Activating CXCR1, CXCR2 and CXCR3 in Tumorigenesis as Potential Therapeutic Targets in Monoclonal Gammopathy of Undetermined Significance and Multiple Myeloma. Cancers (Basel). 2025;17(17):2888.

[45] Shil RK, Mohammed NBB, Dimitroff CJ. Galectin-9 -ligand axis: an emerging therapeutic target for multiple myeloma. Front Immunol. 2024;15:1469794.

[46] Mariño KV, Cagnoni AJ, Croci DO, Rabinovich GA. Targeting galectin-driven regulatory circuits in cancer and fibrosis. Nat Rev Drug Discov. 2023;22(4):295–316.

[47] Okoye I, Xu L, Motamedi M, Parashar P, Walker JW, Elahi S. Galectin-9 expression defines exhausted T cells and impaired cytotoxic NK cells in patients with virus-associated solid tumors. J Immunother Cancer. 2020;8(2):e001849.

[48] Zhang R, Chen S, Luo T, Guo S, Qu J. Activated Tim-3/Galectin-9 participated in the development of multiple myeloma by negatively regulating CD4 T cells. Hematology. 2024;29(1):2288481.

[49] Foster KA, Rees E, Ainley L, Laidler A, Boyle EM, Lee L, et al. Tumour-intrinsic features shape T cell differentiation through precursor to symptomatic multiple myeloma. Nat Commun. 2026;17(1):2400.

[50] Maura F, Boyle EM, Coffey D, Maclachlan K, Gagler D, Diamond B, et al. Genomic and immune signatures predict clinical outcome in newly diagnosed multiple myeloma treated with immunotherapy regimens. Nat Cancer. 2023;4(12):1660–74.

[51] Zheng L, Qin S, Si W, Wang A, Xing B, Gao R, et al. Pan-cancer single-cell landscape of tumor-infiltrating T cells. Science. 2021;374(6574):abe6474.

[52] Cohen YC, Zada M, Wang SY, Bornstein C, David E, Moshe A, et al. Identification of resistance pathways and therapeutic targets in relapsed multiple myeloma patients through single-cell sequencing. Nat Med. 2021;27(3):491–503.

[53] Zylka K, Kubicki T, Gil L, Dytfeld D. T-cell exhaustion in multiple myeloma. Expert Rev Hematol. 2024;17(7):295–312.

[54] Chung DJ, Pronschinske KB, Shyer JA, Sharma S, Leung S, Curran SA, et al. T-cell Exhaustion in Multiple Myeloma Relapse after Autotransplant: Optimal Timing of Immunotherapy. Cancer Immunol Res. 2016;4(1):61–71.

[55] Philipp N, Kazerani M, Nicholls A, Vick B, Wulf J, Straub T, et al. T-cell exhaustion induced by continuous bispecific molecule exposure is ameliorated by treatment-free intervals. Blood. 2022;140(10):1104–18.

[56] Friedrich MJ, Neri P, Kehl N, Michel J, Steiger S, Kilian M, et al. The pre-existing T cell landscape determines the response to bispecific T cell engagers in multiple myeloma patients. Cancer Cell. 2023;41(4):711–25 e6.

