## Supplementary File 1 for "Spatial transcriptomics and machine learning define exhaustion-like bone marrow T-cell islands associated with myeloma progression and clinical risk"

**Supplementary Method for MM Xenium Paper**

**S1** Data import and quality-control (QC) strategy

**S1.1** Data import strategy

The data used in this study were downloaded from the Gene Expression Omnibus (GEO) database under accession number **GSE299207**. The workflow was as follows:

1. The *cell_feature_matrix.h5* was read to obtain the cell × gene expression matrix. To avoid indexing conflicts caused by duplicated gene names, gene names were made unique before downstream analysis.

2. The *cells.csv.gz* was read to extract cell-level annotation information and spatial coordinate information. We used *cell_id* as the unified index and strictly aligned it with the cell order in the expression matrix. The cell centroid coordinates, *x_centroid* and *y_centroid*, were stored in the spatial coordinate matrix for subsequent spatial localization, visualization, and neighborhood analysis, while the remaining cell-level information was retained as observation metadata.

3. The *morphology.ome.tif* was read and incorporated into the unified data structure as a 2-dimensional image object, enabling subsequent integration of tissue morphology with gene expression information.

4. The *transcripts.parquet* was read, and the spatial coordinate fields were standardized to *x* and *y*. For each transcript, the corresponding cell identifier and gene name were retained, and the transcript records were constructed as a point-set object to support subsequent transcript-level QC.

5. The *cell_boundaries.csv.gz* was read, and cell contours were reconstructed by grouping vertices according to *cell_id* and *label_id*. The vertex coordinates of each group were organized as polygon objects. For cells containing multiple boundary fragments, these fragments were further merged into complex polygon objects. After construction, only boundary information that could be matched to the cell indices in the expression matrix was retained, ensuring strict consistency between spatial geometric information and expression data.

After all components were imported, the expression matrix, cell annotations, spatial coordinates, transcript point locations, cell boundaries, and morphology image were jointly encapsulated into a ***SpatialData*** object. This data structure served as the standard input and output format (StdIO) for all subsequent analyses in this study, including QC, clustering, annotation, and downstream spatial analyses.

**S1.2** Data QC strategy

We noted that, when applying QC strategy used in the original study (excluding cells with “*<60 total transcript counts*” or “*<50 detected genes*”), the proportion of cells removed in some samples could be as much as 70%. Although stringent filtering criteria helps remove low-quality cells, excessive QC in spatial transcriptomics data might disrupt the continuity of cellular spatial distributions, generating numerous “spatial void” within tissue regions. This may introduce systematic biases into downstream analyses, especially cell density estimation, spatial neighborhood identification, and local microenvironment characterization. Based on such consideration, we sought to balance between cell quality and preservation of tissue integrity.

Following the recommended standards in the official 10X Genomics Xenium analysis guide*, we adopted the filtering thresholds described in the main text to remove low-quality cells, that is exclusion of cells with “*<20 total transcript counts*” or “*<10 detected genes*”. The maximum proportion of cells removed across samples was approximately 35%, and approximately 1.47M cells were retained in total. Compared with the filtering strategy used in the original study, this approach better preserved the continuity of tissue spatial architecture while still controlling for low-quality cells, thereby providing better data foundation for subsequent spatial analyses.

With aforementioned QC criteria, the overall dataset did not show extensive enrichment of low-quality cells or widespread ambient RNA contamination. However, during cell clustering at different hierarchical levels, we consistently identified a SOX2 Overlapping Transcript (SOX2-OT)^high^ cluster that could not be clearly assigned to a defined cell type. Based on its expression profile and subsequent validation, we considered this cluster to mainly reflect ambient RNA contamination. The detailed criteria for this interpretation and the corresponding handling strategy are described in **S4.1**.

*10X Genomics official guideline, latest version: https://github.com/10XGenomics/analysis_guides/blob/main/Xenium_5k_data_analysis_journey_python.ipynb

The version referenced in this study was the version dated March 16, 2026, which is provided as ***Supplementary File 2***.

**S2** Data integration and cell annotation

**S2.1** Multi-sample integration within clinical groups

Considering the substantial differences in bone marrow microenvironment (BME) composition and disease-associated transcriptional features among the control (Ctrl), monoclonal gammopathy of undetermined significance (MGUS), smoldering myeloma (SM), and multiple myeloma (MM) clinical stages, integration of data across disease states may confound stage-related biological differences with technical variation. Therefore, in this study, multi-sample integration was performed separately within sample of each clinical states. Specifically, joint analyses were conducted independently within the Ctrl, MGUS, SM, and MM groups, without applying a global batch correction across different clinical stages.

Within each clinical groups, we applied a consistent expression-matrix integration workflow. Highly variable genes (HVGs) were selected using the *Seurat v3* method, followed by log-normalization and principal component analysis (***PCA***). ***Harmony*** algorithm was further introduced for batch correction with *sample* as the batch variable. Uniform manifold approximation and projection (***UMAP***) embedding was then computed, followed by ***Leiden*** clustering. The clustering resolution for each clinical group was adjusted according to within-group heterogeneity and annotation interpretability, in order to obtain results that balanced clustering stability with biological interpretability.

Based on clustering results, we used *sc.tl.rank_genes_groups* to identify differentially expressed genes (DEGs) for each cluster and performed initial annotation using a predefined biomarkers, which covered the major cell types in the BME, including hematopoietic stem cells, erythroid cells, megakaryocytes, monocytes/macrophages, T/NK cells, B cells, plasma cells, neutrophils, endothelial cells, and multiple stromal cell populations. Clusters with clear biomarker features were directly assigned to the corresponding cell-type labels. For clusters with insufficient expression information or ambiguous annotation, robust cell type decomposition (***RCTD***) and building aggregates with a neighborhood kernel and spatial yardstick (***BANKSY***) results were further incorporated to assist the final interpretation.

**S2.2** ***RCTD***-assisted annotation based on reference single-cell data

We used the ***RCTD*** function from the ***R*** package *spacexr* to perform reference mapping for each Xenium sample separately. The reference single-cell dataset used for ***RCTD*** was obtained from Foster et al., 2026*.

For each spatial transcriptomics sample, a Reference object was constructed using the processed single-cell reference dataset. The *create.RCTD* and *run.RCTD* functions were then called to complete the mapping. This procedure was performed independently for all samples, rather than being run once after within-group integration.

After ***RCTD*** was completed, we extracted the result information from *results_df*, removed cells labeled as *reject*, and retained only cells with valid mapping results together with their *first_type* labels as the final reference-derived labels. The ***RCTD*** results for each sample were then mapped back to the corresponding ***SpatialData*** object to generate the *rctd_inferred_celltype* field. This field served as independent evidence to assist the interpretation of ambiguous ***Leiden*** clusters identified in **S2.1**.

* Foster, et al., 2026 (In-text citation number 47)

Foster KA, Rees E, Ainley L, Laidler A, Boyle EM, Lee L, et al. Tumour-intrinsic features shape T cell differentiation through precursor to symptomatic multiple myeloma. Nat Commun. 2026;17(1):2400.

**S2.3** Handling ambiguous clusters using ***BANKSY***

In several clinical groups, we found that ***Harmony****-****Leiden*** clustering, which was based solely on the expression matrix, produced a small number of clusters with atypical biomarker expression, a limited number of DEGs, or inconsistent cell-type assignment in ***RCTD*** mapping. To further incorporate spatial neighborhood information and improve cluster interpretability, we introduced ***BANKSY*** for multi-sample spatially aware clustering. ***BANKSY*** was used as an important auxiliary approach for reviewing ambiguous clusters, mainly in clinical groups with more challenging annotation, including MGUS, SM, and MM.

***BANKSY*** was performed using the *.adata* object of each sample as input. Cell spatial coordinates were extracted from *adata.obsm["spatial"]* and written into the *x_slide_mm* and *y_slide_mm* fields. To support multi-sample joint spatial clustering, multiple samples within the same clinical group were concatenated. A fixed interval shift along the x-axis was then applied to different samples, so that originally independent tissue sections were mapped into a shared coordinate system while avoiding spatial overlap between samples. The corrected spatial coordinates were subsequently used to construct *coord_xy*, which served as the basis for ***BANKSY*** neighborhood-structure calculation.

During ***BANKSY*** preprocessing, the concatenated object was normalized, log-transformed, and subjected to HVGs selection. The spatial neighborhood graph was then initialized based on the spatial coordinates. The *initialize_banksy* and *generate_banksy_matrix* functions were used to construct the spatially enhanced expression matrix. After obtaining the ***BANKSY*** low-dimensional representation, we further applied ***Harmony*** correction to its’ ***PCA*** results with *sample* as the batch variable to reduce residual batch effects during multi-sample integration. ***UMAP*** embedding was then computed, followed by ***Leiden*** clustering, generating the final *banksy_leiden* cluster labels. The corresponding ***PCA*** coordinates, ***UMAP*** embeddings, and clustering labels were saved and reintegrated into the objects generated in **S2.1**.

The purpose of ***BANKSY*** was to provide additional spatial-organization evidence for ambiguous clusters that could not be fully resolved in **S2.1** and **S2.2**. For clusters that were difficult to define, we jointly examined their ***BANKSY*** cluster assignment, spatial continuity, and consistency with ***RCTD***-inferred labels. If a suspicious cluster could be stably incorporated into a known cell-type neighborhood in ***BANKSY*** and was consistent with biomarker expression or ***RCTD*** results, this evidence was used to strengthen the confidence of its annotation. Conversely, if a cluster remained independent and poorly informative across multiple evidence systems, it was retained as an undefined subpopulation and discussed separately in subsequent analyses.

**S2.4** Integrated cell-annotation strategy

Based on the workflow above, clusters with clear biomarker expression, stable differential-expression features, and consistent evidence from ***RCTD/Banksy*** were directly assigned to the corresponding cell-type labels. Clusters with insufficient biomarker information yet rather clear assignment tendencies in ***RCTD/Banksy*** were annotated after integrating evidence from multiple sources. Cell populations that lacked clear biological identity across multiple methods but formed stable independent clusters were retained as undefined subpopulations.

This hierarchical annotation framework avoided overreliance on a single clustering method or a single reference-mapping result. It also reduced potential mis-annotation caused by expression noise, spatial mixing, or ambient RNA contamination in the complex BME. The final annotation results were written back into the within-group integrated objects for subsequent cell-composition comparison, spatial neighborhood analysis, and other downstream analyses. Special undefined clusters, such as the recurrent SOX2-OT^high^ cluster observed across multiple clinical groups, are discussed in **S4.1** regarding their potential nature and retention strategy.

**S3** Advanced analysis workflow

**S3.1** Neighborhood analysis

To systematically evaluate the spatial organization of T cells and other BME cell types at the local level, we constructed a coordinate-annotation matrix for Ctrl, MGUS, SM, and MM based on the final cell annotations. Specifically, we extracted the 2-dimensional spatial coordinates from *adata.obsm["spatial"]* of the integrated objects for each clinical group and assigned final cell-type annotations to an *annotation* field. For T-cell subtypes, the secondary annotations were further propagated back to the corresponding major T-cell annotation. Considering that the *SOX2-OT Unknown* population is more likely to represent low-information or ambient RNA-derived signals and may introduce noise into local composition estimates, these cells were excluded when constructing the input matrix for neighborhood analysis. The final coordinate-annotation matrix contained *X*, *Y*, *annotation* and was used for all subsequent centroid-based neighborhood computations.

During neighborhood calculation, analysis was performed separately for each individual sample within each clinical group. We used *scipy.spatial.cKDTree* to construct a spatial index based on cell centroids and retrieved neighboring cells within a fixed radius around each cell. Considering that the spatial organization of the BME may vary across scales, we performed sensitivity analyses using three radii: 50, 75, and 100 μm. For each center cell, neighboring cell-type proportions were computed after excluding the center cell itself, defined as the *neighborhood percentage*. In addition, the global proportion of each cell type within the corresponding sample was used as background, and enrichment was quantified as

$$Fold=\frac{neighborhood percentage}{sample percentage}$$

Finally, both neighborhood percentage matrices and enrichment fold-change matrices were generated, along with metadata including *cell_id*, *sample*, *cell type*, and *n_neighbors* (number of neighbors). The neighborhood percentage matrices were further used for downstream non-negative matrix factorization (***NMF***)-based neighborhood pattern identification, group comparisons, and visualization.

**S3.2** Neighborhood pattern recognition

To further reduce high-dimensional neighborhood composition profiles into interpretable local microenvironmental states, we performed neighborhood pattern recognition using ***NMF***-based on th.e fixed-radius neighborhood results. The *neighborhood percentage* matrix at a 50 μm radius was used as the primary input. For each sample within each clinical group, we retained only cells with more than 5 neighboring cells as valid central cells, and applied *log1p(pct × 100)* transformation to neighborhood proportion features to avoid the influence of extreme values.

***NMF*** was performed independently for each sample, decomposing each cell’s neighborhood composition into a low-dimensional matrix of component scores (*W*), while retaining the component–cell-type loading matrix (*H*) for biological interpretation of each neighborhood pattern. ***NMF*** was initialized using *nndsvda*, with *random_state = 42* and *max_iter = 1000*. The number of components (*K*) was selected according to dataset complexity: *K = 3-5* for Ctrl and MGUS, *K = 3-6* for SM, and *K = 3-10* for MM. After obtaining the ***NMF*** *W* matrix, we constructed a k-nearest neighbor graph in the *W* space and performed ***Leiden*** clustering with resolutions of 0.05, 0.1, and 0.2 to identify neighborhood states with similar local cellular compositions. Each ***Leiden*** cluster was assigned a unique identifier incorporating *sample*, *K* value, and *resolution* to avoid label ambiguity across samples. Finally, we aggregated neighborhood labels, cluster sizes, and ***NMF*** reconstruction errors across samples, K values, and resolutions, and used ***NMF*** *H*-loading heatmaps to interpret the dominant cellular composition underlying each neighborhood state.

**S3.3** Colocalization analysis

To more directly characterize the spatial relationship between T cells and plasma cells at the local level, we further quantified local density based on the geometric centroids of cell boundaries and represented the T cell-plasma cell colocalization state as a 2-dimensional continuous variable. Specifically, after obtaining the final cell annotations, we mapped them back to the *cell_boundaries* object of each sample and extracted the *geometric centroid* of each cell as its spatial coordinate. A ***KDTree*** was then used to query neighboring cells within a fixed radius of 200 μm. For each query, we counted the number of T cells and plasma cells within the neighborhood and normalized by the window area (*πr²*) to obtain *local_t_density* and *local_plasma_density*.

To eliminate biases introduced by differences in overall cell density and tissue area across samples, we further performed within-sample normalization of T cell and plasma cell local densities separately, yielding *t_norm* and *p_norm*. If the maximum local density for a given cell type within a sample was 0, the corresponding normalized value was set to 0. Based on these normalized variables, we used the joint 2-dimensional distribution of *t_norm* and *p_norm* as a continuous representation of T cell-plasma cell colocalization. Both single-sample and pooled 1-dimensional/2-dimensional distributions were then generated to examine the overall distribution patterns of local densities and to provide a basis for subsequent spatial stratification analyses.

**S3.4** Spatial partitioning based on Gaussian Mixture Model (***GMM***)

Based on *t_norm* and *p_norm* obtained in ***S3.2***, we further applied ***GMM*** to perform unsupervised partitioning of local spatial states within each sample. Specifically, each cell was represented by a two-dimensional feature vector *[t_norm, p_norm]*, and modeled using *sklearn.mixture.GaussianMixture* with *covariance_type="full"* and *random_state = 0*. The number of components was set to *K = 4* as the primary configuration, and AIC/BIC values were recorded for each sample to evaluate model fit. The model output included the cluster assignment for each cell, posterior probabilities, and the centroid position of each cluster in the two-dimensional density space. All results were written back into the original ***anndata*** object to enable downstream analysis with cell annotations and spatial visualization.

To improve interpretability across samples, we further assigned biologically meaningful labels to GMM clusters based on *cluster center*. Specifically, the cluster with the highest *t_norm_center* was defined as the *T_Dominant* region, while the cluster with the highest *p_norm_center* was defined as the *Plasma_Dominant* region. All remaining clusters were categorized as *Mixed* regions. When *T_Dominant* and *Plasma_Dominant* were assigned to the same cluster, a manual review step was performed. If confirmed, the cluster was redefined as an *Engaging Zone* (In SM and MM samples) or *Infiltrating Zone* (In Ctrl and MGUS samples), while other clusters were retained as *Mixed*. The final *cluster annotations*, and *corresponding centroid coordinates* were mapped back to the ***anndata*** object for downstream comparisons of cellular composition, Plasma B-cell Malignancy (***PBM***) ***score***, and T cell functional states across spatial regions.

**S3.5** ***PBM*** ***score*** and regional comparison

To evaluate differences in plasma cell malignancy across spatial regions, we incorporated the ***PBM score*** proposed by Li, et al 2024*. This score is derived from a signature gene set representing normal plasma B-cell biomarkers and quantifies the degree of deviation of plasma cell transcriptomic profiles from a normal plasma B-cell state using a rank-based signature scoring strategy. Higher ***PBM scores*** indicate greater perturbation of plasma B-cell biomarker expression, suggesting increased malignancy. The original study demonstrated that ***PBM score*** serves as a quantitative metric of CD138^+^ plasma cell malignancy and is associated with MM progression and prognosis.

In this study, ***PBM scores*** were mapped onto annotated plasma cells or malignant plasma cells in the spatial transcriptomics dataset. We further compared ***PBM score*** distributions across ***GMM***-defined spatial compartments described in **S3.3**. To avoid confounding effects arising from differences in cellular composition, comparisons of ***PBM scores*** across regions were restricted to the plasma cell lineage, i.e., only cells annotated as plasma cells or malignant plasma cells were included. This allowed us to assess whether *Plasma_Dominant* regions correspond to higher levels of plasma cell malignancy.

*Li, et al., 2024 (In-text citation number 19)

Li JR, Arsang-Jang S, Cheng Y, Sun F, D'Souza A, Dhakal B, et al. Enhancing prognostic power in multiple myeloma using a plasma cell signature derived from single-cell RNA sequencing. Blood Cancer J. 2024;14(1):38.

**S3.6** T cell functional and pathway scoring

To further assess the functional states of T cells in the BME, ***AUCell*** scoring was performed on T cell populations. At the pathway level, cytotoxicity- and exhaustion-related gene sets were retrieved from two GMT files, and only genes overlapping with the Xenium 5K panel were retained. Gene rankings within each cell were constructed using *AUCell_buildRankings* based on normalized expression matrices. Subsequently, *AUC* scores were computed for each gene set at the single-cell level using *AUCell_calcAUC*. The resulting *AUC* matrix was transposed and integrated back into the *meta.data* slot. All score fields were uniformly prefixed with *AUCell* and exported together with *secondary annotation* labels for downstream comparison across T cell subtypes and spatial regions.

**S3.7** Spatial cell-cell communication analysis

Cell-cell communication analysis was performed using CellChat v2.1, which provides optimized support for spatial transcriptomics data (Jin S, et al. 2025*).The analysis followed the standard workflow recommended by the developers, as described in the official documentation (https://github.com/jinworks/CellChat).

* Jin S, et al., 2025 (In-text citation number 20)

Jin S, Plikus MV, Nie Q. CellChat for systematic analysis of cell-cell communication from single-cell transcriptomics. Nat Protoc. 2025;20(1):180-219.

**S3.8** Pseudobulk differential expression analysis

For each sample, raw Xenium counts were aggregated by niche and major cell type. Specifically, all cells belonging to the same sample-niche-cell type combination were summed to construct pseudobulk expression profiles. This aggregation was performed independently within each disease group and for each major cell type. For selected immune-related genes, pseudobulk expression values were computed using the same sample-niche-cell type aggregation strategy. The aggregated counts were normalized to counts per 10,000 (CP10K) and subsequently log-transformed as *log2(CP10K + 1)* for downstream comparison.

**S3.9** Construction of the multilayer perceptron (***MLP***) model

scRNA-seq datasets often lack spatial context , therefore not applicable for spatial architecture identification , like exhaustion-like bone marrow T-cell islands (eBM-TIs) in our research. However, from our Xenium data anlysis, we confirmed that cells localized in certain region might carry identical transcriptomic features, which could be utilized for spatial information prediction. We developed a machine learning-assisted spatial niche transfer framework to evaluate whether transcriptional signatures associated with eBM-TIs could be captured in independent single-cell cohorts.

Gene expression readouts between Chromium Single-cell and Xenium in situ platform is similar according to 10X Genomics. In Xenium In Situ Grant Application Resource*, both Chromium single-cell and Xenium in situ data were generated from the tissue block, and readouts between these 2 platforms were compared. After analyzing data from 2 platforms individually, similar t-SNE projection and celltype annotation results were obtained using genes existed in both platform. These technical observations supported the feasibility of using Xenium-derived gene expression profiles as the training feature space and applying the trained model to external single-cell gene expression datasets after restricting features to shared genes.

Specifically, spatial niche labels defined by ***GMM*** in the Xenium dataset were used as training labels. A ***MLP*** classifier was constructed to capture transcriptional features corresponding to *T_Dominant*, *Plasma_Dominant*, *Mixed*, and related spatial niche states. The trained model was then applied to previously published MM scRNA-seq datasets to estimate, at the single-cell level, the probability of each T cell being transcriptionally similar to eBM-TIs-associated spatial niches. The ***MLP*** model generated continuous soft niche probability scores for both T cells and plasma cells, enabling the reconstruction of *spatial niche similarity* in datasets without spatial coordinates. At the patient level, we further quantified the proportions of *T_Dominant*-like, *Plasma_Dominant*-like, and *Mixed*-like cells based on these probability scores.

*Xenium In Situ Grant Application Resource, latest version:

https://assets.10xgenomics.com/m/29745c7439be2b5d/original/10x_LIT000166_Grant-Application_Xenium_Letter_Digital.pdf

The version referenced in this study was the version dated June 23, 2026, which is provided as ***Supplementary File 5***.

**S4** Additional analytical considerations

**S4.1** Identification of an SOX2-OT^high^ undefined cluster

Across clustering results at multiple resolutions, a SOX2-OT^high^ cluster was consistently identified. Based on differential expression analysis using *sc.tl.rank_genes_groups*, this cluster could not be assigned to any well-defined cell type and did not match known canonical biomarker gene signatures. This cluster was characterized by generally low overall transcript abundance, with the exception of SOX2-OT, and lacked a stable set of highly expressed biomarker genes. Therefore, it did not conform to the typical expression patterns expected for canonical biological cell populations.

To further evaluate its biological nature, we applied *spacexr::run.RCTD* for deconvolution analysis. All cells within this cluster were consistently classified as *reject* by ***RCTD***. In addition, ***BANKSY***-based spatial clustering across multiple samples repeatedly identified this cluster as a stable but independent group.

Taken together, these results suggest that the SOX2-OT^high^ cluster most likely represents a low-information signal dominated by ambient RNA contamination rather than a biologically distinct cell population. However, completely removing this cluster would potentially disrupt the continuity of spatial information, potentially biasing local density estimation and spatial boundary definition. To avoid introducing artifacts caused by excessive filtering, we retained this population as the *SOX2-OT Unknown* subcluster and included it in downstream analyses.
