## Supplementary File 3 for "Spatial transcriptomics and machine learning define exhaustion-like bone marrow T-cell islands associated with myeloma progression and clinical risk"

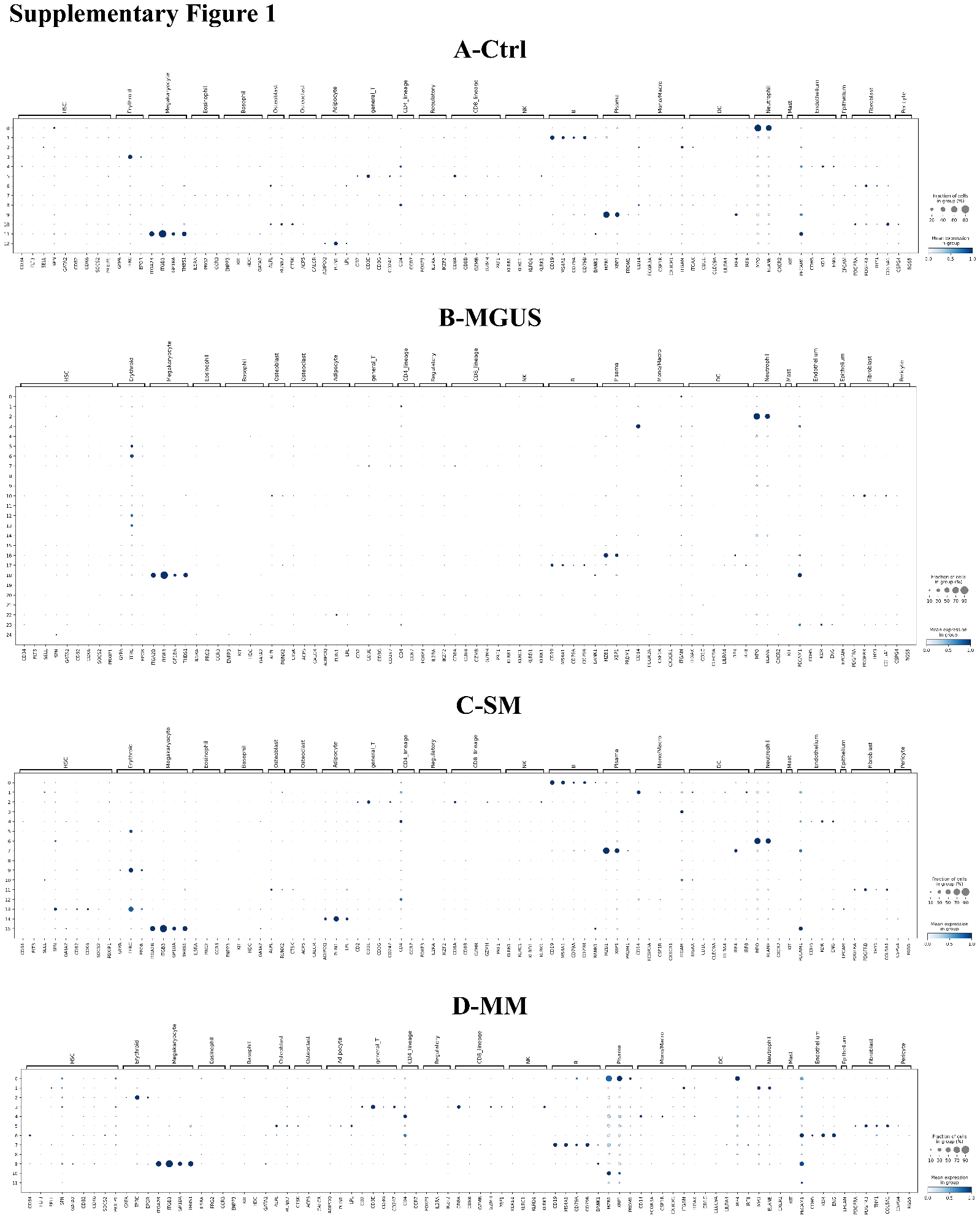
**Supplementary Figure 1** Dotplot showing clusters expressing major celltype biomarker genes in BME, used for celltype annotation

(A-D) Dotplot showing biomarker gene expression of major cell types in Ctrl, MGUS, SM, and MM samples.

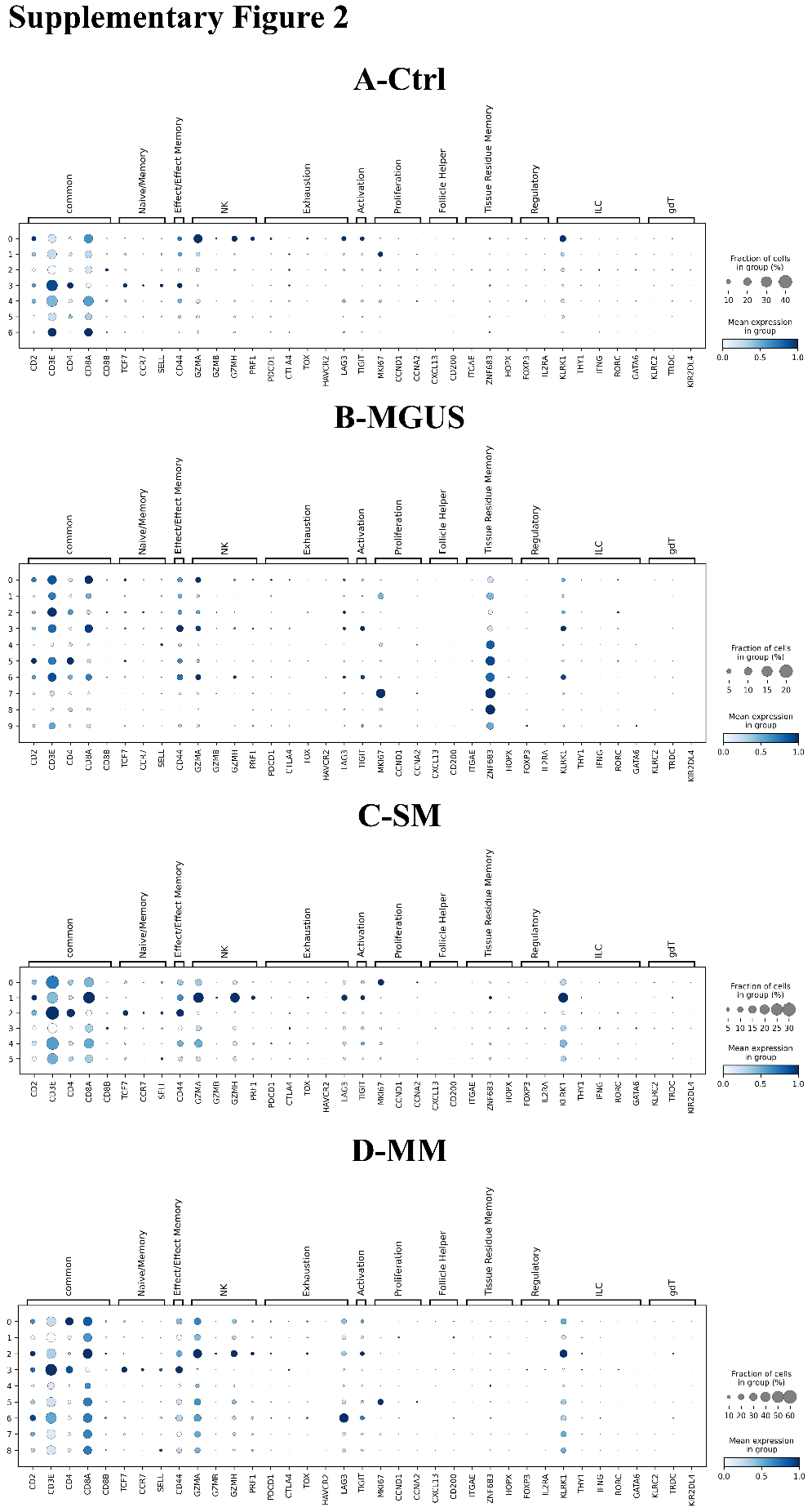
**Supplementary Figure 2** Expression distribution of biomarker genes for T cell subpopulation annotation

(A-D) Dotplot showing biomarker genes corresponding to T cell subpopulations in Ctrl, MGUS, SM, and MM samples.

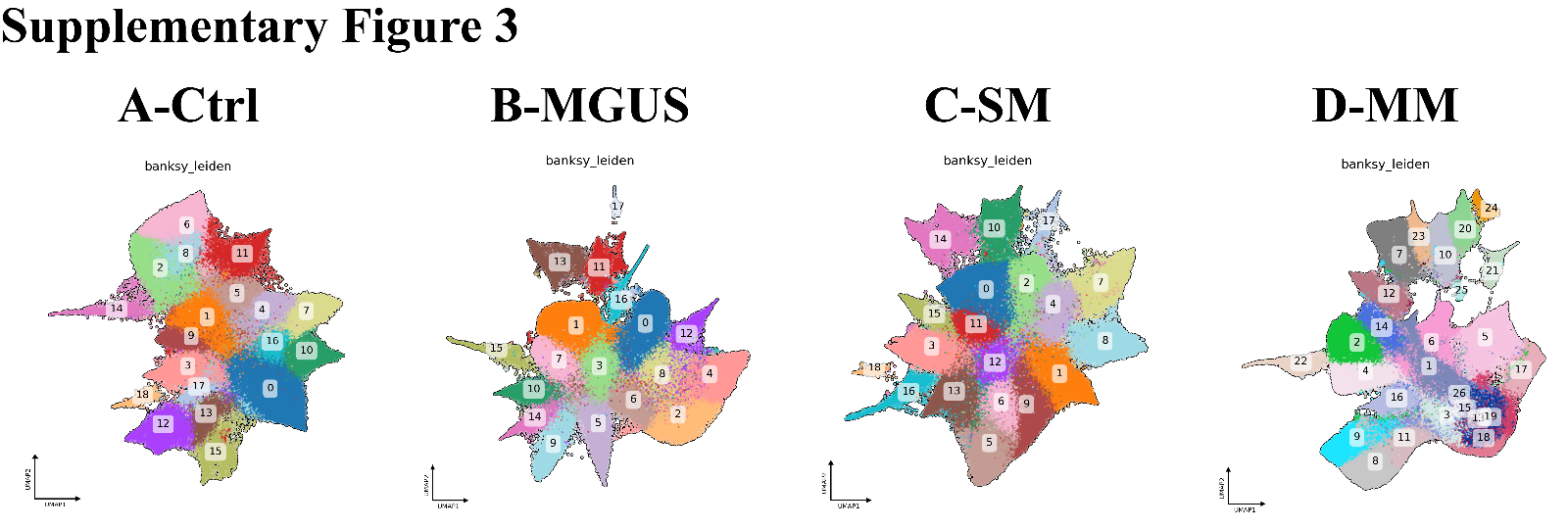
**Supplementary Figure 3** BANKSY spatial clustering results

(A-D) UMAP plots of BANKSY-based Leiden clustering in Ctrl, MGUS, SM, and MM samples.

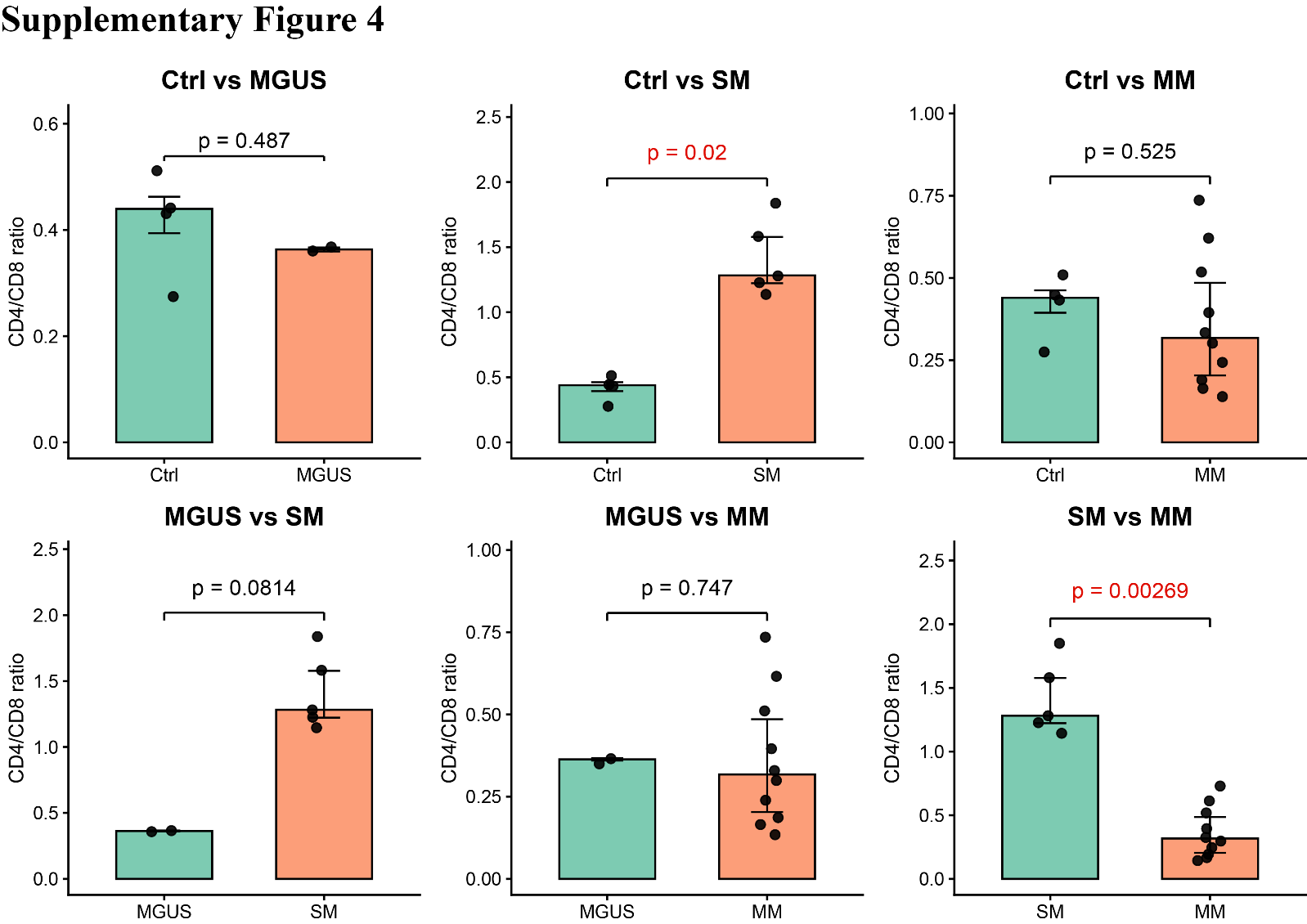

**Supplementary Figure 4** Comparison of CD4/CD8 ratio across disease stages

Pairwise comparisons of CD4+/CD8+ T cell ratios across Ctrl, MGUS, SM, and MM stages. P-values are indicated in the figure, calculated using Wilcoxon rank-sum test.

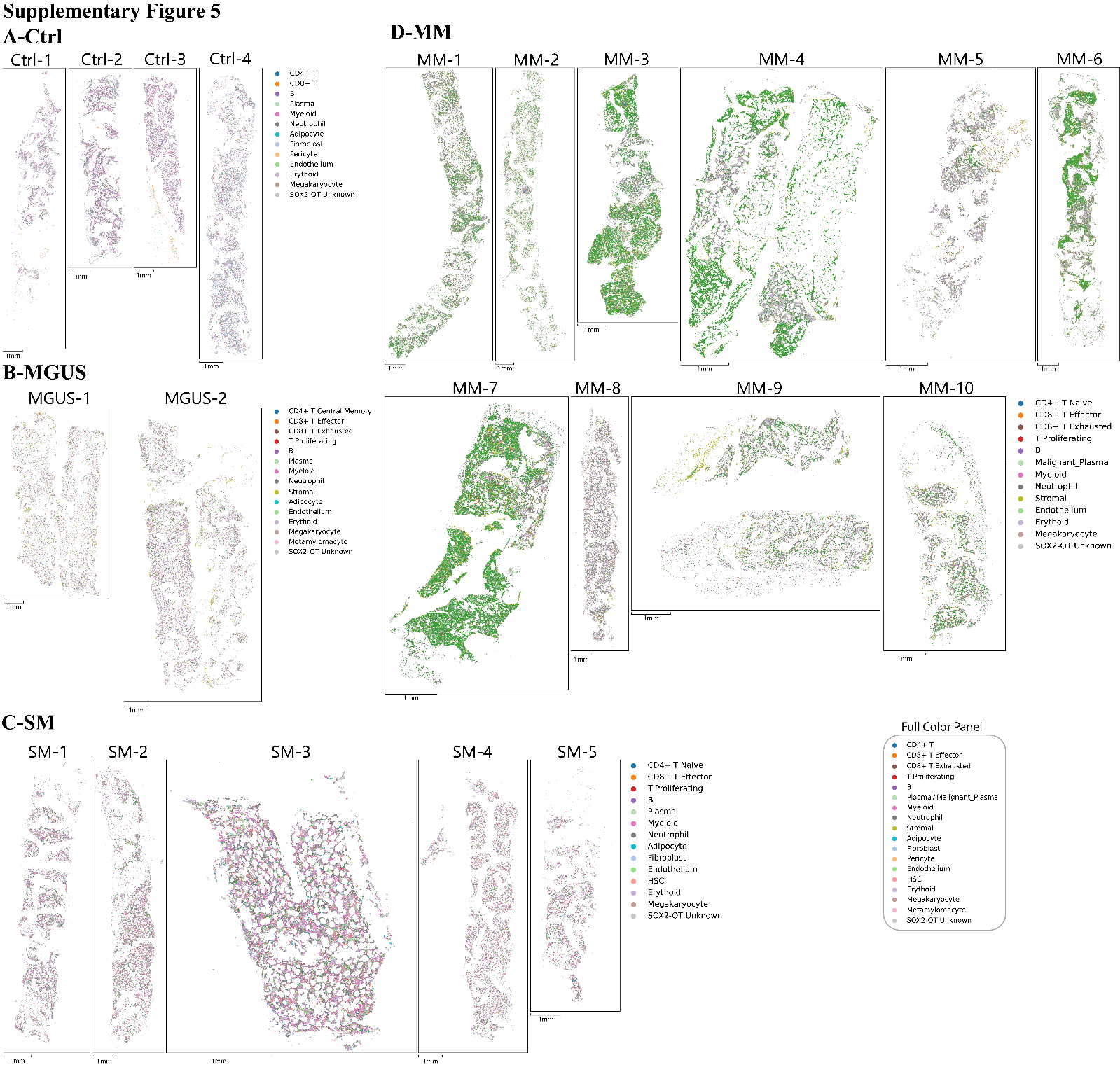
**Supplementary Figure 5** Spatial distribution of major cell types across all samples

(A-D) Spatial maps of major cell types in Ctrl, MGUS, SM, and MM samples.

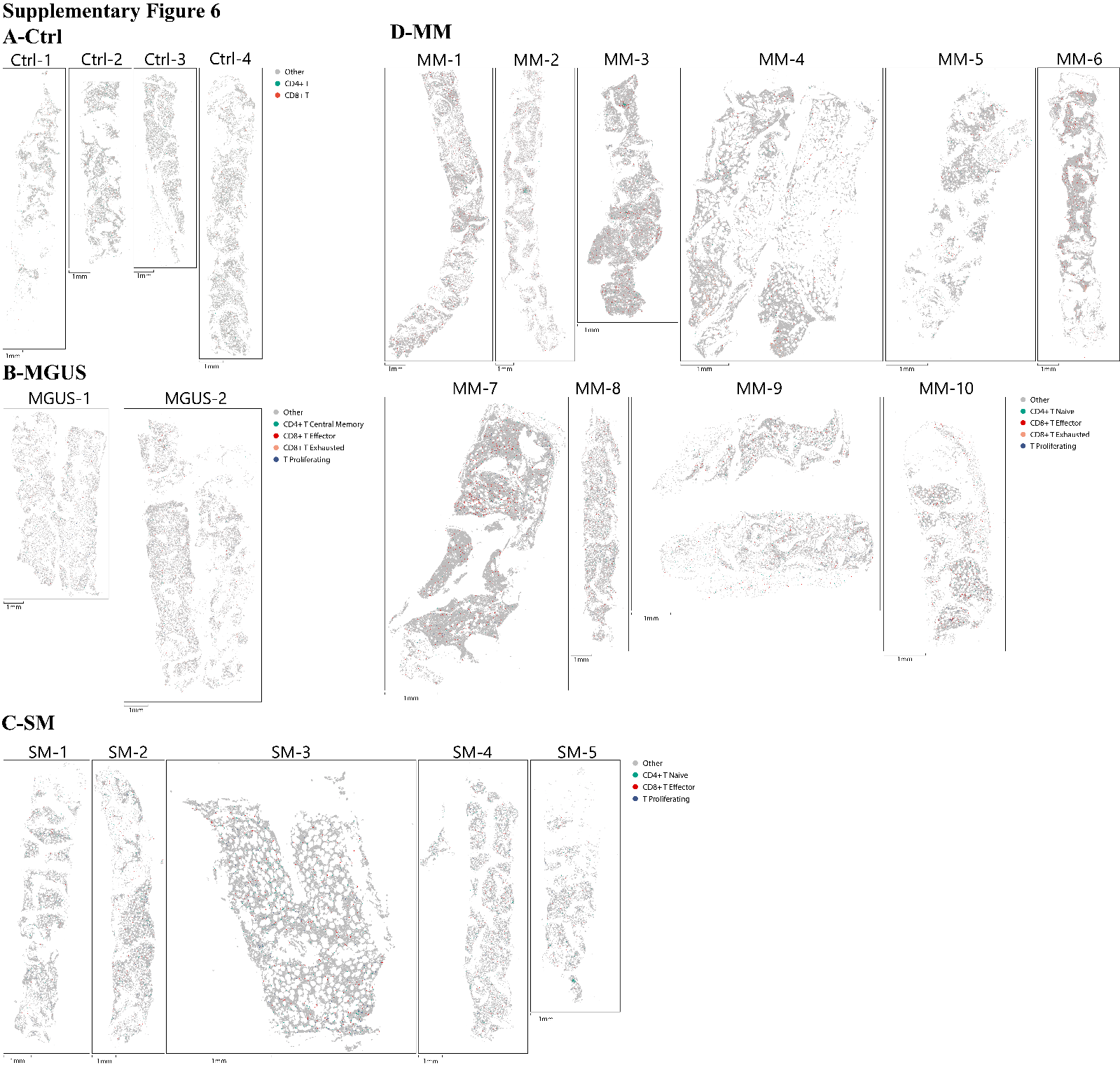
**Supplementary Figure 6** Spatial highlight of T cell subpopulations across all samples

(A-D) Spatial highlight maps of T cell subpopulations in Ctrl, MGUS, SM, and MM samples. Grey indicates non-T cells, and colored polygons represent different T cell subtypes.

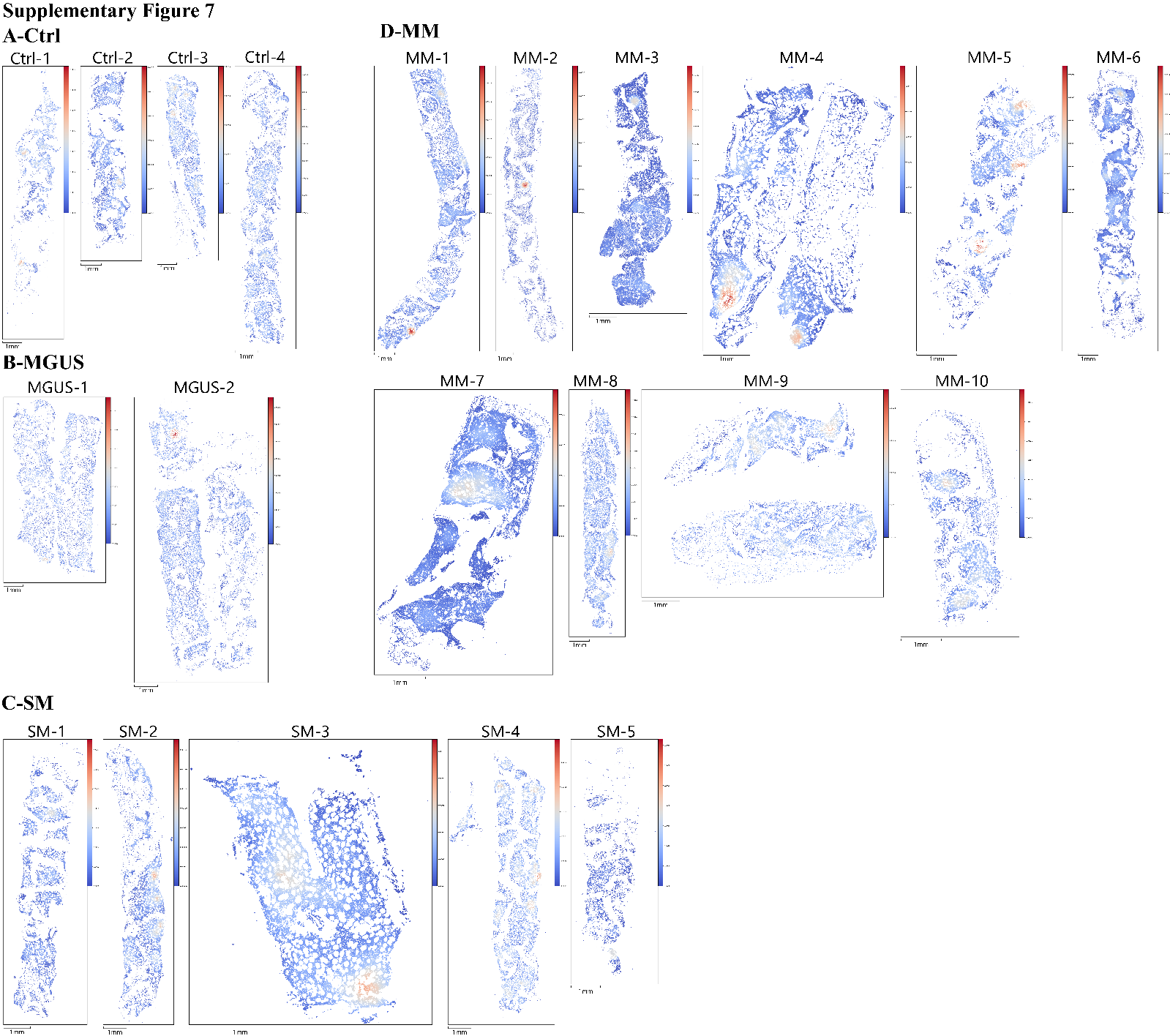
**Supplementary Figure 7** Spatial distribution of local T cell density across all samples

(A-D) Spatial maps showing local T cell density in Ctrl, MGUS, SM, and MM samples.

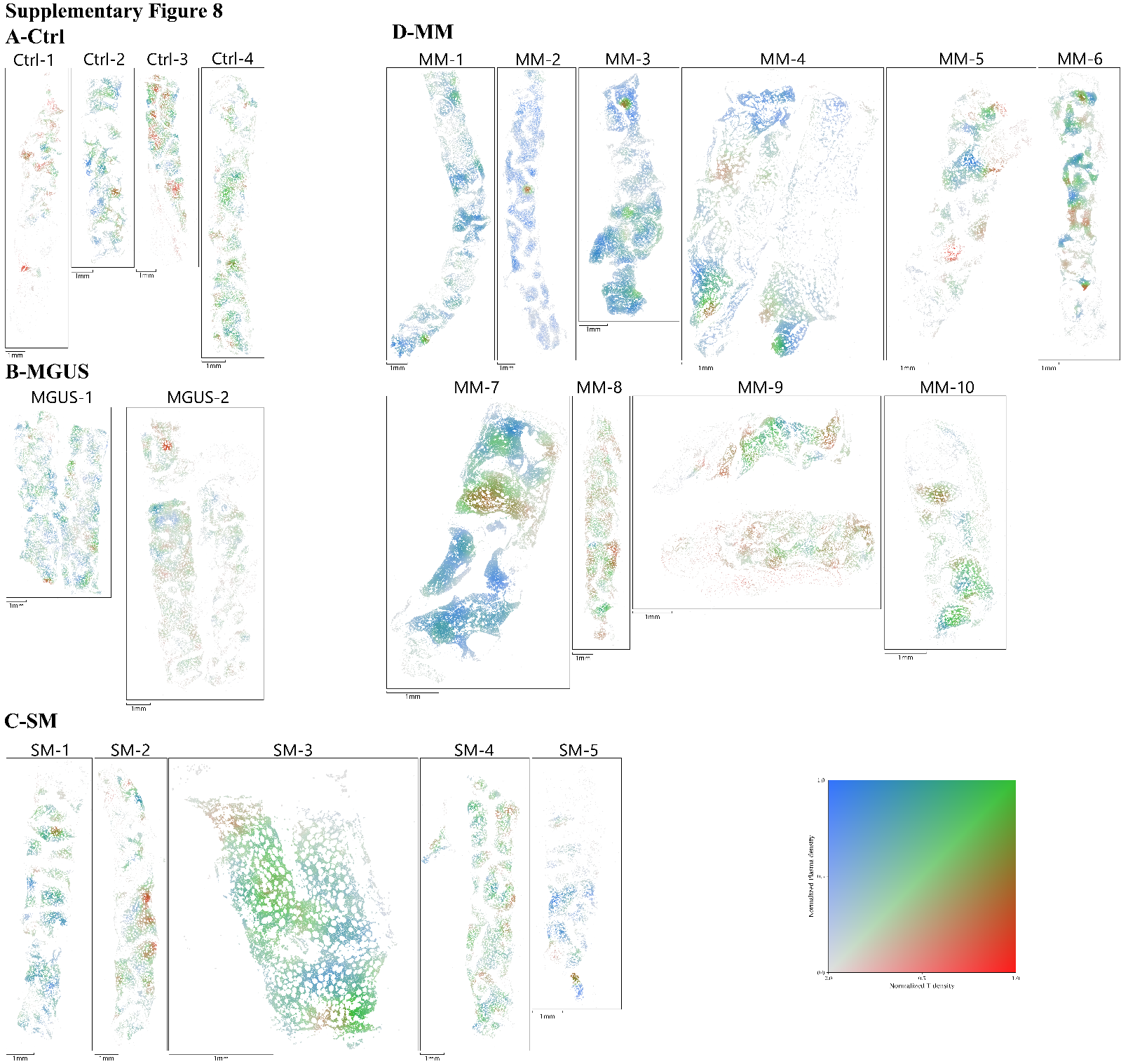
**Supplementary Figure 8** Joint spatial distribution of T cell and plasma cell local density across all samples

(A-D) Bivariate spatial maps of local T cell density and plasma cell density in Ctrl, MGUS, SM, and MM samples.

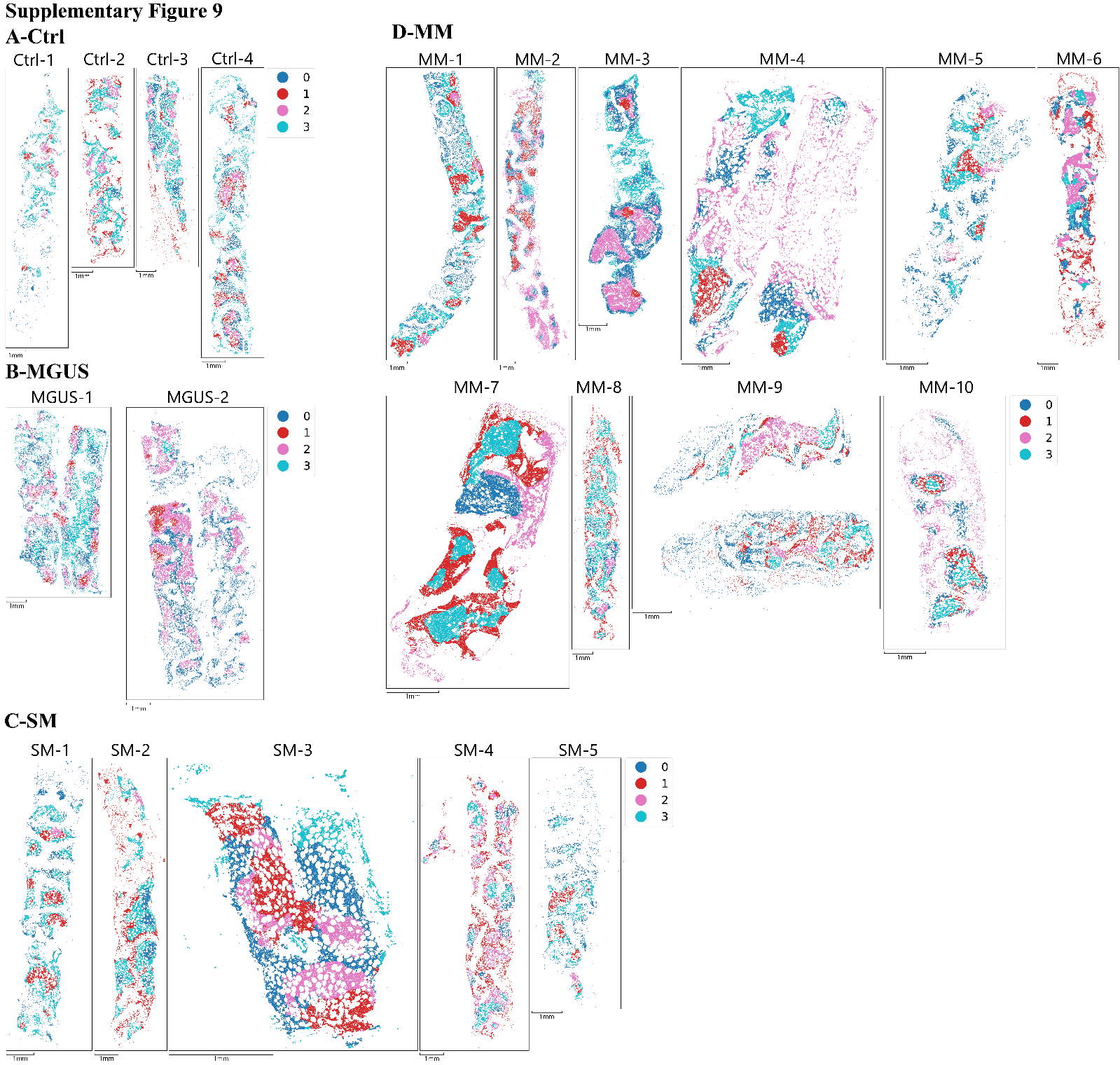
**Supplementary Figure 9** GMM-based unsupervised spatial clustering across all samples

(A-D) GMM clustering results based on local T cell and plasma cell density in Ctrl, MGUS, SM, and MM samples.

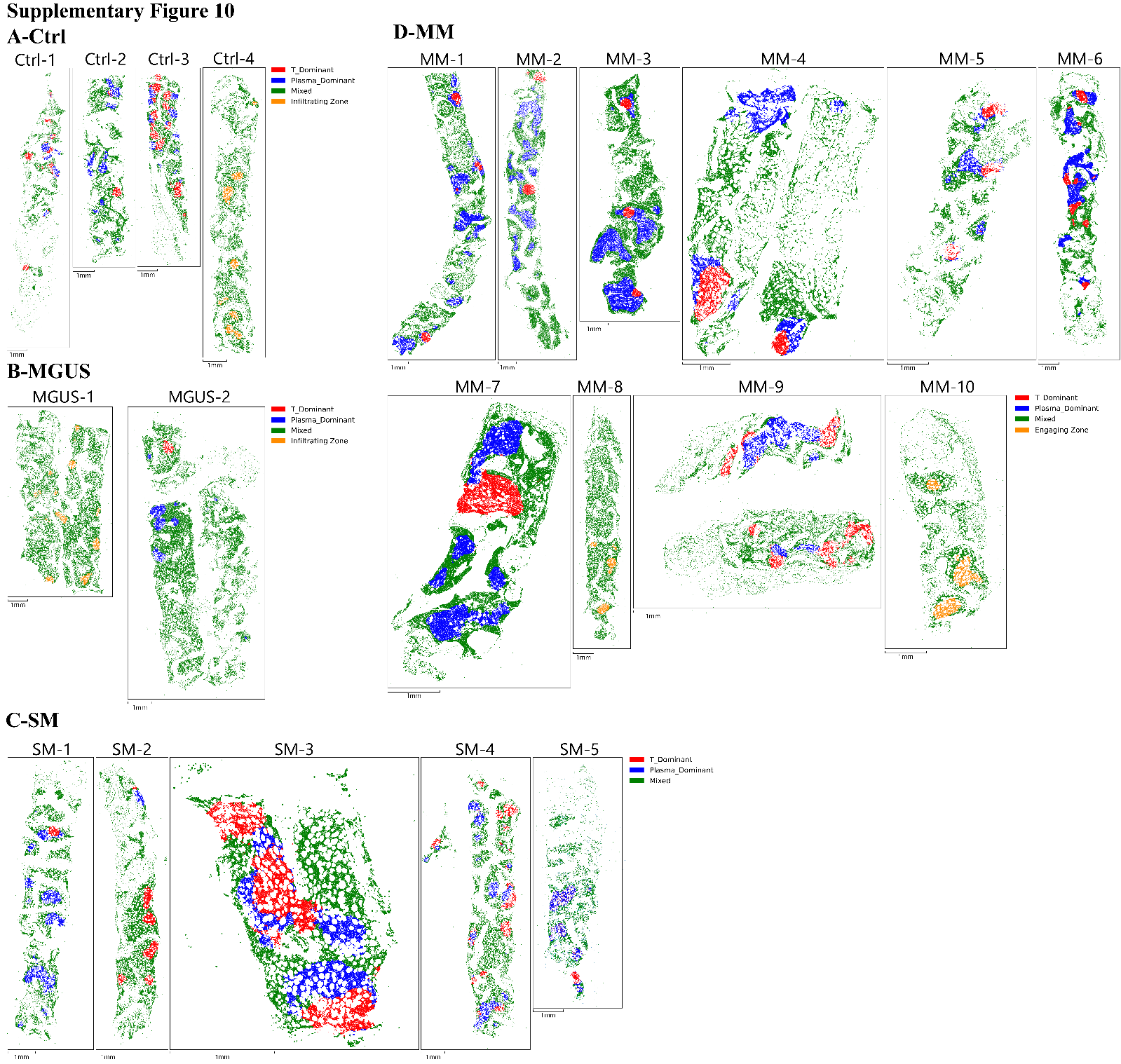
**Supplementary Figure 10** Annotated spatial niches derived from GMM clustering across all samples

(A-D) Spatial niche annotations derived from GMM clustering in Ctrl, MGUS, SM, and MM samples, including T_Dominant, Plasma_Dominant, Mixed, and additional niche states.

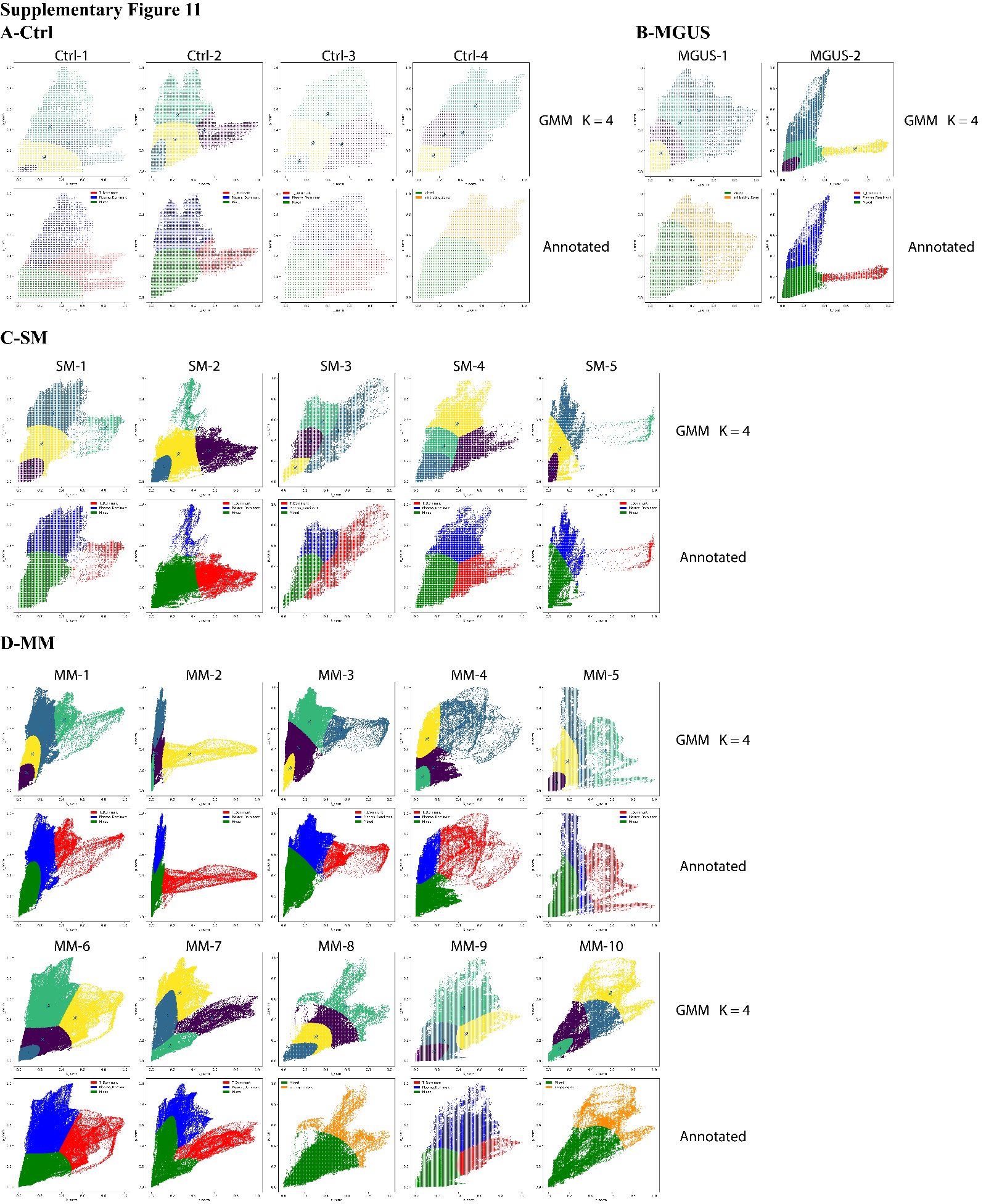
**Supplementary Figure 11** GMM clustering in 2-dimensional density space with interpretation

(A-D) GMM K=4 clustering results in local T cell and plasma cell density space (top row) and corresponding annotated spatial niches (bottom row) in Ctrl, MGUS, SM, and MM samples.

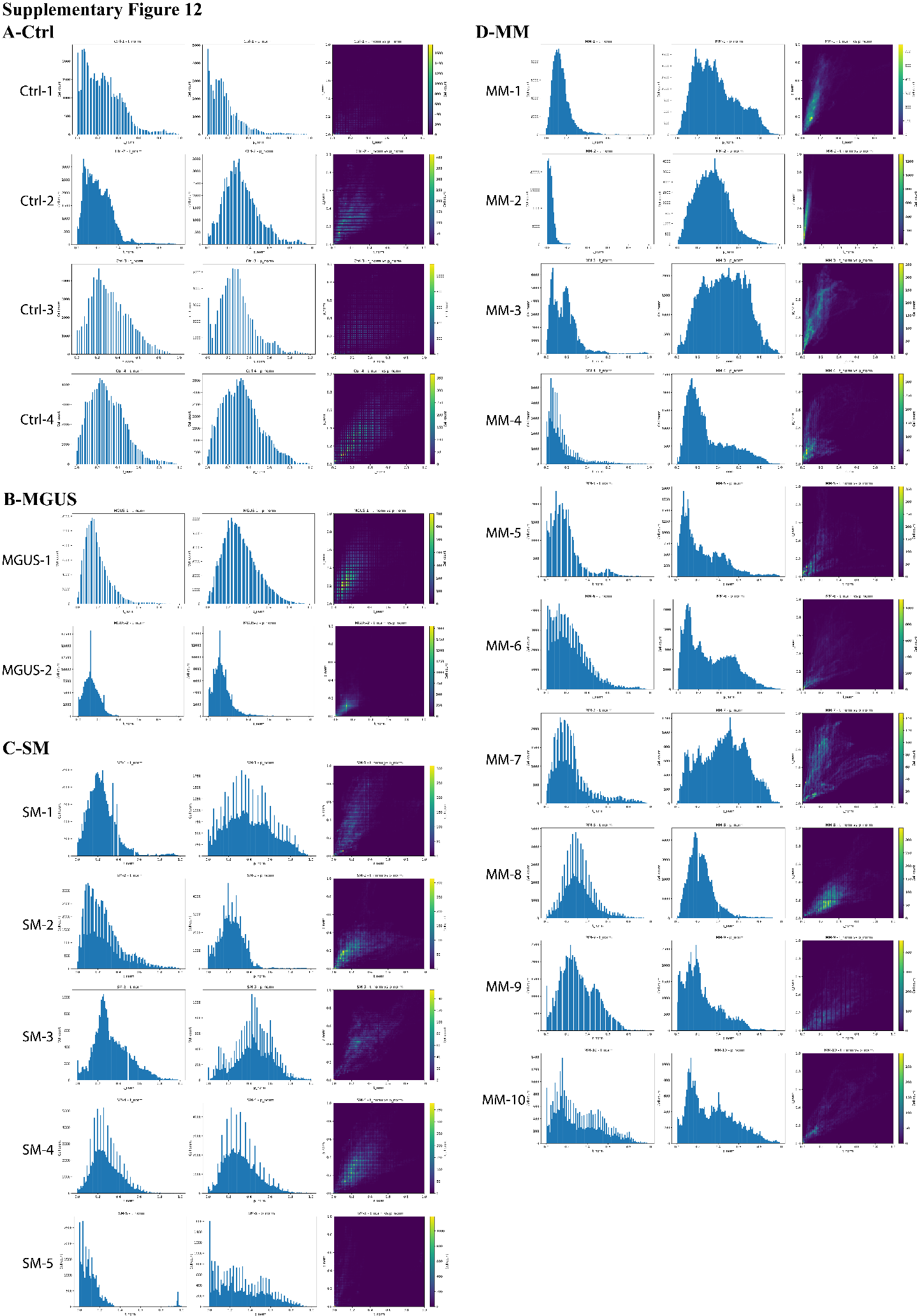
**Supplementary Figure 12** Distribution of input features for GMM clustering across samples

(A-D) Distributions of local T cell density (first column), local plasma cell density (second column), and joint density distribution (third column) in Ctrl, MGUS, SM, and MM samples.

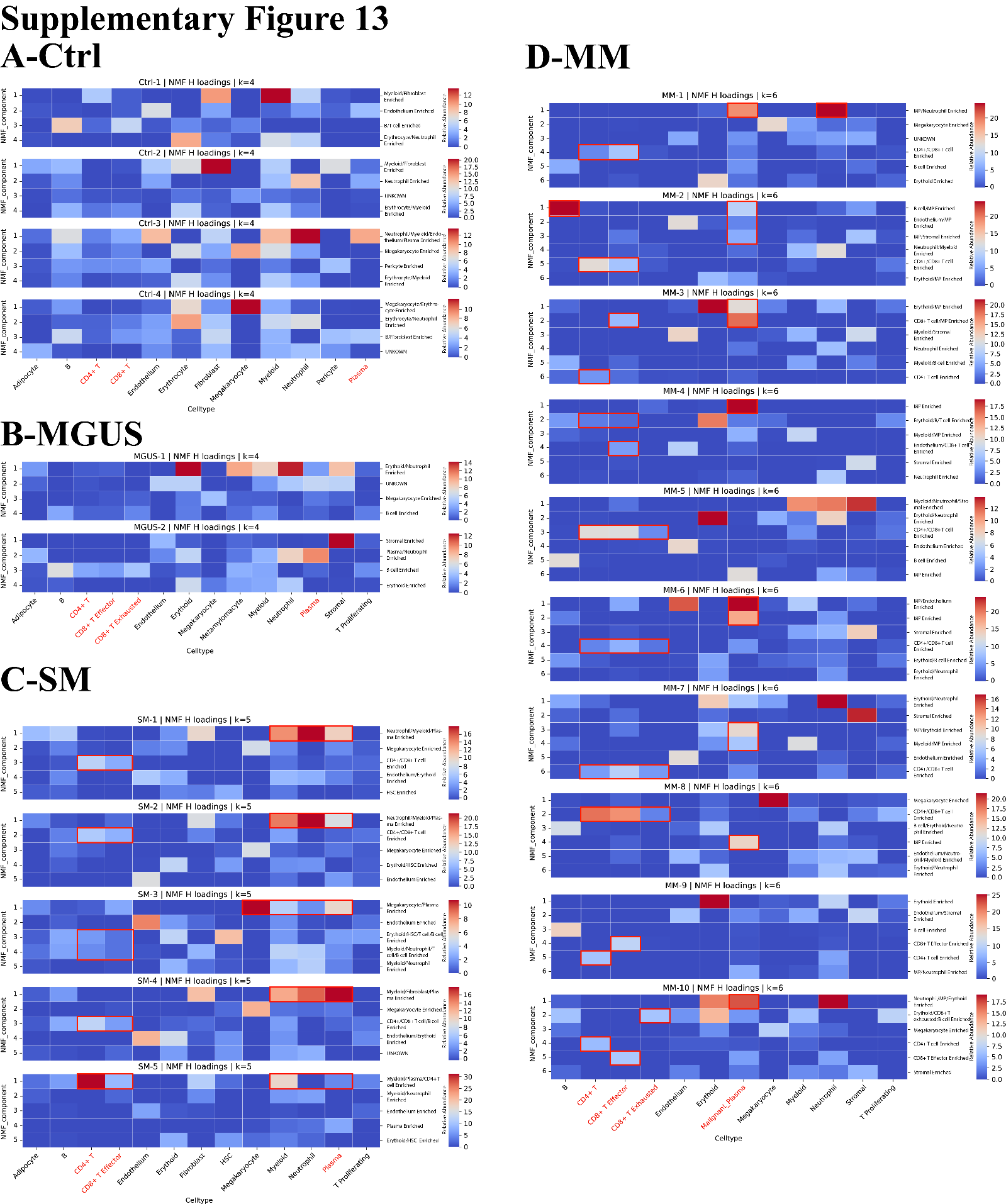
**Supplementary Figure 13** NMF-based neighborhood composition analysis validating spatial niche structure

(A-D) Heatmaps of NMF H-loading matrices based on neighborhood cell composition in Ctrl, MGUS, SM, and MM samples.

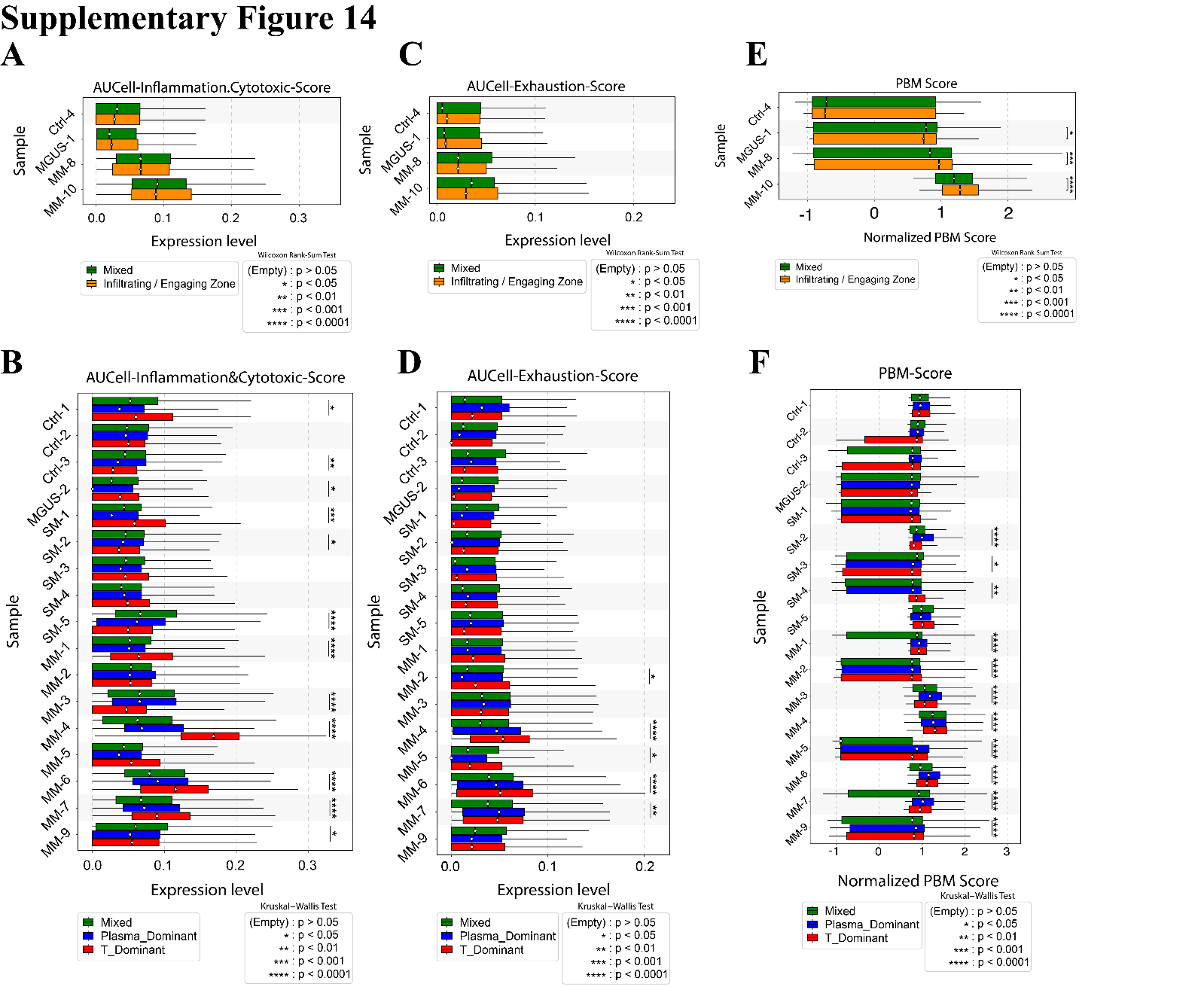
**Supplementary Figure 14** Functional scoring of T cells and PBM score analysis across spatial niches

(A, C, E) Comparison of AUCell inflammation/cytotoxic scores, AUCell exhaustion scores, and PBM scores between Mixed and Infiltrating/Engaging regions.

(B, D, F) Comparison of AUCell inflammation/cytotoxic scores, AUCell exhaustion scores, and PBM scores across Mixed, Plasma_Dominant, and T_Dominant regions.

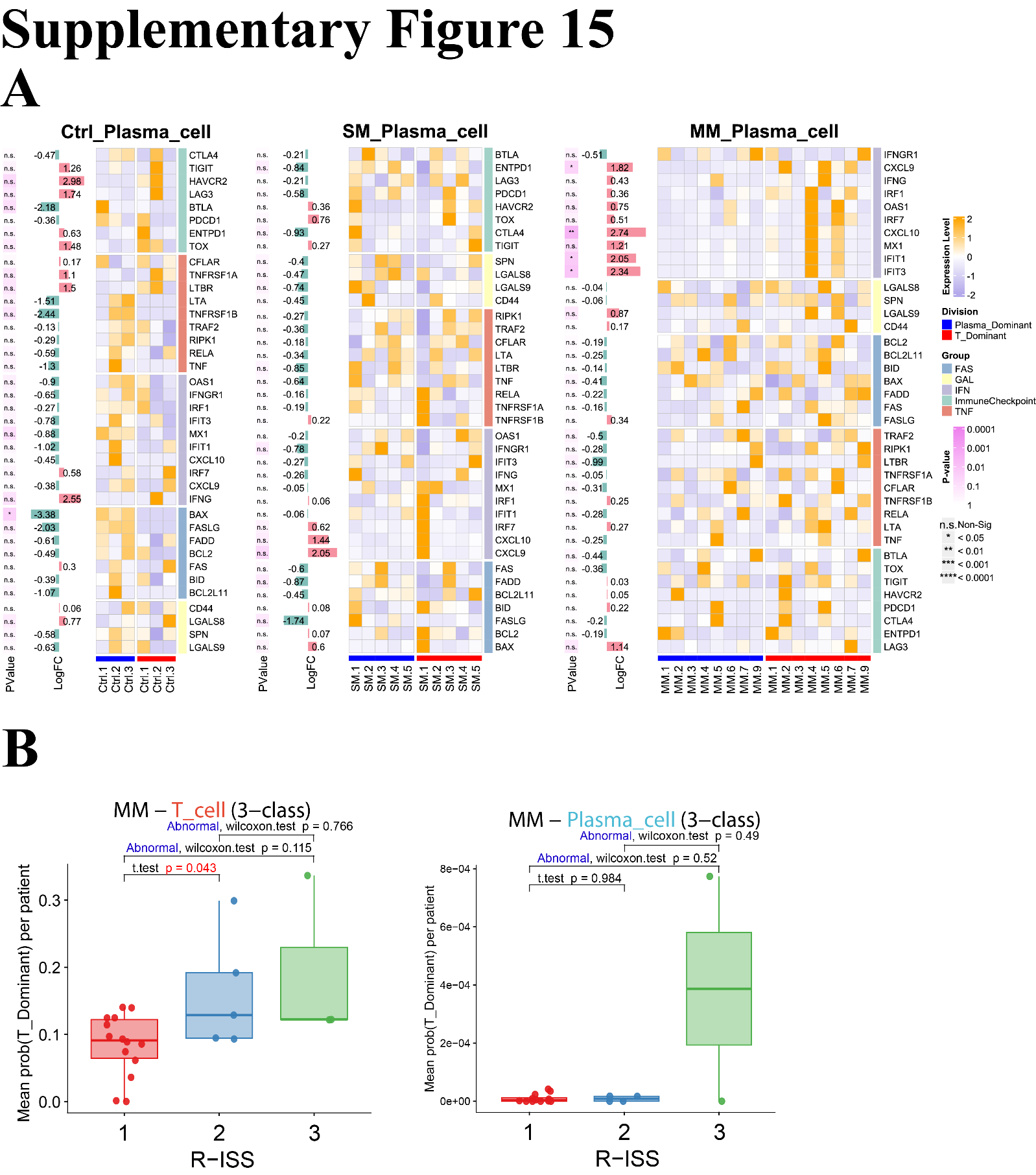
**Supplementary Figure 15** Molecular features of plasma cell niches and MLP-based spatial transfer analysis

(A) Pseudobulk differential expression analysis of plasma cells across T_Dominant and Plasma_Dominant niches in Ctrl, SM, and MM samples.

(B) Comparison of T_Dominant-like soft niche probabilities for T cells and plasma cells stratified by R-ISS stage in an external MM scRNA-seq cohort.

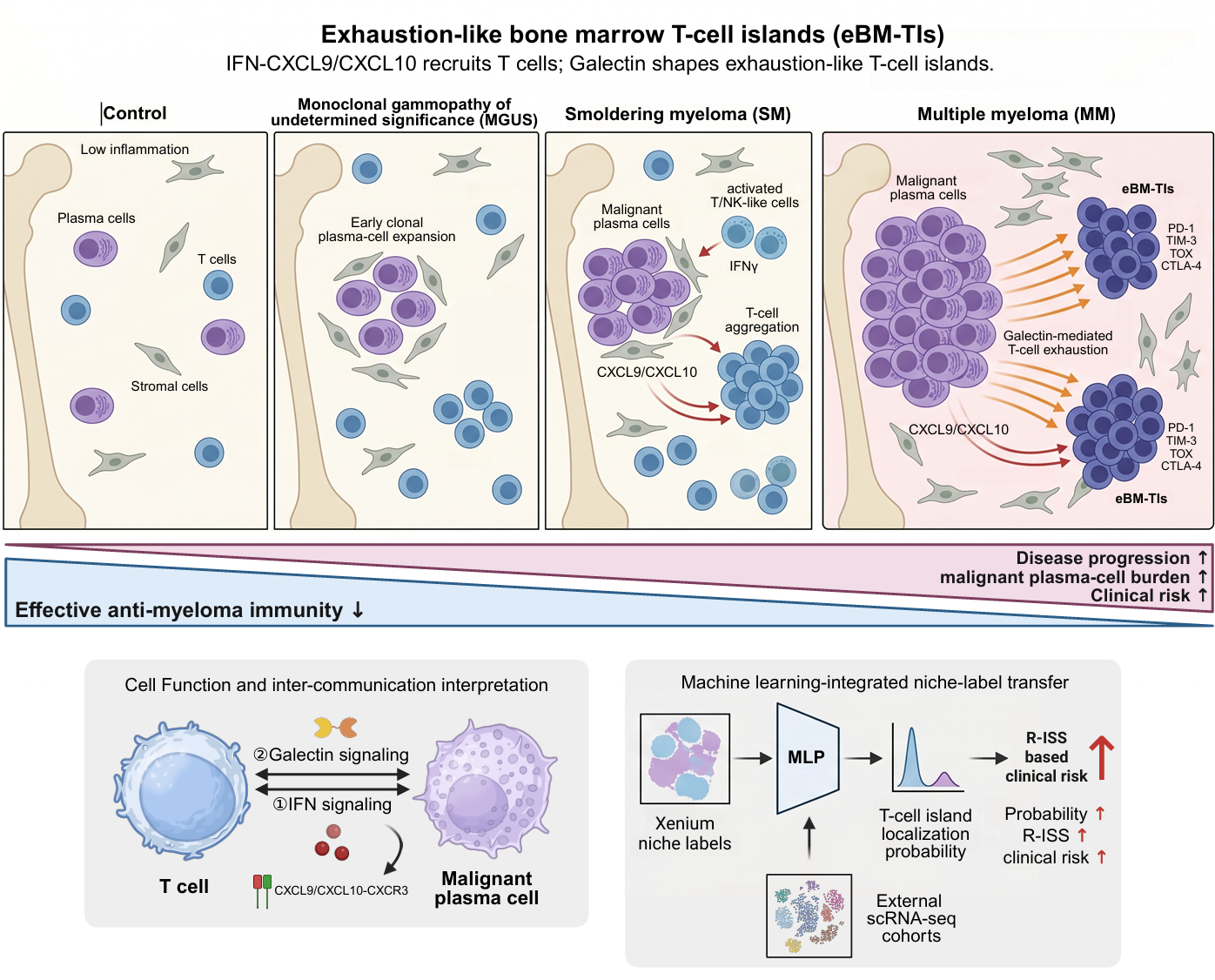
**Graphical Abstract**

**Graphical Abstract**

Li XY, Jiang XL, et al. identified a population of exhaustion-like T-cell islands in the bone marrow microenvironment of multiple myeloma. These spatial immune niches are characterized by increased abundance during disease progression, enhanced Galectin-mediated T-plasma cell interactions, and IFN-CXCL9/CXCL10-associated inflammatory chemotaxis. Their biological features and association with clinical parameters collectively indicate that exhaustion-like T-cell islands represent a key immune unit linking chronic inflammation, T-cell exhaustion, and clinical risk in multiple myeloma, and may serve as a potential biomarker of disease progression.
