## Supplementary File 4 for "Spatial transcriptomics and machine learning define exhaustion-like bone marrow T-cell islands associated with myeloma progression and clinical risk": Graphical_Abstract.pdf

### Exhaustion-like bone marrow T-cell islands (eBM-TIs)

IFN-CXCL9/CXCL10 recruits T cells; Galectin shapes exhaustion-like T-cell islands.

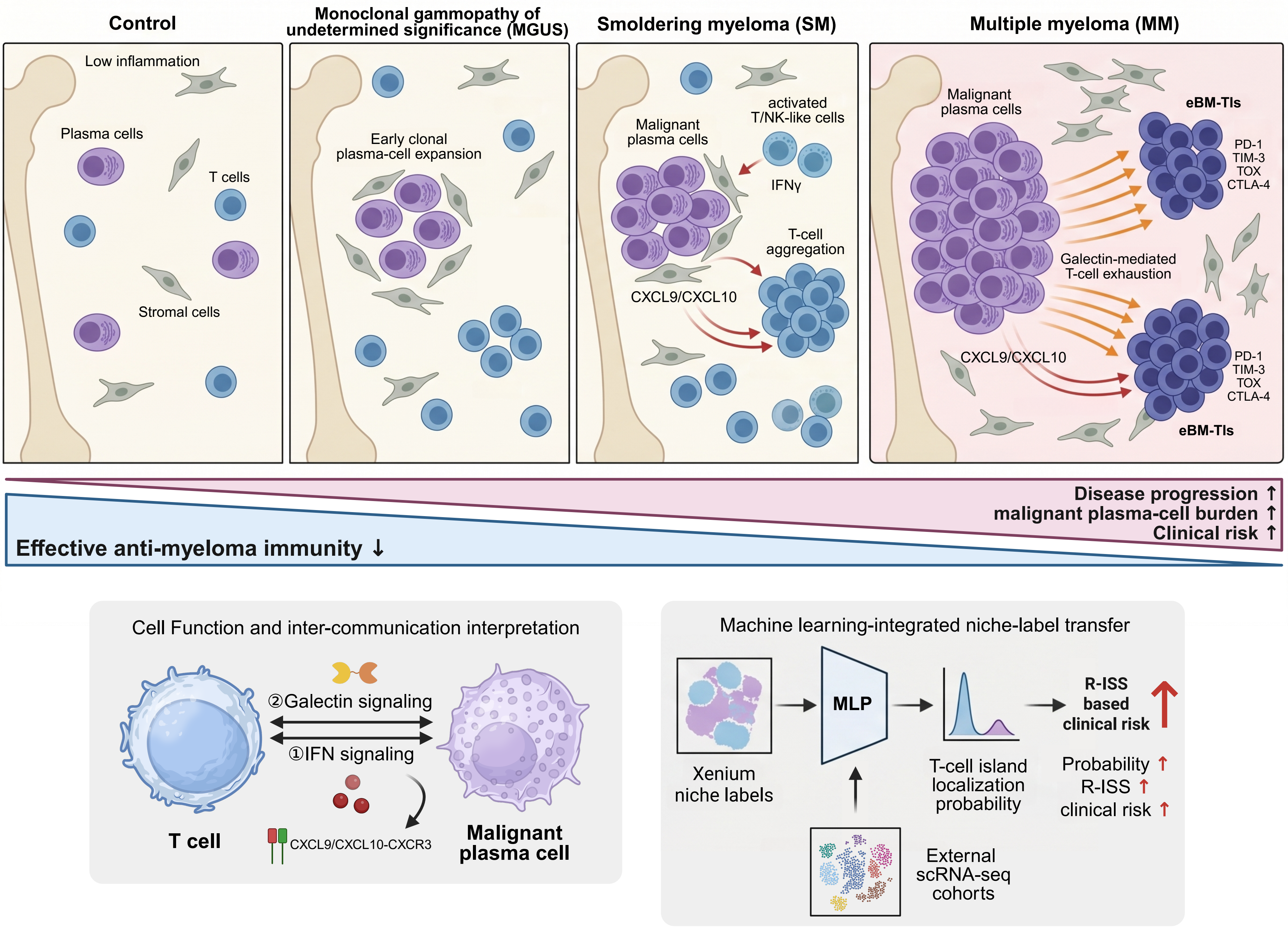
