## Supplementary figures and images for "Spatial transcriptomics and machine learning define exhaustion-like bone marrow T-cell islands associated with myeloma progression and clinical risk"

### Figure 1.pdf

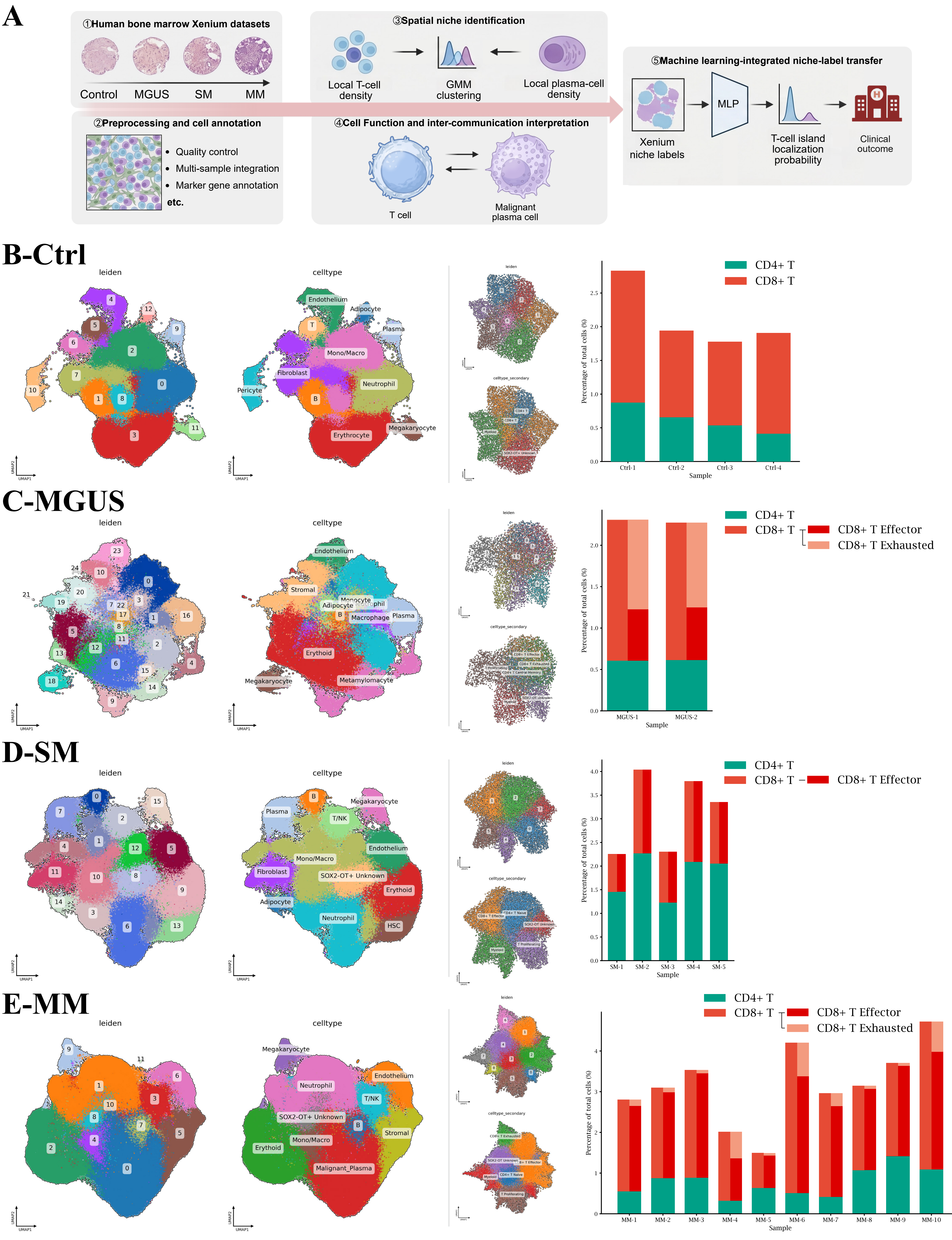

### Figure 2.pdf

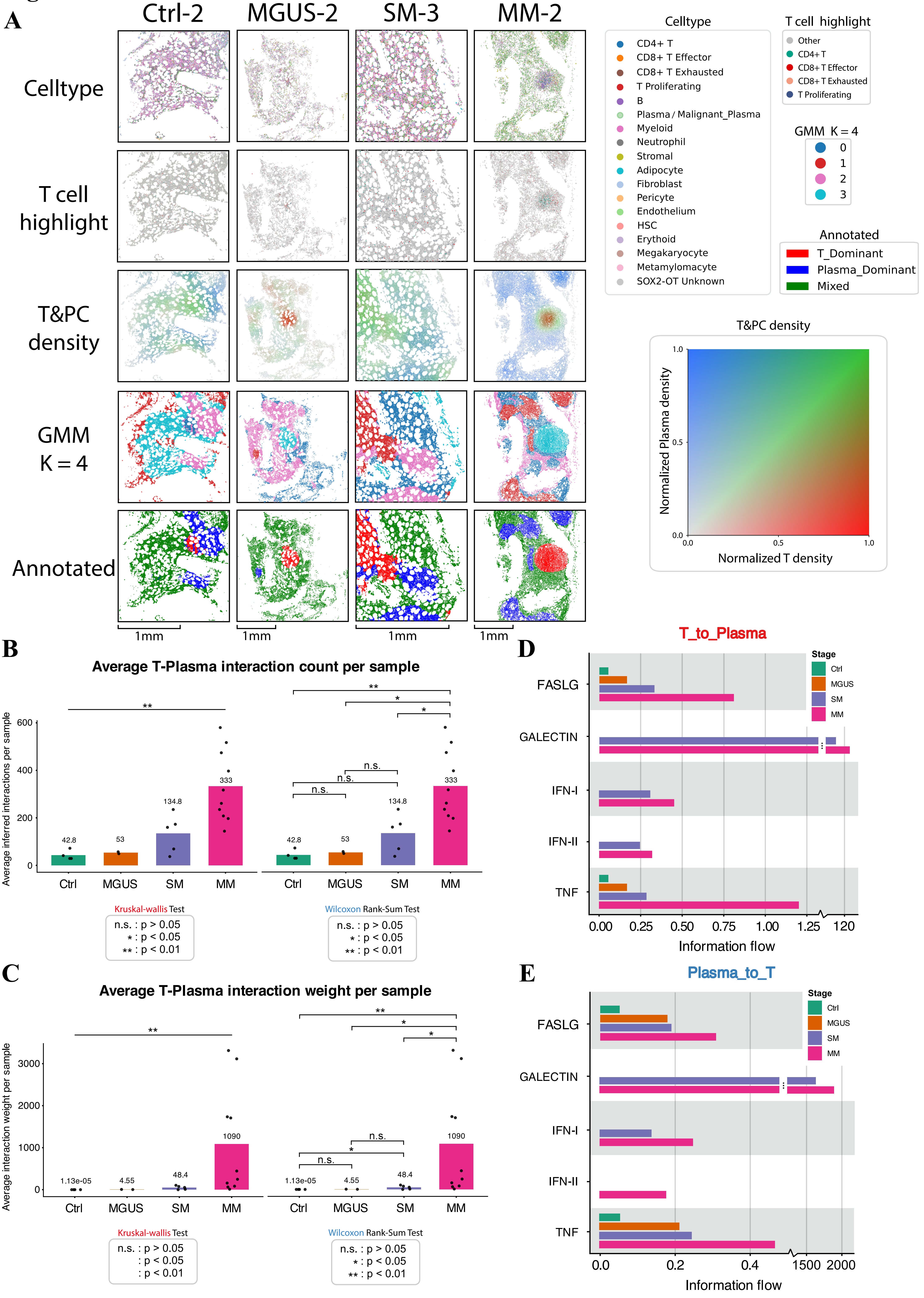

### Figure 3.pdf

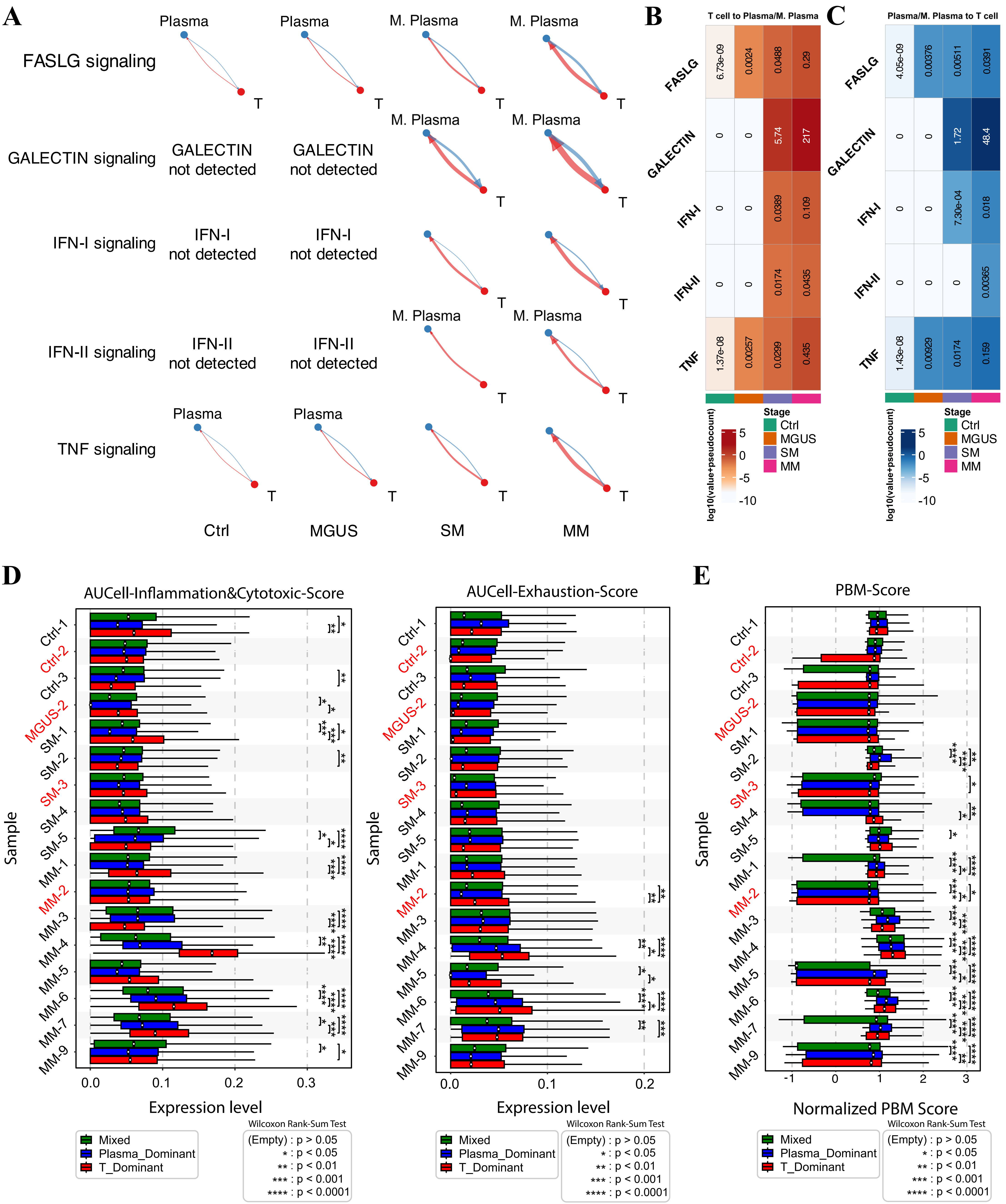

### Figure 4.pdf

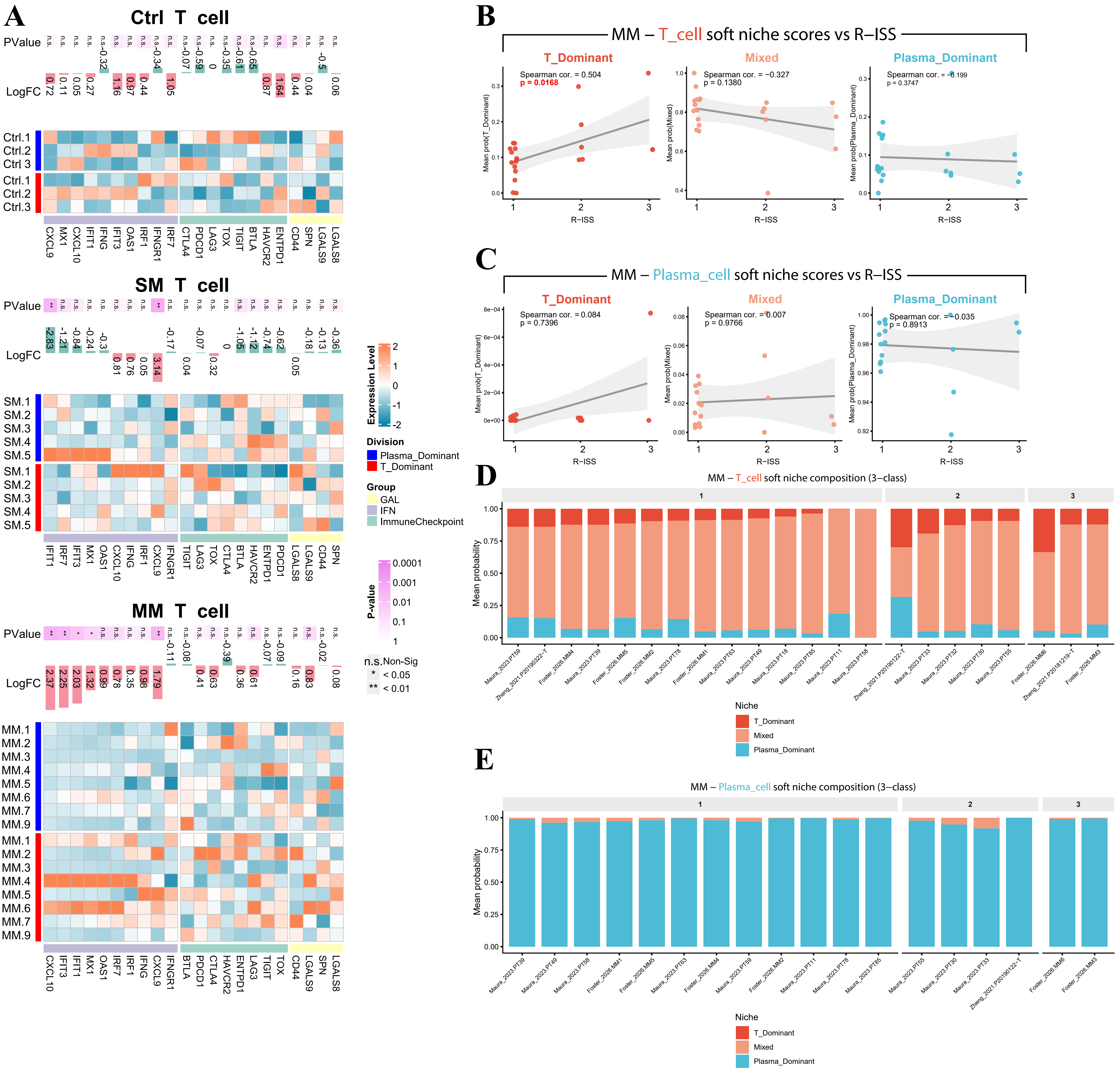

### Supplementary Figure 1.pdf

#### Supplementary Figure 1

##### A-Ctrl

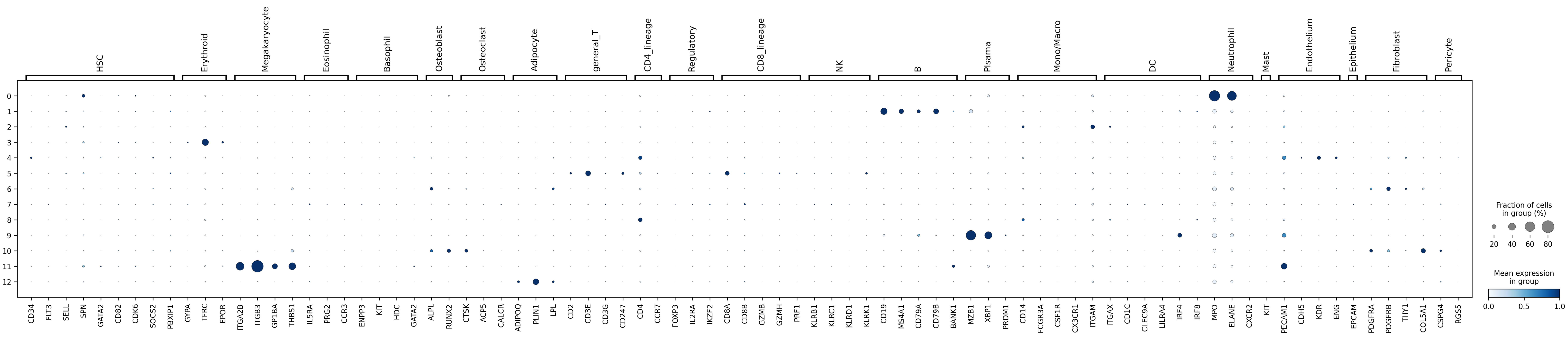


##### B-MGUS

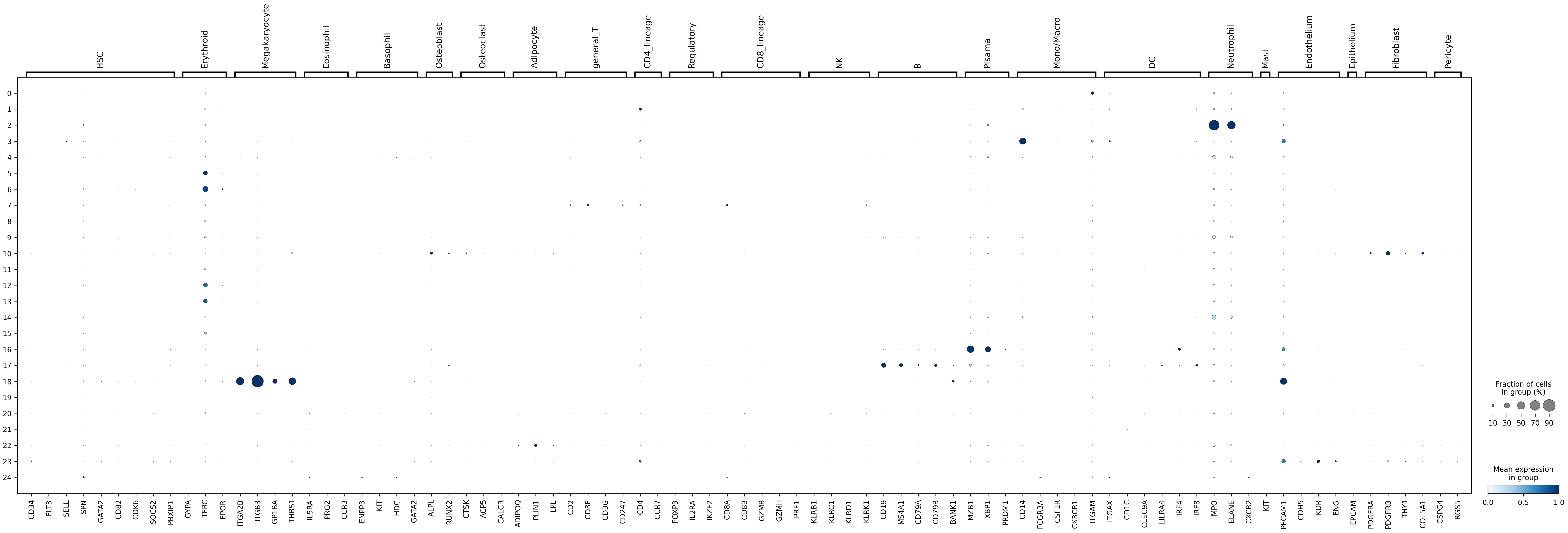


## C-SM

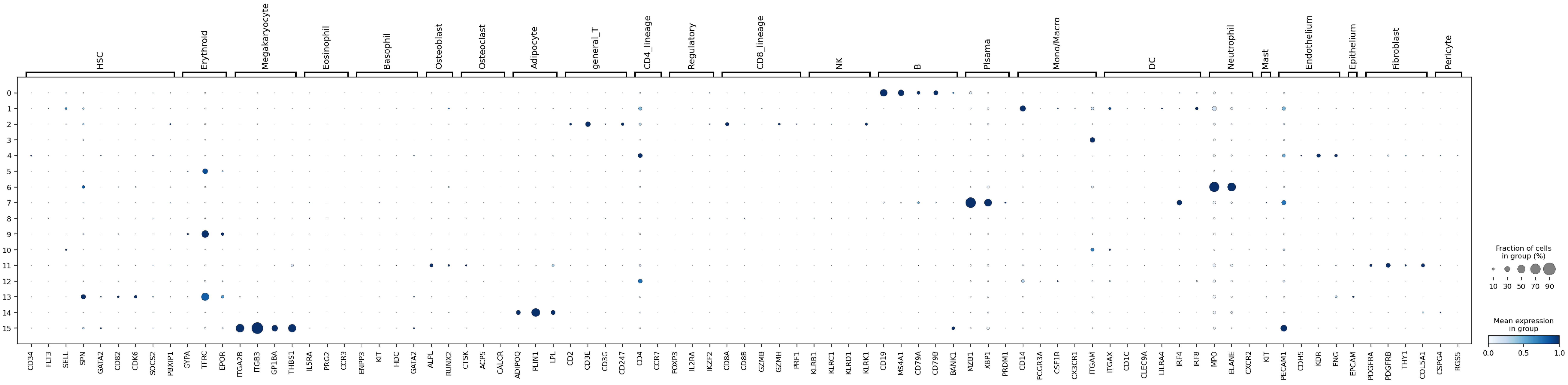


## D-MM

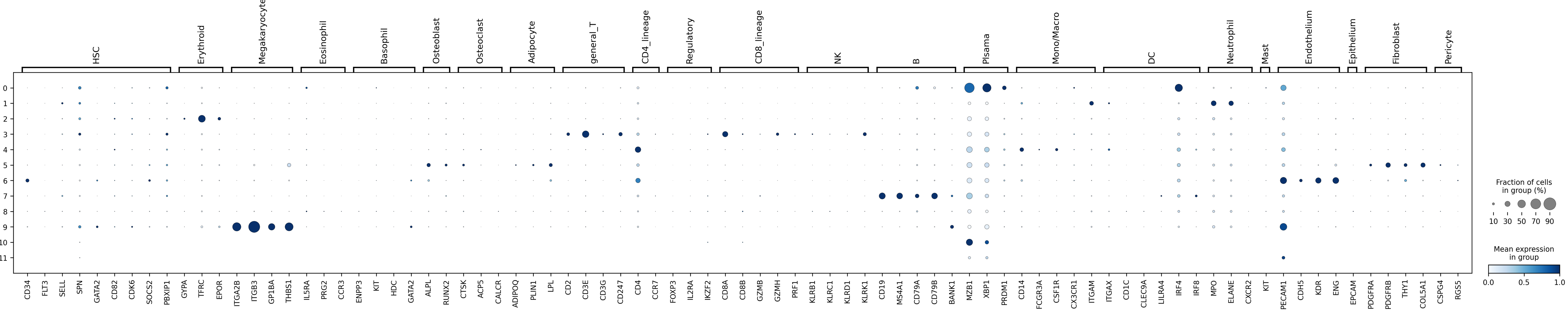

### Supplementary Figure 2.pdf

#### Supplementary Figure 2

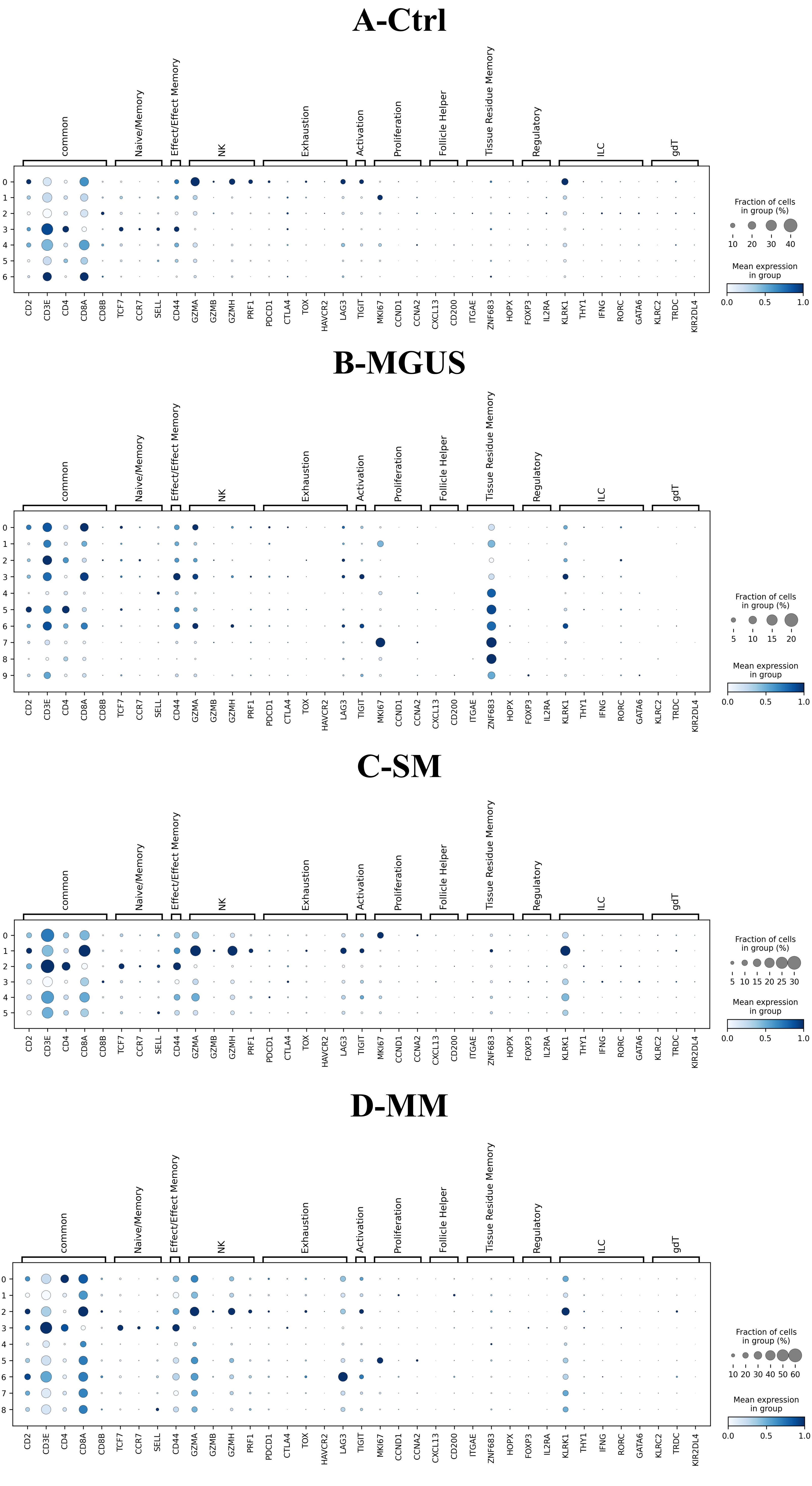

### Supplementary Figure 3.pdf

#### Supplementary Figure 3

##### A-Ctrl

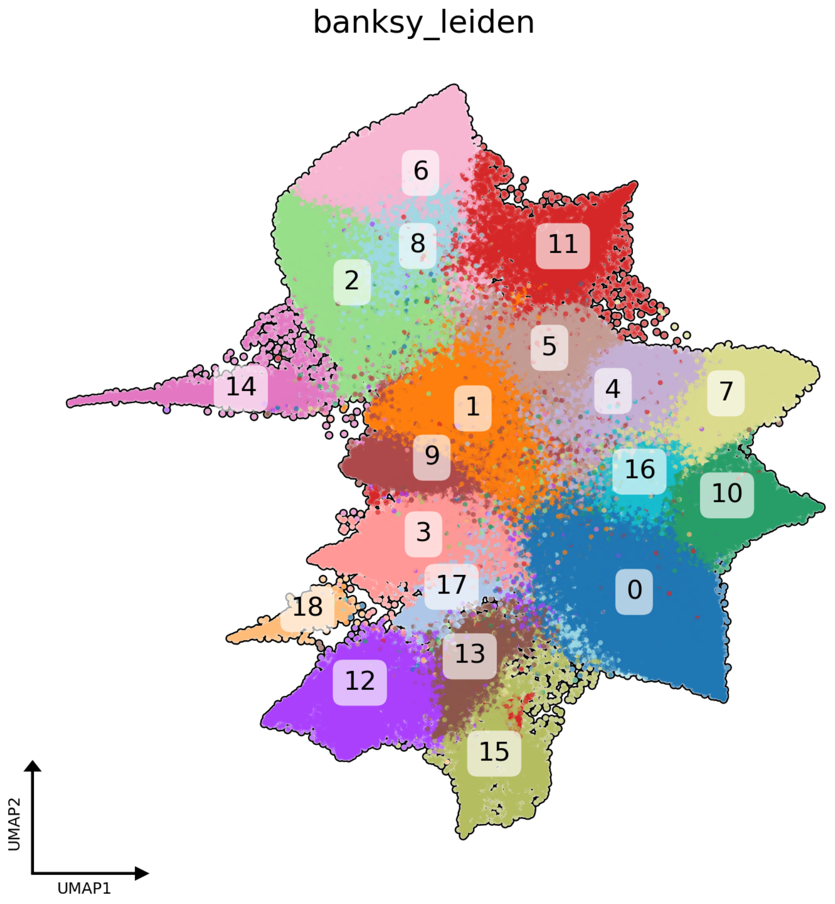


##### B-MGUS

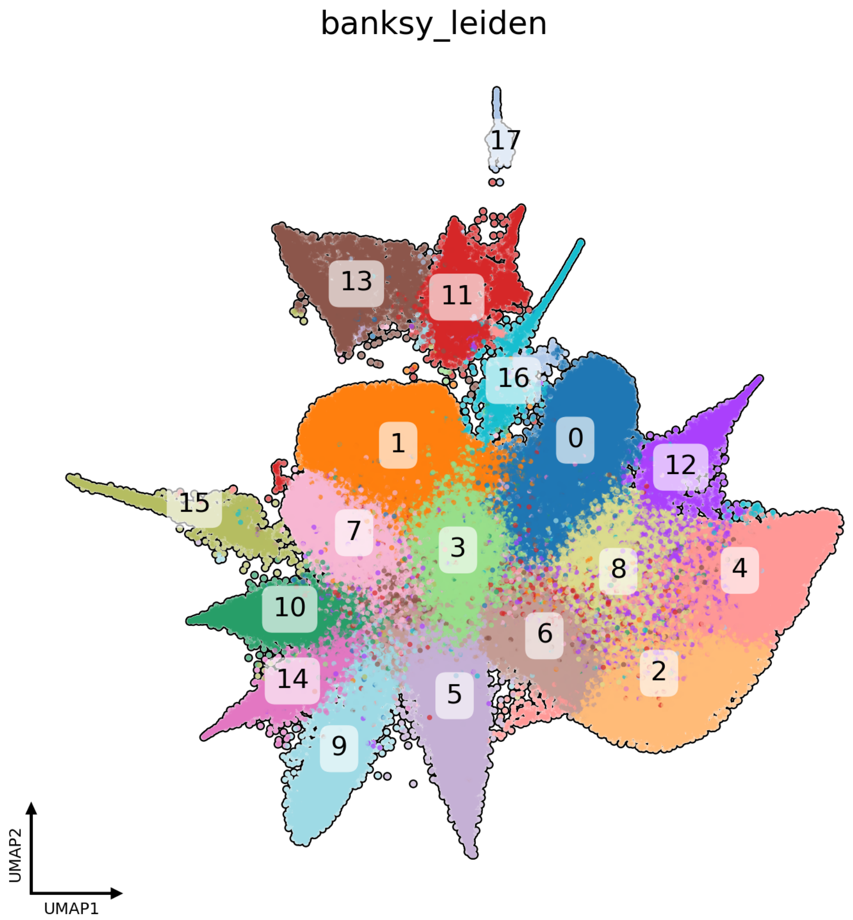


## C-SM

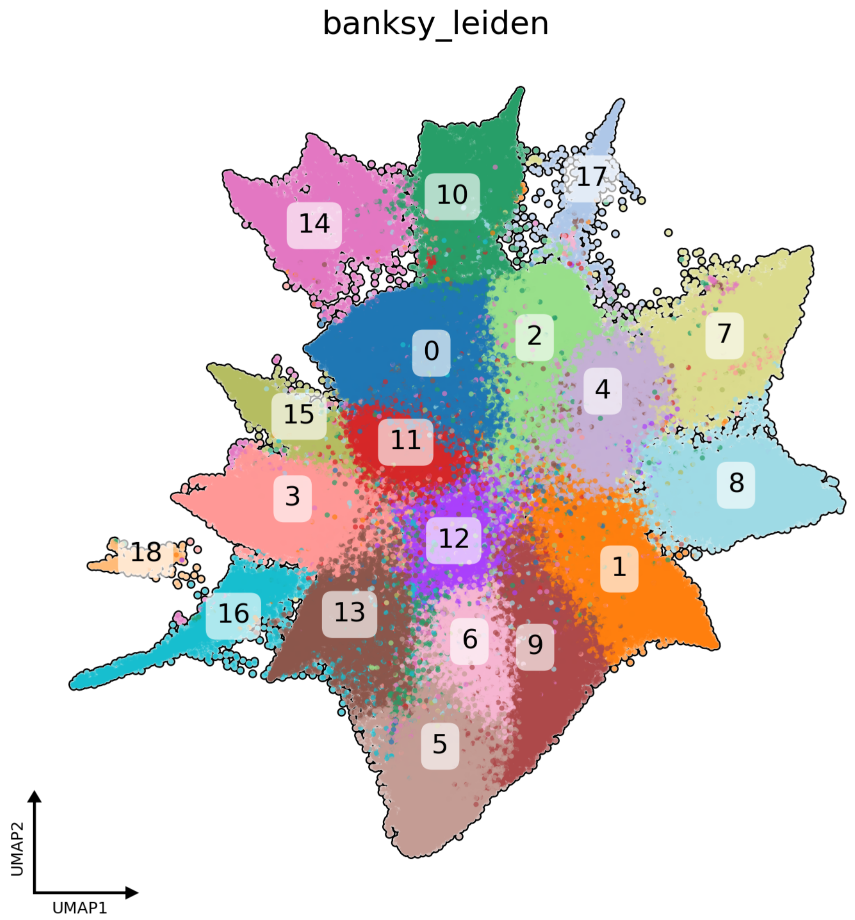


## D-MM

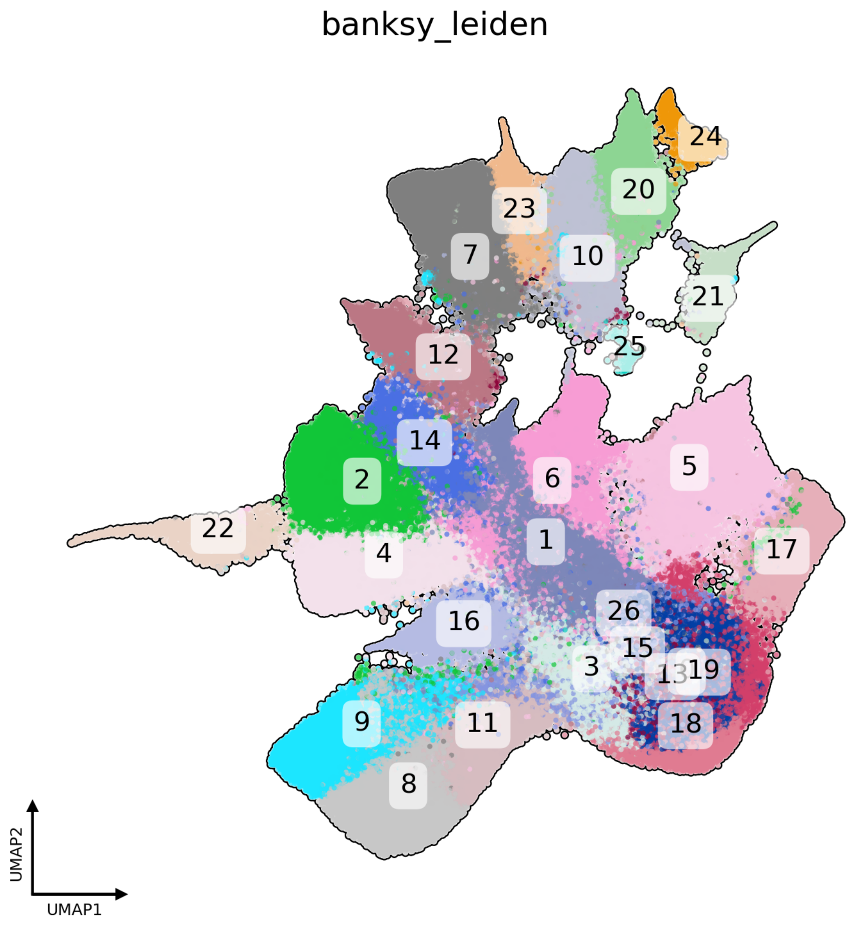

### Supplementary Figure 4.pdf

#### Supplementary Figure 4

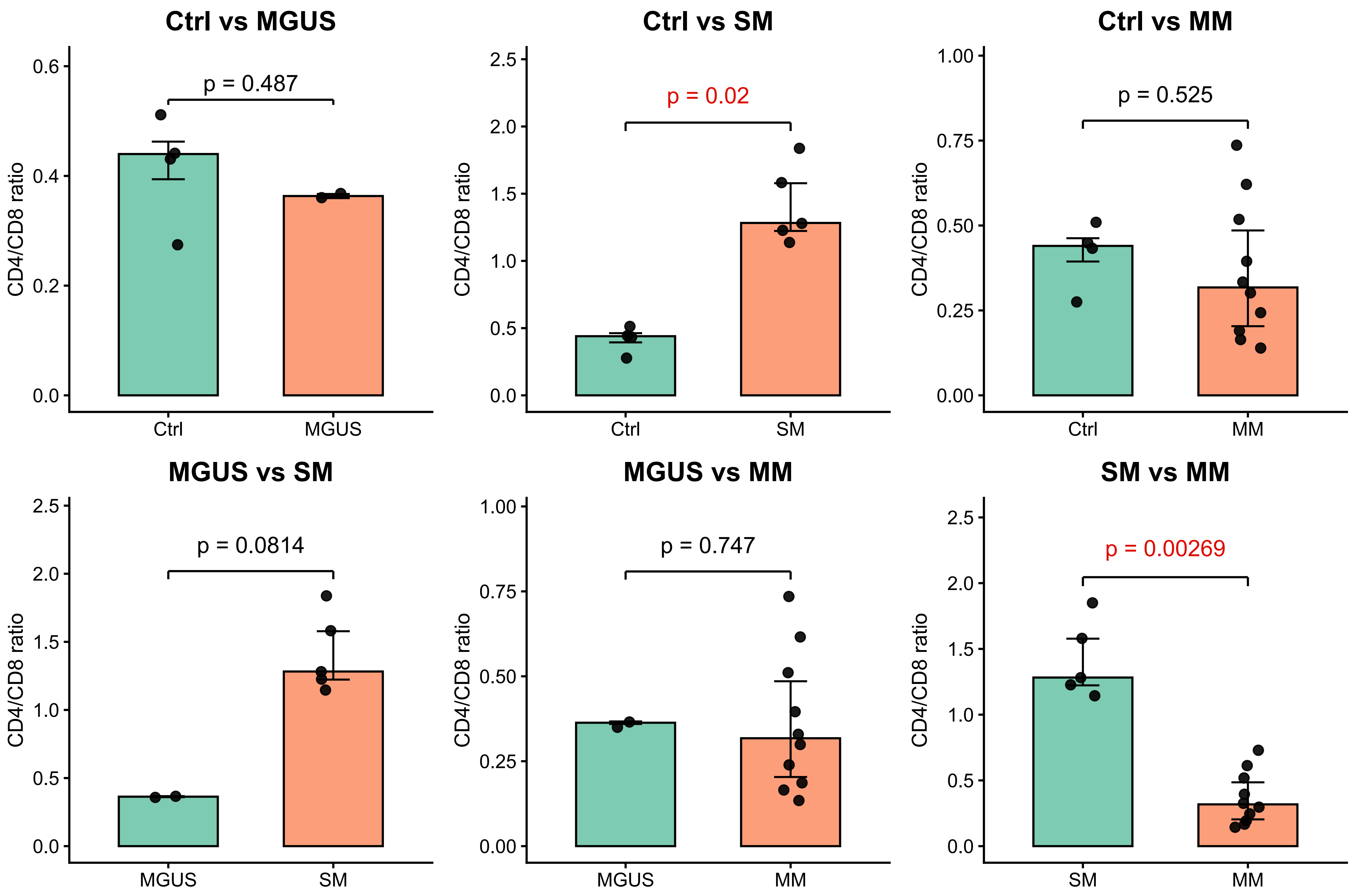

### Supplementary Figure 5.pdf

#### Supplementary Figure 5

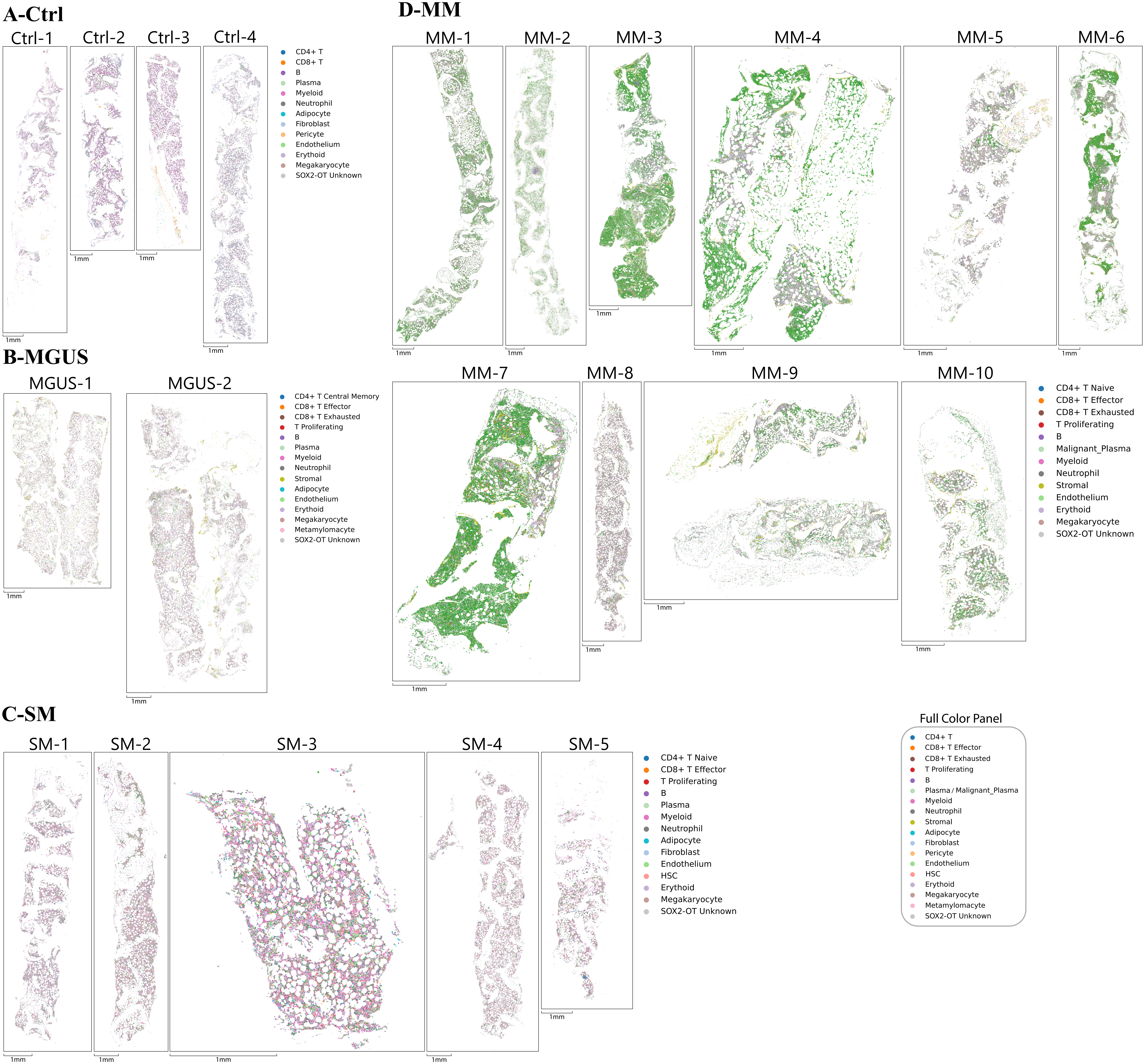

### Supplementary Figure 6.pdf

##### Supplementary Figure 6

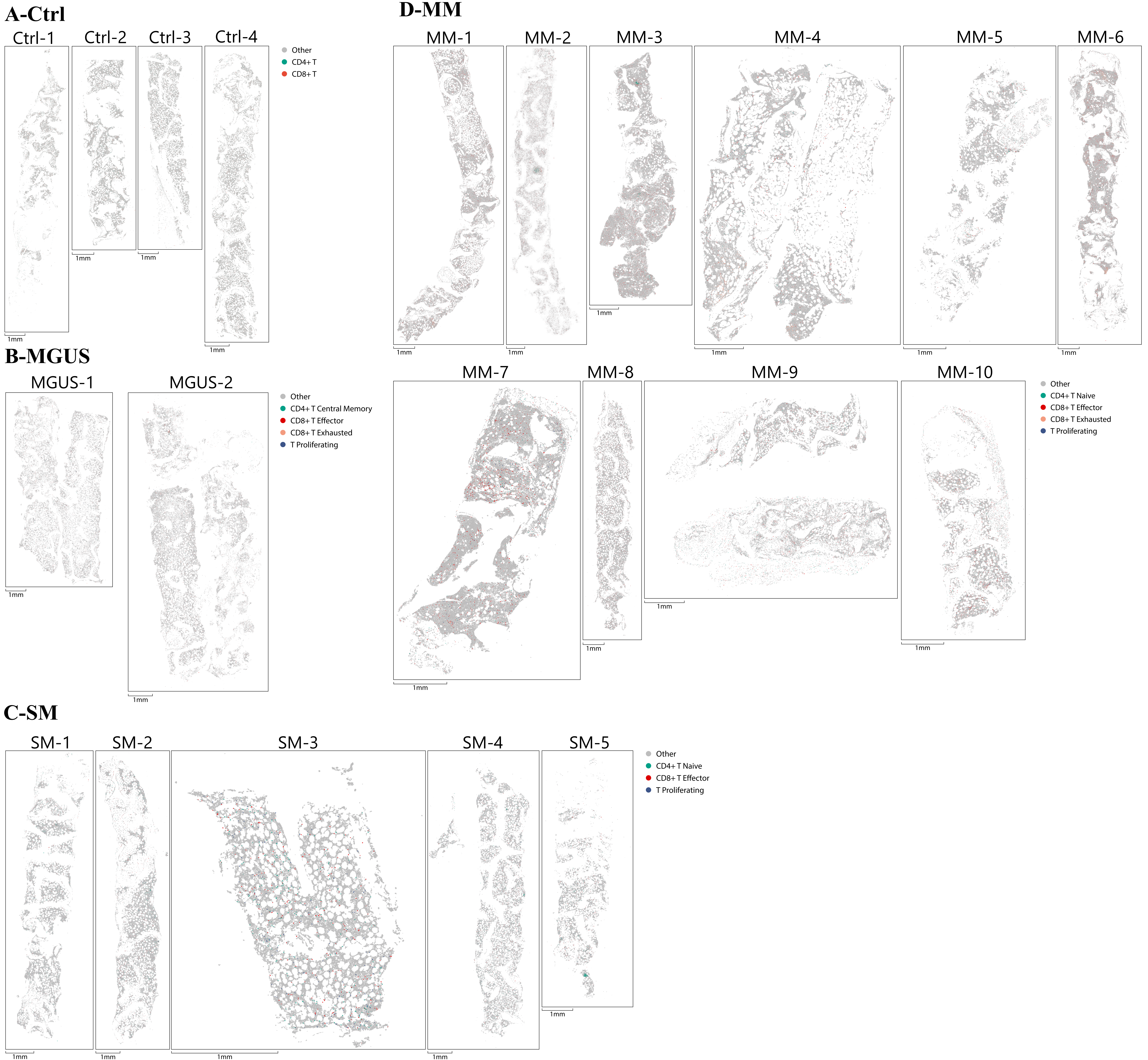

### Supplementary Figure 7.pdf

#### Supplementary Figure 7

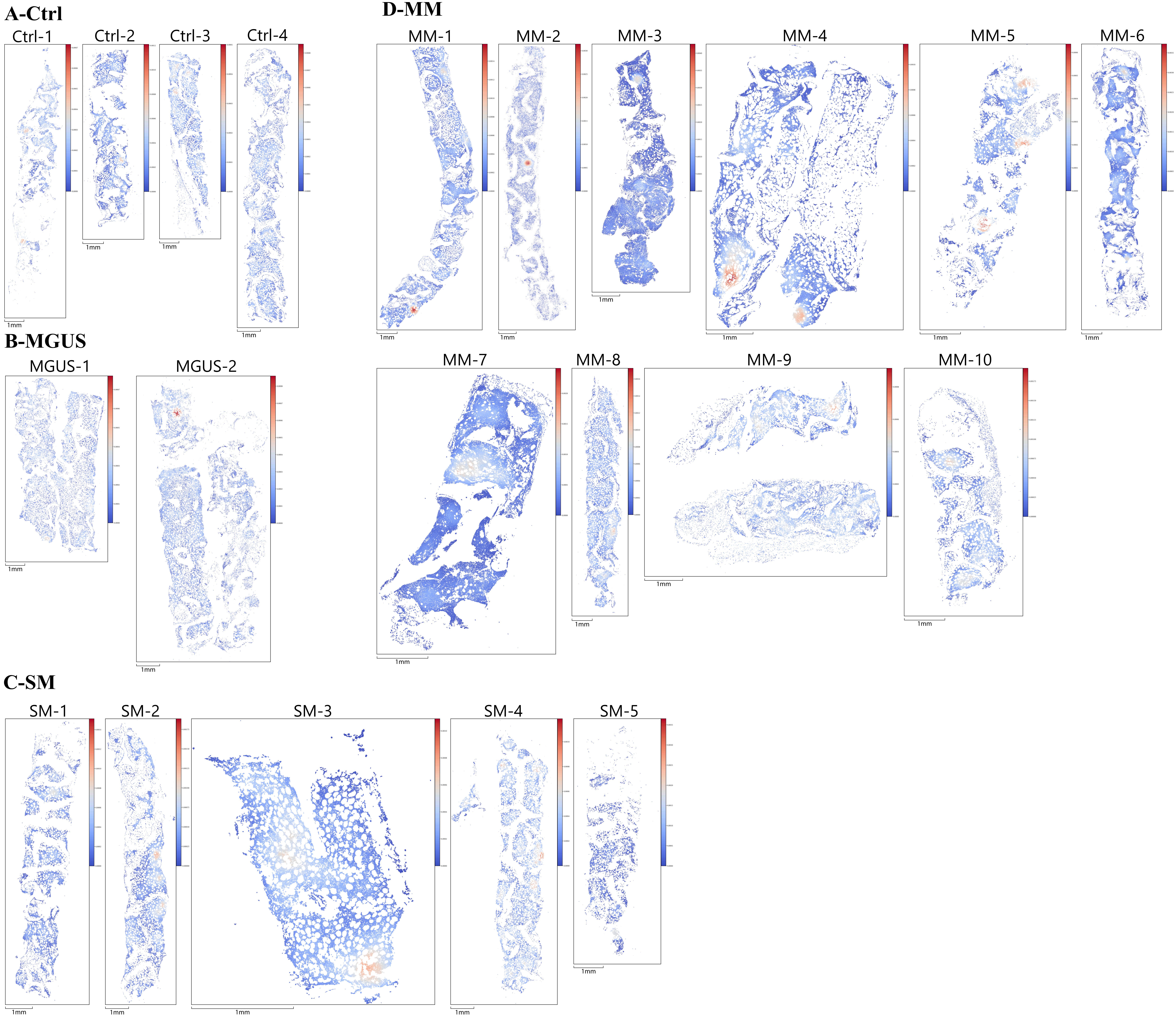

### Supplementary Figure 8.pdf

#### Supplementary Figure 8

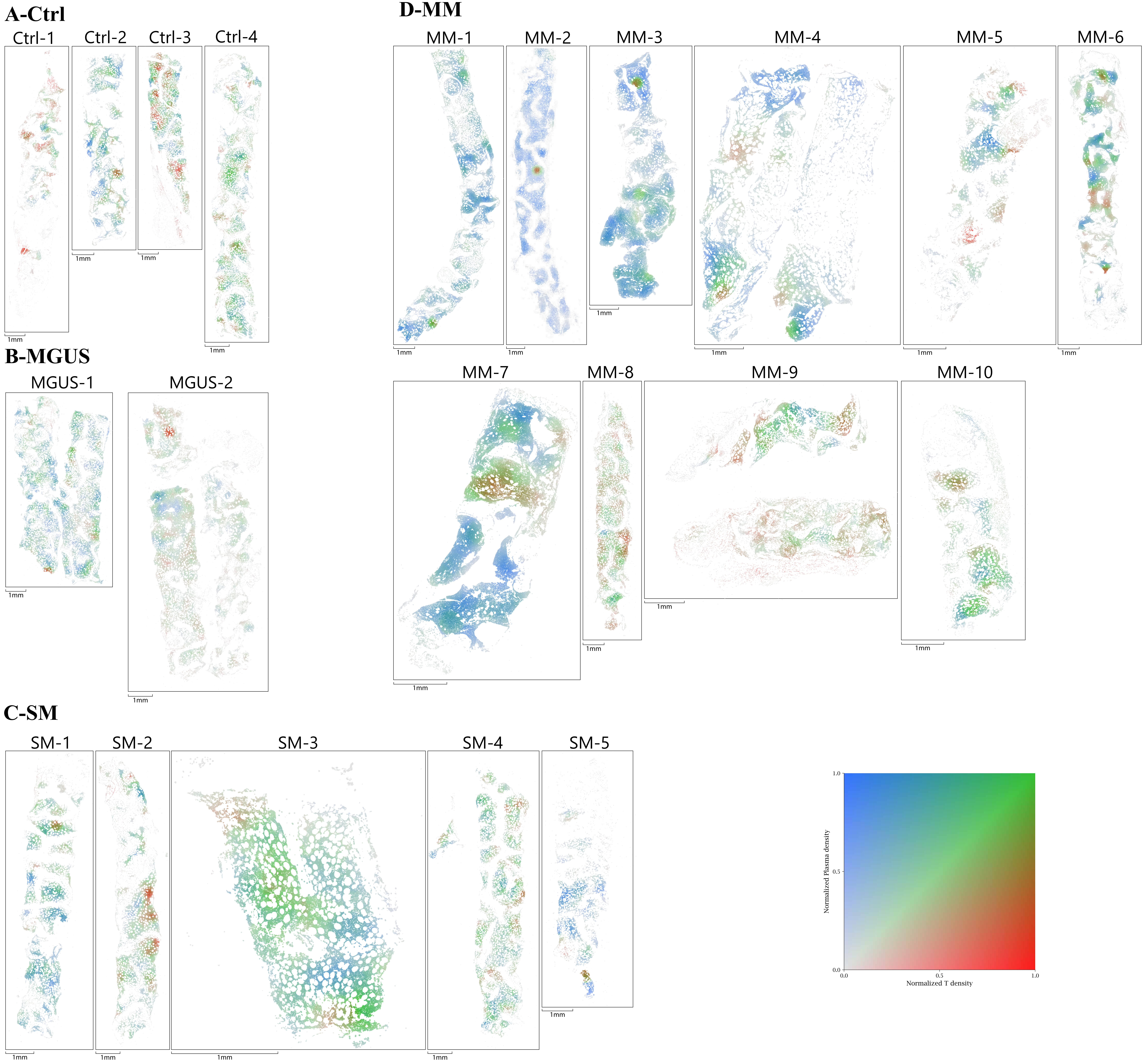

### Supplementary Figure 9.pdf

Supplementary Figure 9

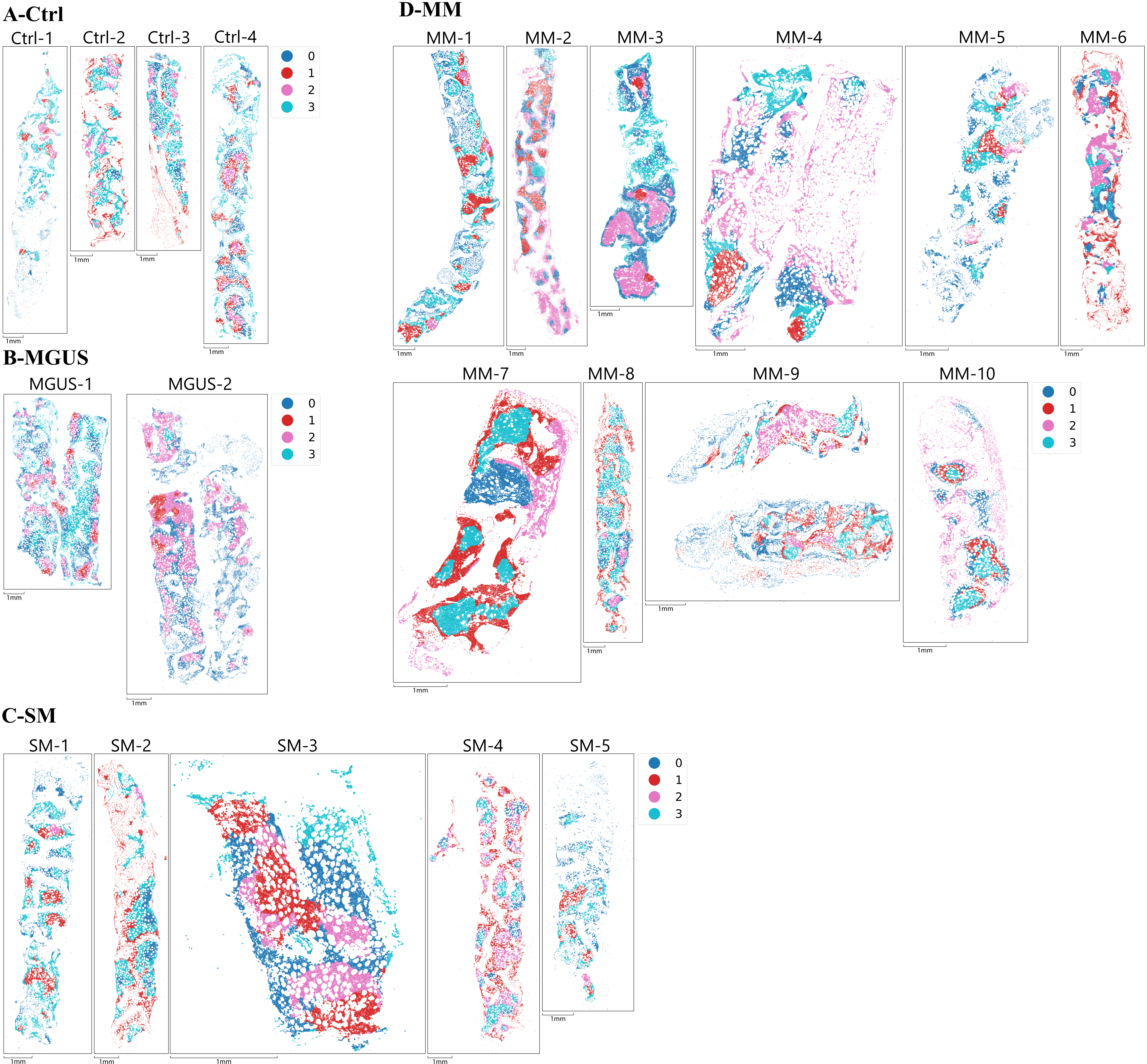

### Supplementary Figure 10.pdf

##### Supplementary Figure 10

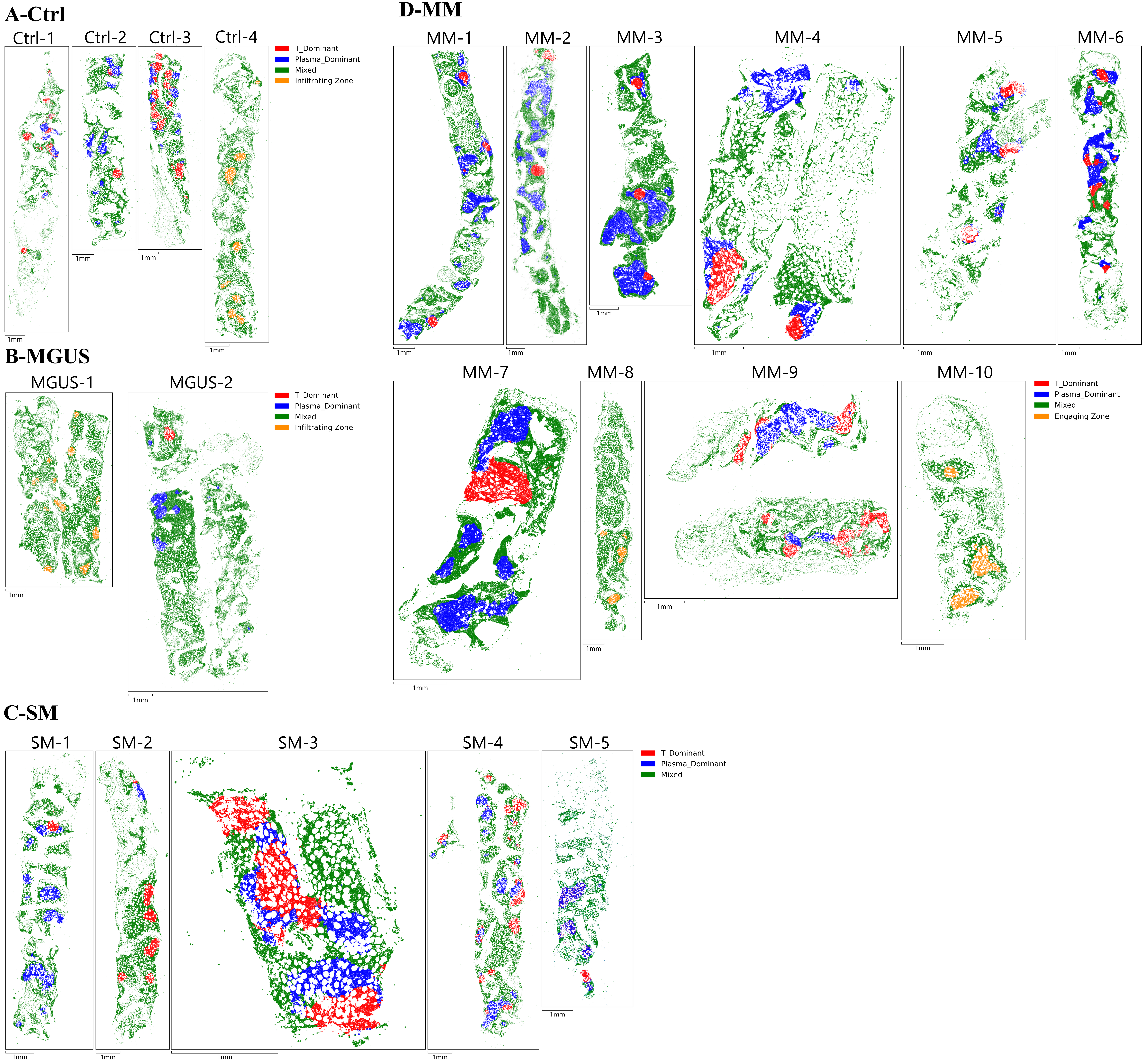

### Supplementary Figure 11.pdf

#### Supplementary Figure 11

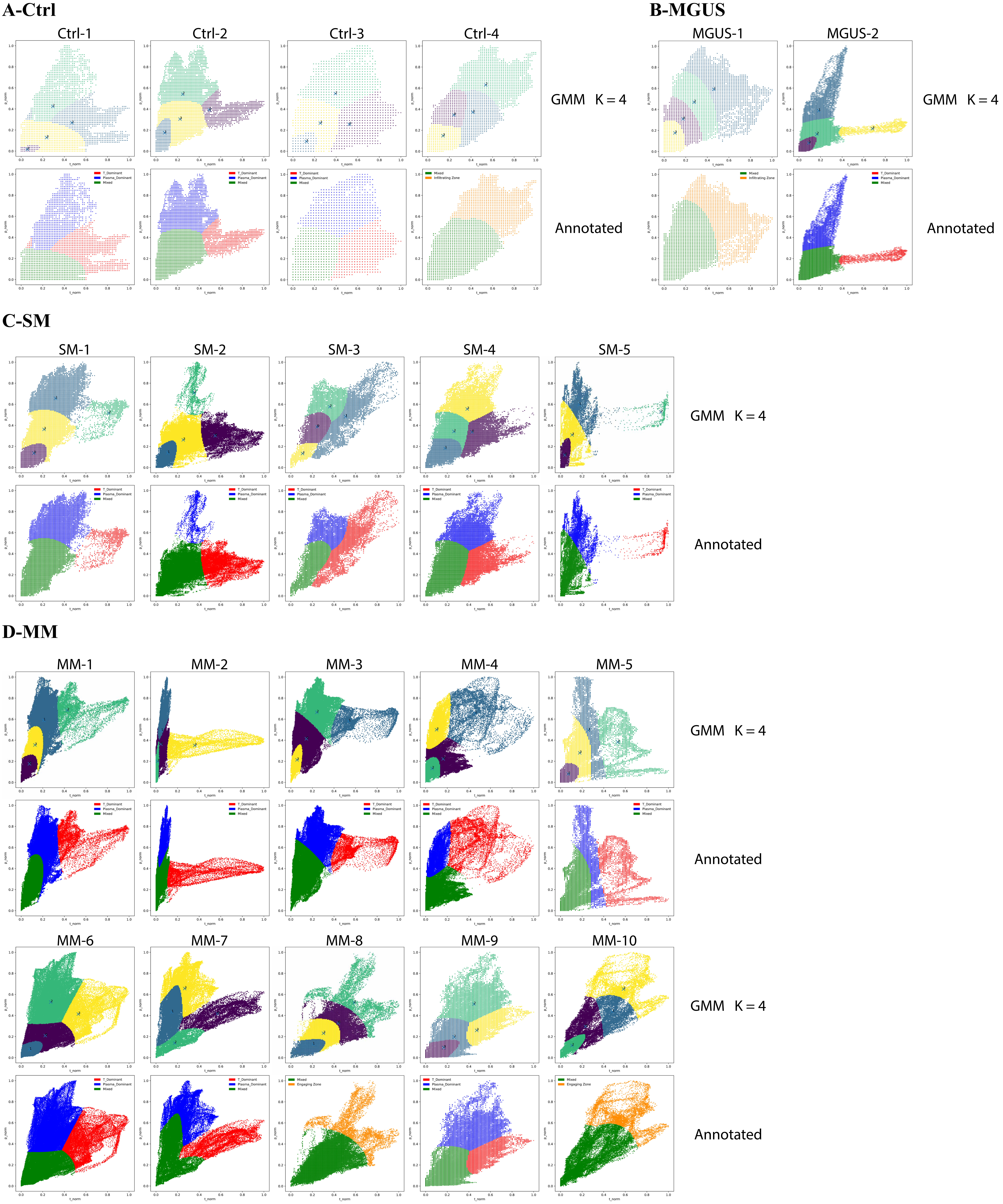

### Supplementary Figure 12.pdf

#### Supplementary Figure 12

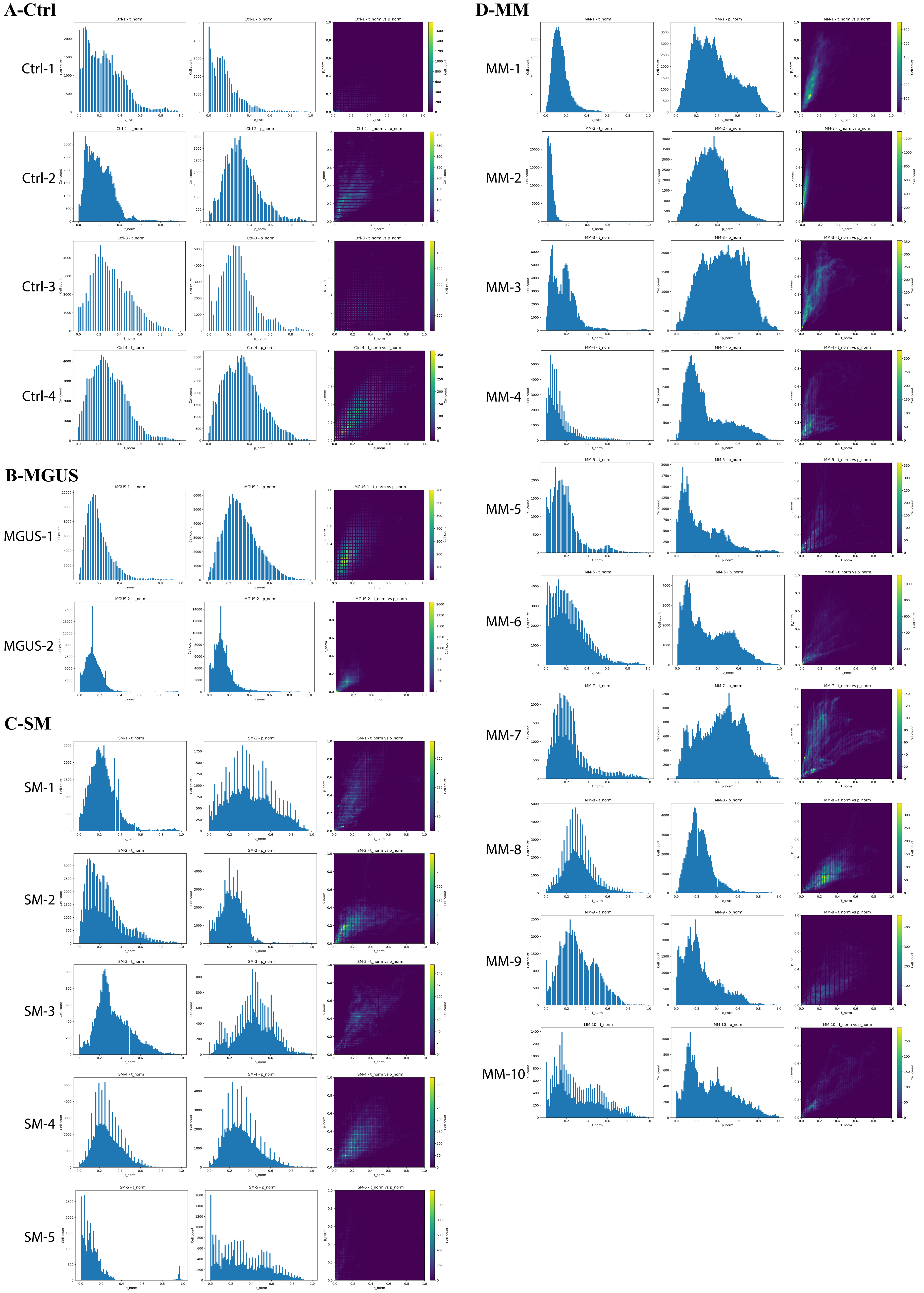

### Supplementary Figure 13.pdf

#### Supplementary Figure 13

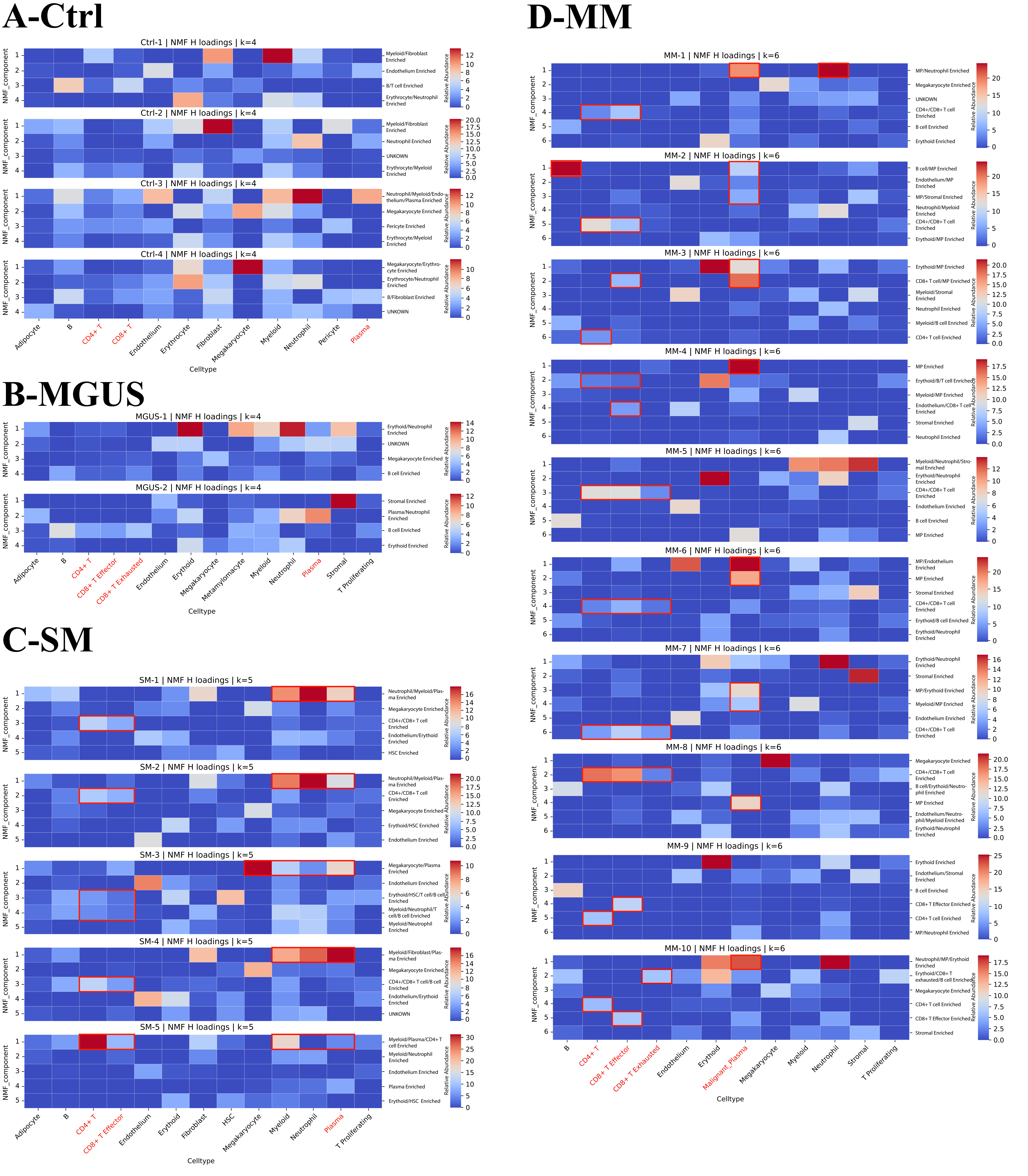

### Supplementary Figure 14.pdf

#### Supplementary Figure 14

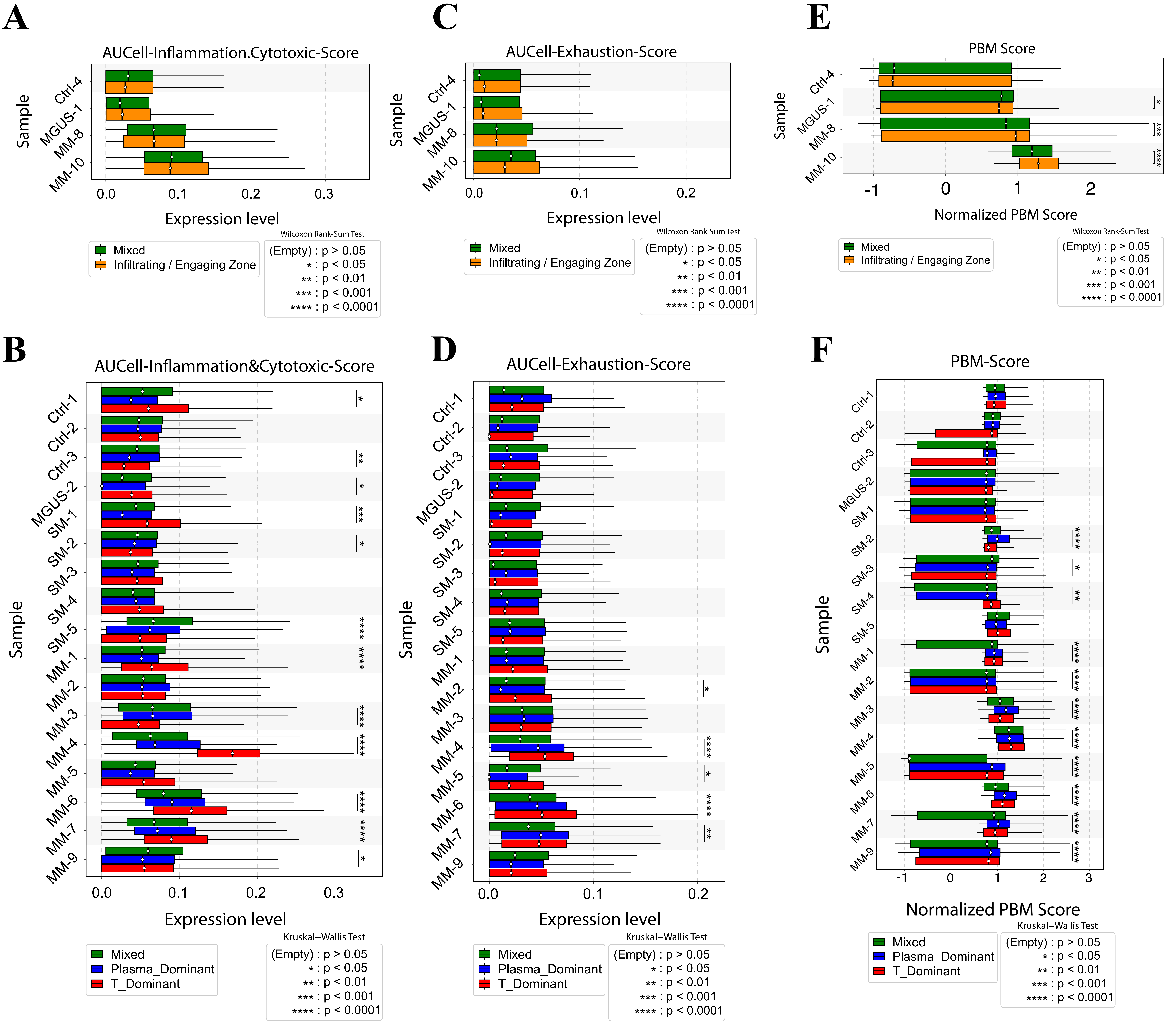

### Supplementary Figure 15.pdf

#### Supplementary Figure 15

# A

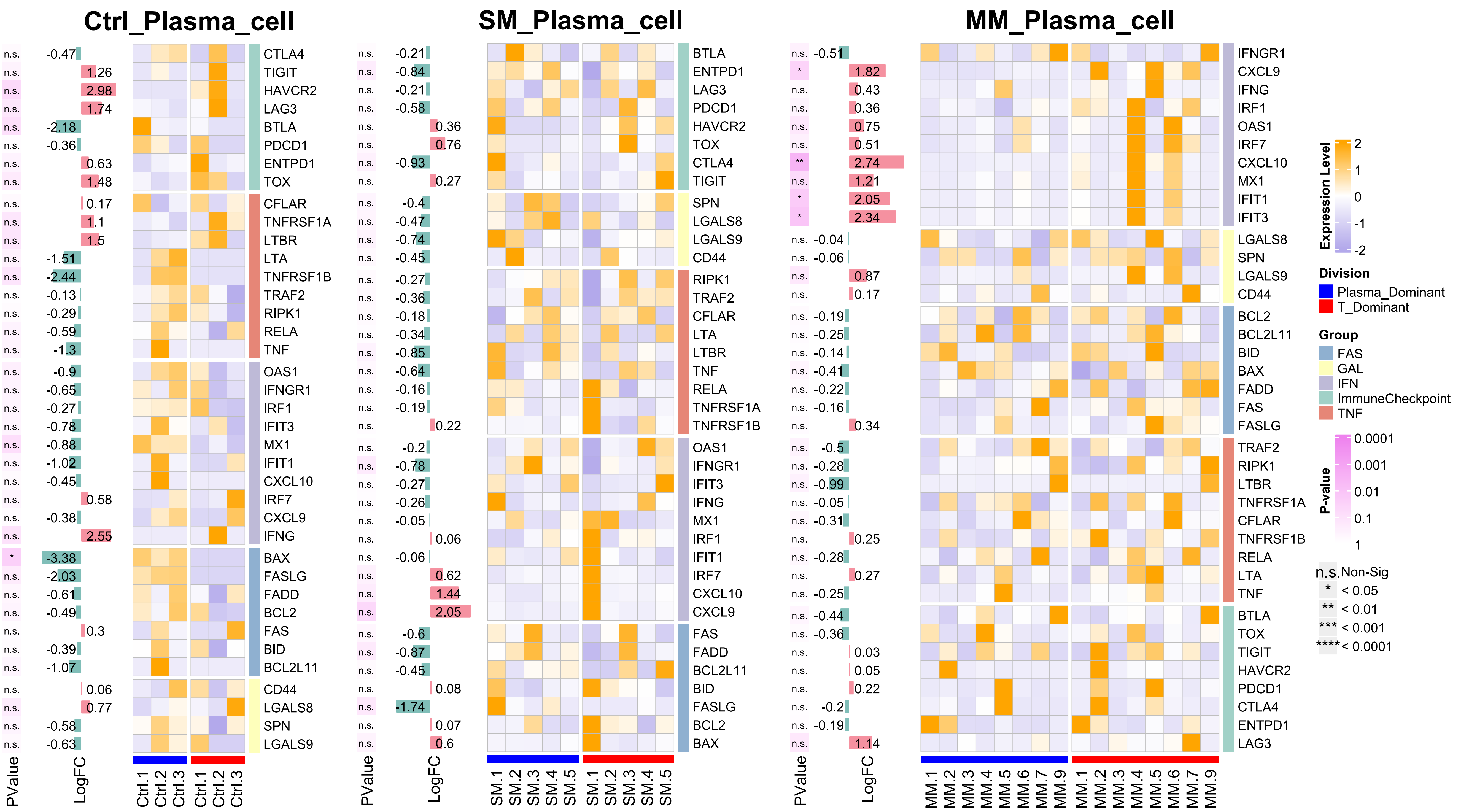


# B

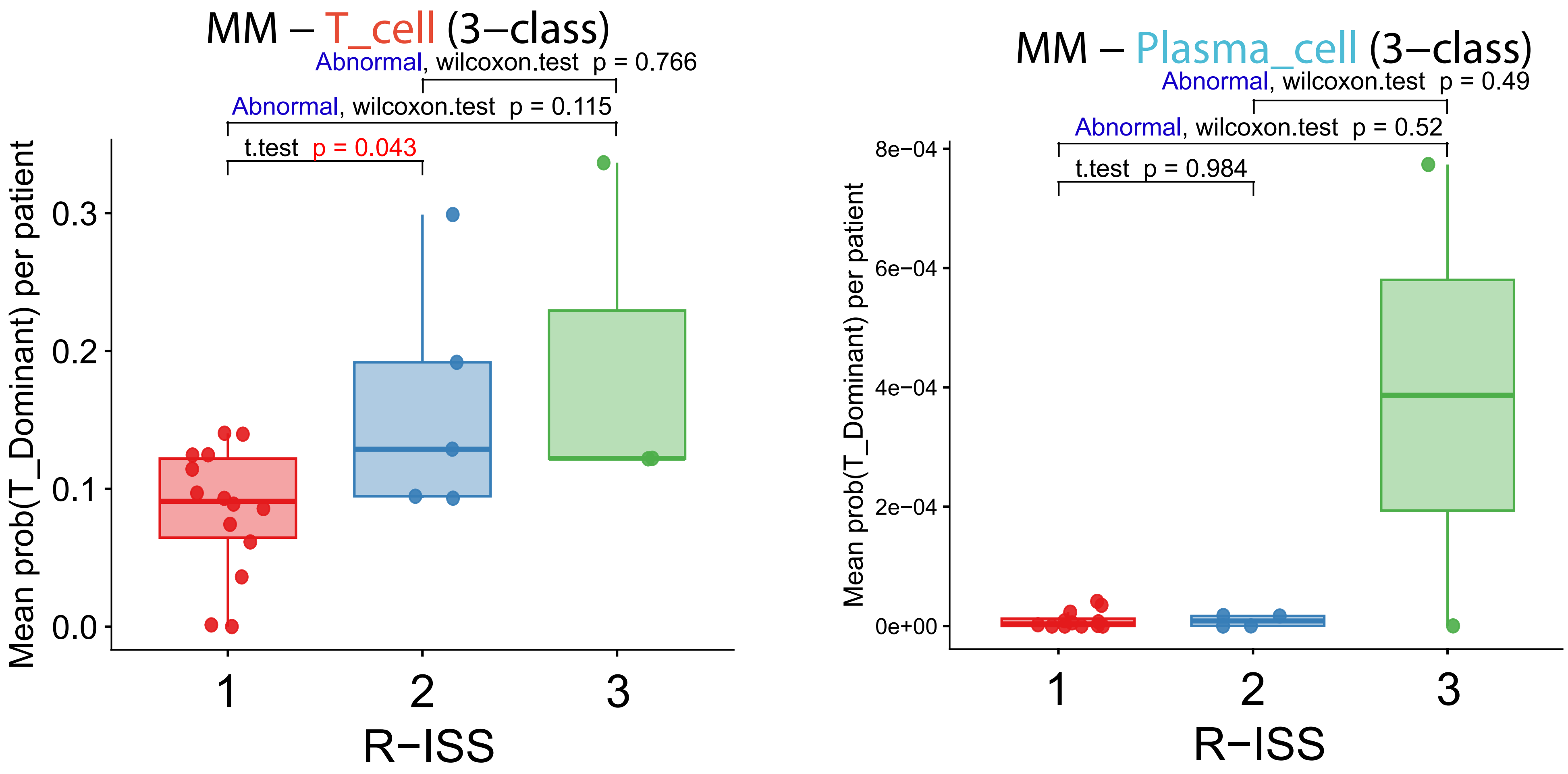
