## Supplementary File 5 for "Spatial transcriptomics and machine learning define exhaustion-like bone marrow T-cell islands associated with myeloma progression and clinical risk"

### Xenium In Situ Grant Application Resource

High-performance imaging-based spatial transcriptomics

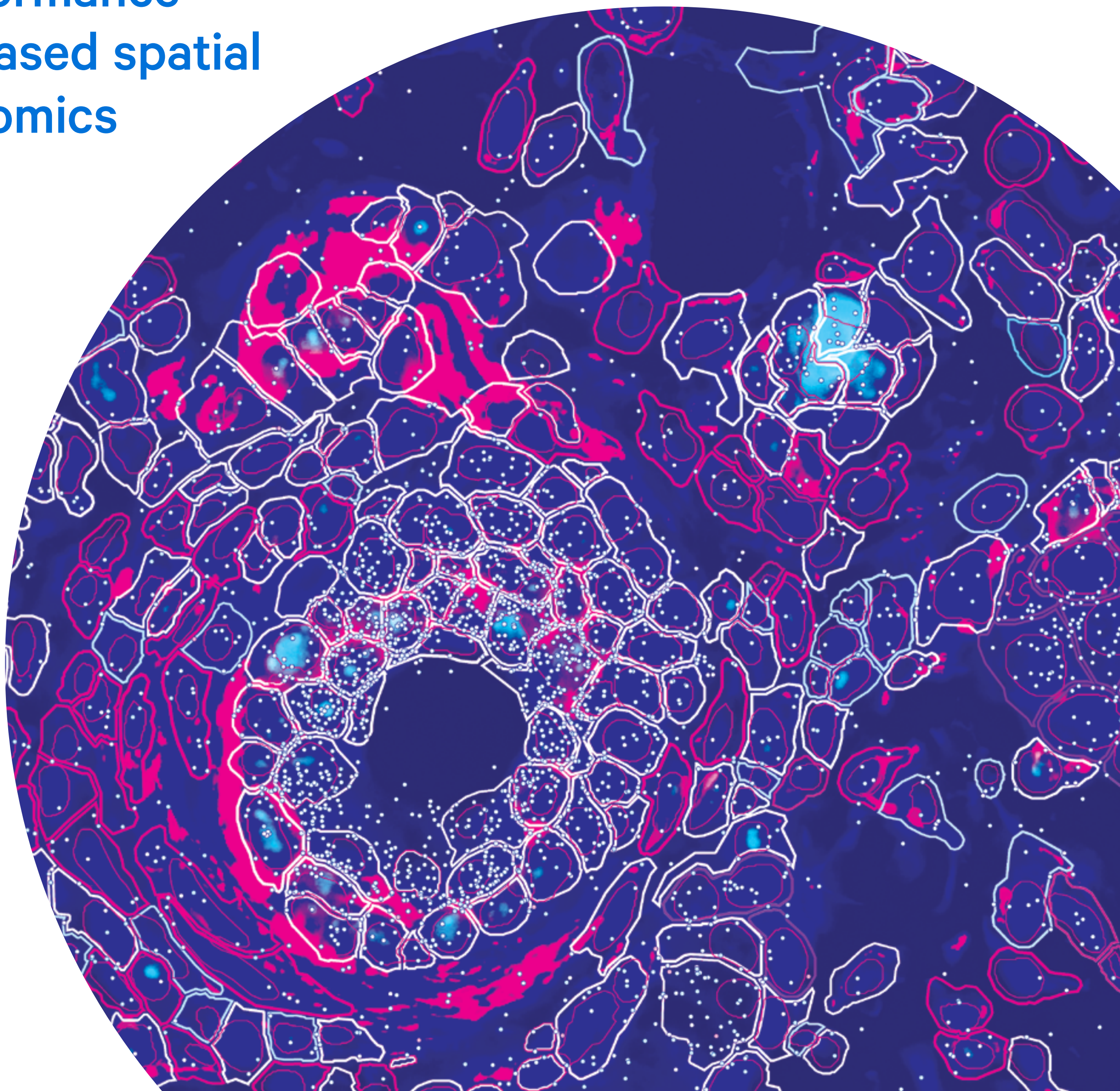

10xgenomics.com

### Xenium In Situ

#### Summary statement

The Xenium platform (Xenium) is an end-to-end solution (hereafter referred to as "Xenium") from 10x Genomics that enables high-throughput subcellular mapping of up to 5,000 genes alongside multiplexed protein in the same tissue section. Compatible with fresh frozen (FF) and formalin-fixed, paraffin-embedded (FFPE) tissues, the platform includes the Xenium Analyzer, a versatile and easy-to-use instrument; sensitive and specific chemistry; multimodal cell segmentation; a diverse menu of customizable panels; and software. Data can be visualized with the intuitive Xenium Explorer or community-developed analysis tools. As with other 10x Genomics products, the Xenium platform has a robust roadmap to enhance the core platform with more capabilities and analytes.

#### Overview

The advancements of single cell spatial imaging analysis provided by Xenium give scientists the capability to view their samples with subcellular resolution and at a depth like never before, by simultaneously profiling up to 5,000 genes and up to 27 proteins, each in the context of their spatial localization. This type of analysis enables studies to not only locate and type cells within their biological context, but also address questions about cell–cell communication, profile cellular microenvironments, and identify rare cell infiltration. Therefore, Xenium In Situ provides another powerful tool that can be used alongside insights gained from Chromium Single Cell and Visium Spatial Gene Expression.

The Xenium platform is a complete end-to-end solution that enables scientists to visualize, quantify, and analyze gene expression and protein in FF and FFPE-preserved tissue sections immobilized onto a Xenium slide. A diverse menu of validated, customizable, fit-for-purpose gene panels, as well as fully custom panels, are available for highly sensitive profiling of genes of interest. Ready-to-use and fully optmized Xenium Protein subpanels enable the multomic analysis of 27 immune and cancer microenvironment markers alongside <500-gene panels in human FFPE tissues.

The platform includes the Xenium Analyzer, a robust, versatile instrument for high-throughput analysis, that comes with onboard analysis capabilities to process image data, localize RNA and protein signals, and perform secondary analysis. You can also easily transfer data off the instrument to perform visualization and further analysis with Xenium Explorer, Xenium Ranger, and/or third-party tools of your choice. Additionally, researchers have access to 10x Genomics technical experts who can provide support through scientific and technical consultations, workflow optimization, and methodology troubleshooting.

### Xenium In Situ platform

##### Sample preparation

The Xenium workflow starts with sectioning tissues onto a Xenium slide (Figure 1). The sections are then treated to preserve and access the RNA with circularizable DNA probes. The DNA probes are flanked by two regions that independently hybridize to the target RNA and also contain a gene-specific barcode sequence. Ligation of the probe ends to each other then generates a circular DNA probe which is enzymatically amplified. In the event that one part of the probe experiences off-target binding, ligation will not occur, thus suppressing off-target signals and ensuring high specificity. If mutimodal cell segmentation is being used, the tissue will then be stained with interior, nuclear, and membrane stains.

Integrated multiplexed protein detection leverages DNA-barcoded antibodies for labeling of protein targets in the same tissue section as RNA, enabling same-cell multiomic analyses of your samples.

The Xenium workflow is also non-destructive. With the sample morphology intact at the end of the workflow, both hematoxylin & eosin (H&E) and immunofluorescence (IF) staining are possible on the same tissue section, enabling the direct comparison of Xenium transcriptional data to morphological data.

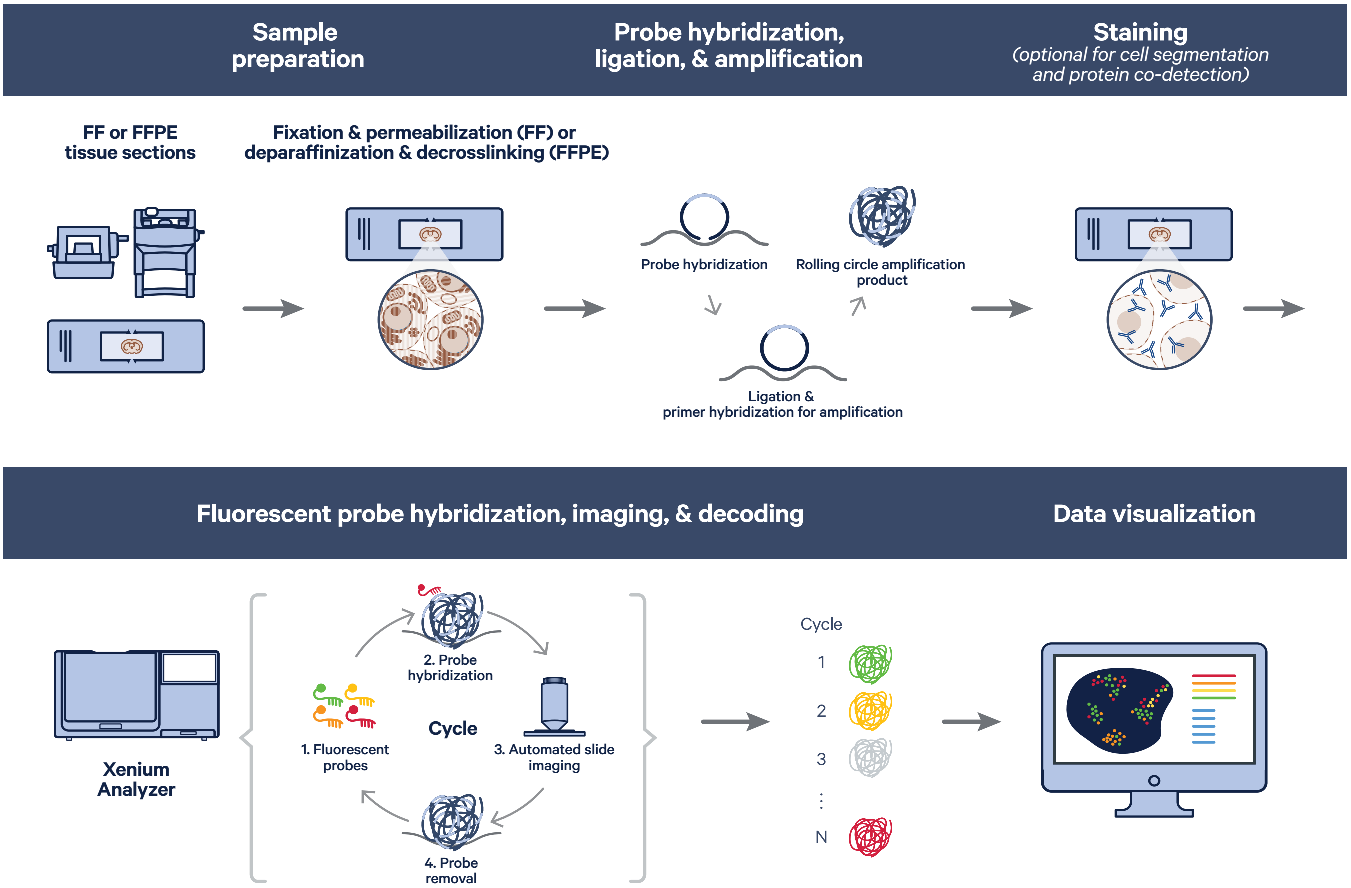


Figure 1. Workflow overview for Xenium In Situ Gene Expression assays.

#### Panels

The Xenium platform uses targeted panels to detect gene and protein expression at the subcellular level, and has been built for the flexibility needed to address different research questions in a variety of samples. Multiple panel options are available so you can find the one to address your specific research needs.

##### RNA panels

10x Genomics offers a menu of pre-designed panels featuring validated, biologically relevant targets. Some pre-designed panels focus on addressing a specific research need, while multi-tissue panels are able to perform cell typing across multiple tissue types. Currently available Xenium panels range from several hundred to 5,000 genes and can be customized with up to 100 additional genes specific to your project.

10x Genomics also offers fully custom panels for human or mouse samples. These panels offer researchers the complete flexibility of selecting up to 480 genes. These are designed using the same algorithm as the pre-designed panels.

Probes for isoforms, gene fusions, viral or bacterial sequences, protein tags, and fluorescent reporters can be developed with our Advanced Custom Panel Design option.

##### Protein panels

10x Genomics has developed and validated ready-to-use Xenium Protein subpanels for exploration of immune and tumor microenvironments. Comprised of 6 subpanels containing 3–4 markers each, the modular subpanels can be mixed and matched according to the researcher's preference. Compatible with <500-gene panels and human FFPE tissues, Xenium Protein subpanels enable same-cell multiomics of up to 27 proteins, including 4 structural markers, alongside 100’s of RNAs.

#### Decoding on Xenium Analyzer

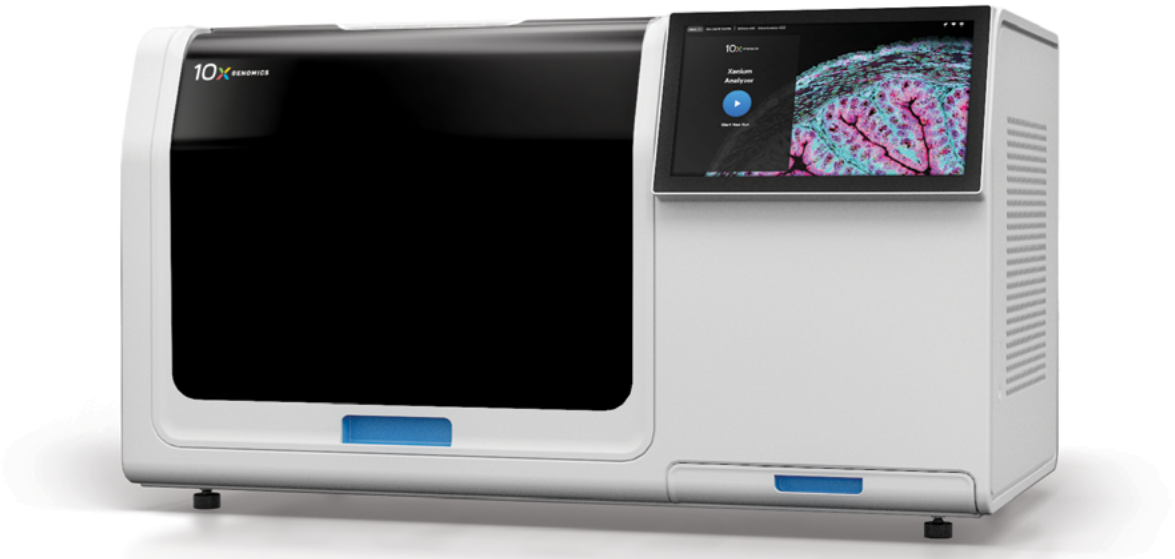

Figure 2. Xenium Analyzer instrument.

The Xenium Analyzer (Figure 2) fully automates and integrates sample handling, liquid handling, and wide-field epifluorescence imaging. Data processing is performed in parallel to a run, allowing data outputs to be prepared quickly for interpretation and making them available for analysis as soon as a run completes. The imager uses a high numerical aperture and a fast area scan camera with a low read noise sensor to achieve ~200 nm-per-pixel resolution. The imager's field of view is 600 x 720 µm, and the Z stacks are acquired with a 0.75-µm step size across the whole tissue thickness. The standard tissue thickness is typically 5 µm for FFPE sections and 10 µm for FF sections.

The acquired images are processed through the Xenium Analyzer's onboard analysis software to enable single-molecule localization with a lateral (XY-axis) precision within <30 nm and about <100 nm z-axis precision. Up to two slides, each with an imageable area of 12 x 24 mm, can be loaded onto the Xenium Analyzer per run. This large imageable area allows the flexibility to place multiple sections and samples on each slide.

Analyte detection and image acquisition are performed on the Xenium Analyzer in cycles. The reagents, including fluorescently labeled probes for the detection of RNA and protein molecules, are automatically cycled in, incubated, imaged, and removed by the instrument. An optical signature specific to each RNA transcript within the sample is generated, enabling identification of the target gene. Proteins are identified using DNA-barcoded antibodies. At the end of the instrument run, the data is integrated to build a spatial map of the transcripts and proteins across the tissue section.

##### Software—data management and analysis

Xenium analysis software allows for immediate exploration of the assay's subcellular readout. This functionality is delivered in two parts: (1) comprehensive onboard analysis and (2) off-instrument analysis that leverages the exploration-ready output generated by the Xenium Analyzer (Figure 3).

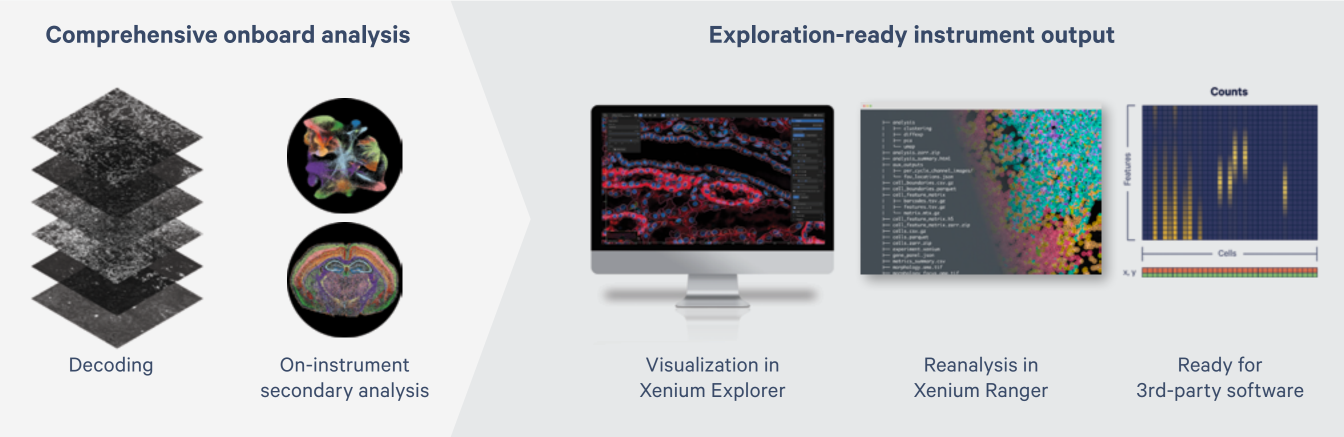


**Figure 3. Xenium's streamlined onboard and off-instrument data analysis.**

The first step of Xenium onboard analysis is the decoding of optical signatures into transcripts. As described in the "Decoding on Xenium Analyzer" section, chemistry and imaging cycles consist of fluorescent oligos hybridizing to amplified products. In successive cycles, fluorescent oligos bind to rolling circle amplification products and images are acquired. It is the unique optical signature of a transcript across cycles—as it lights up in different channels—that allows the onboard image processing to decode and determine transcript localization across the tissue. The next step of Xenium on-board analysis is cell segmentation. The tissue is stained with a single mix containing multiple stains targeting nuclear, cell interior, and cell boundary targets. Then, a purpose-built AI algorithm uses morphological features for precise, data-driven cell segmentation. Xenium onboard analysis then leverages the morphological information provided by these stains to perform precise, data-driven cell segmentation. The final step of the on-instrument analysis workflow is assignment of transcripts and proteins to cells and computation of additional secondary analyses, including clustering, dimensionality reduction, and differential expression.

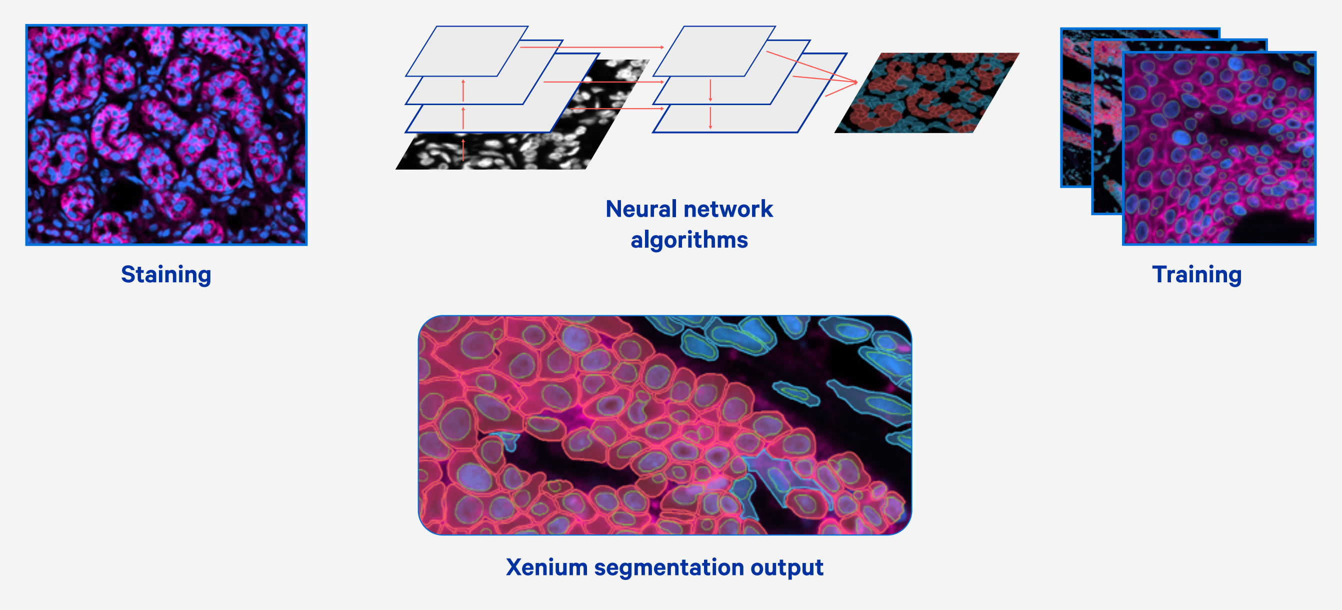


**Figure 4. Xenium's integrated, deep learning approach to cell segmentation.**

Immediately following an experiment run, the Xenium Analyzer output can be easily transferred off instrument for interactive visualization using the Xenium Explorer desktop application or community-developed tools (Figure 3). Xenium Explorer is a data visualization application for exploring, QCing, and analyzing Xenium In Situ data. It is designed to allow map-like panning and zooming across high-resolution morphology images, cells, and transcript and protein data, offering easy navigation of gene lists and cell groups to explore spatial- and cell-type-specific patterns of gene and protein expression in addition to facilitating tissue-scale comparisons.

Finally, our Xenium Ranger analysis pipelines offer you increased flexibility in analyzing your data by enabling you to relabel transcripts, resegment cells, and import third-party cell segmentation to reassign transcripts and visualize results in Xenium Explorer.

##### Pilot studies, training, and support

To help facilitate pilot data for grants and instrument purchase, 10x Genomics offers the Xenium Catalyst program. Researchers whose projects are selected can submit their samples to a Xenium Catalyst lab, choose any pre-designed Xenium RNA panel or design their own custom panel, and receive high-quality Xenium data for their unique applications.

After purchase and acquisition of the Xenium Analyzer, 10x Genomics provides on-site installation and calibration of the instrument by a qualified Field Service Engineer (FSE). Comprehensive training for users is performed by a trained Field Application Scientist (FAS). Training topics include sample preparation, instrument operation, data interpretation, and data analysis. After completion of training, the customer will receive comprehensive support from our Technical Support, FAS, FSE, and Applied Bioinformatics teams, covering all aspects of the workflow, consumables, instrument, software and analysis. Local support is available in each geographic region.

##### Foundational technology

The Xenium platform is based upon foundational technologies acquired from ReadCoor and Cartana, which stem from developments in the laboratories of George Church and Mats Nilsson, respectively, combined with proprietary developments from 10x Genomics. The resulting proprietary 10x Genomics in situ technology features sensitivity, specificity, and throughput that are improved many-fold over the foundational technologies.

##### Representative data

The following datasets were generated using our Xenium Analyzer in conjunction with a 313-plex human breast tissue panel on an FFPE-preserved human breast invasive carcinoma tissue section and compared with a hematoxylin and eosin (H&E)-stained section from the same block, which had been previously annotated by a pathologist and run through the Visium Spatial Gene Expression for FFPE workflow and analysis pipelines.

##### Xenium data

The analysis of an FFPE-preserved human breast invasive carcinoma resulted in a spatial map of the transcripts with X, Y, and Z coordinates. Nuclei boundaries were identified through DAPI staining, and the cell boundaries were estimated by expanding the nuclei edges. RNA transcripts were assigned to specific cells and a gene-by-cell matrix was generated. Based on each cell's RNA content and the markers making up the panel, cell types were then assigned (Figure 5). For example, *FASN* was used to identify invasive tumor cells, and POSTN was used to identify stromal cells. Cells were colored by their assigned cell types. This resulted in a spatial map of cells, identified by type, with the assigned transcripts.

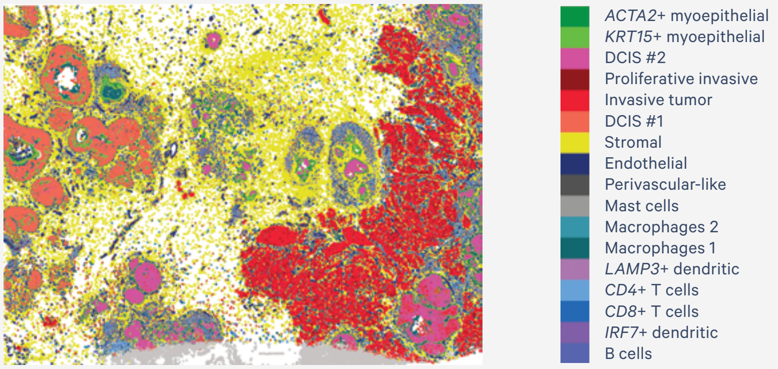


**Figure 5. Xenium data provide extremely high-resolution single cell information with spatial localization from a targeted panel of genes.**

##### Correlation data

To validate that the human breast panel for Xenium accurately captured the biological heterogeneity of the sample, the gene expression data from Xenium was compared to the expected cell types. The expected cell types in the sample were identified using Chromium Gene Expression Flex data generated from the same breast cancer block. When the Chromium Gene Expression Flex data was filtered to the 313 genes used in the Xenium human breast panel, the same cell-type populations were identified, confirming that the Xenium human breast panel captured the biological heterogeneity in the breast cancer sample (Figure 6).

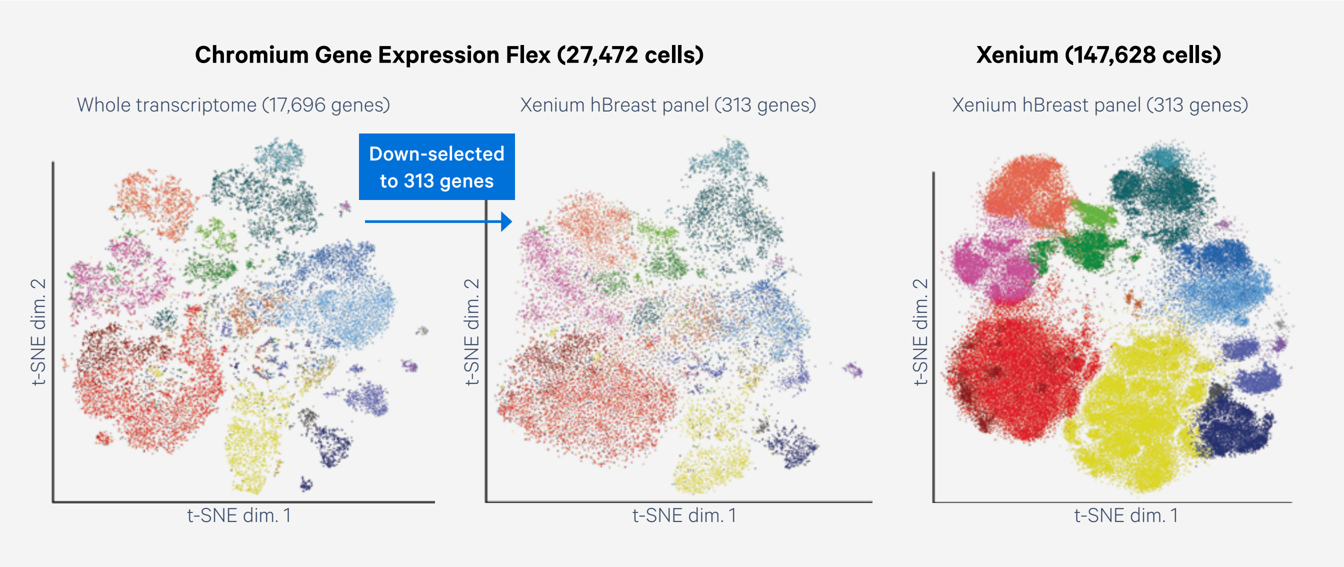


**Figure 6. Xenium captures the full biological heterogeneity observed in Chromium Gene Expression Flex data.**

Xenium also accurately captures spatial localization of transcripts and correlates well with the corresponding morphology as seen in the H&E image (Figure 7). To assess accurate capture of the spatial components of Xenium data, we were able to compare it with an image of the same tissue section H&E-stained post Xenium workflow. These images showed the same morphology. Xenium data also correlates well with the whole transcriptome data generated by Visium. Comparing the expression of three hormone receptor genes between Visium (detected through NGS) and Xenium (detected through microscopy at subcellular resolution) data showed a strong correlation of spatial localization of these transcripts. Further, because of the subcellular resolution of Xenium, only after these three genes were located via Xenium were we able to identify the same patterns in Visium.

Taken together, these data demonstrate that the Xenium platform can reproduce the cellular heterogeneity captured by our Chromium platform, the spatial morphology shown during H&E staining, and the spatial distribution of gene expression observed with our Visium platform—all within the same platform.

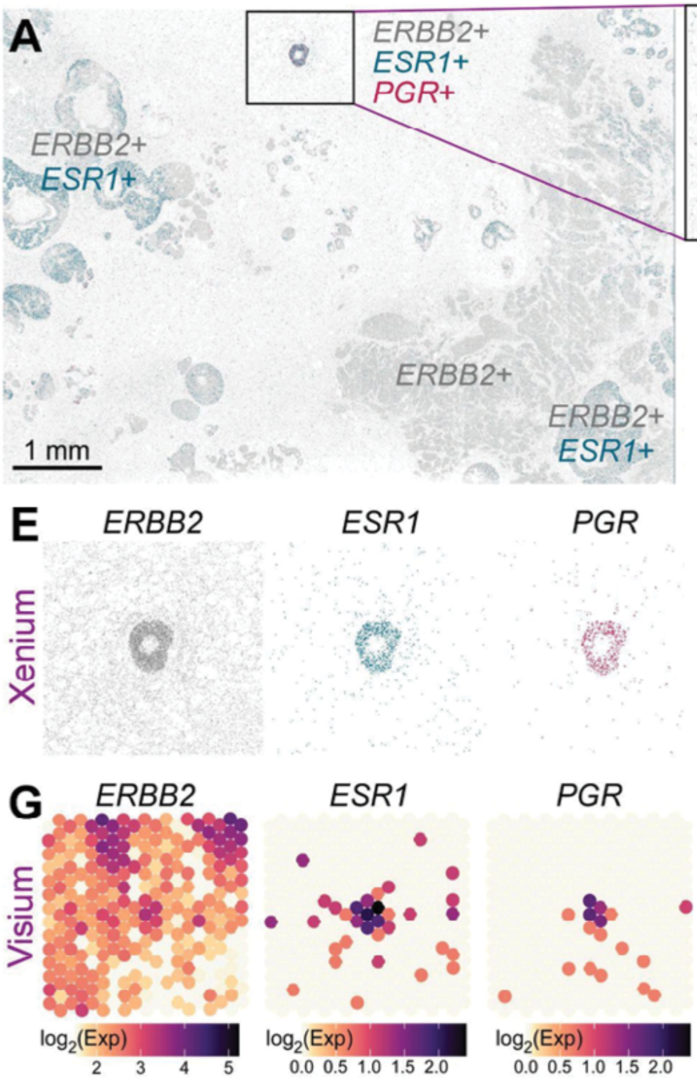


**Figure 7. Xenium reproduces the morphology seen in H&E images and spatial gene expression data obtained with the Visium platform. A.** Xenium spatial plot for *ERBB2* (HER2- gray), *ESR1* (estrogen receptor - green), and *PGR* (progesterone receptor - magenta) decoded transcripts. **B.** Closer view of triple-positive ROI. **C.** Corresponding H&E image. **E.** Individual Xenium spatial plots from (B). **G.** Triple-positive region is identified in Visium data (given a priori knowledge from Xenium).

Finally, to benchmark the sensitivity of the Xenium assay, it was compared against the median gene sensitivity of Chromium Gene Expression Flex. Comparing median gene expression from both technologies ensured no bias from high or low expressors. The number of transcripts per cell for Xenium was compared to the number of UMIs per cell for Chromium Gene Expression Flex. Overall, Xenium was 1.4x more sensitive than 10x Genomics' most sensitive single cell assay, Chromium Gene Expression Flex (Figure 8).

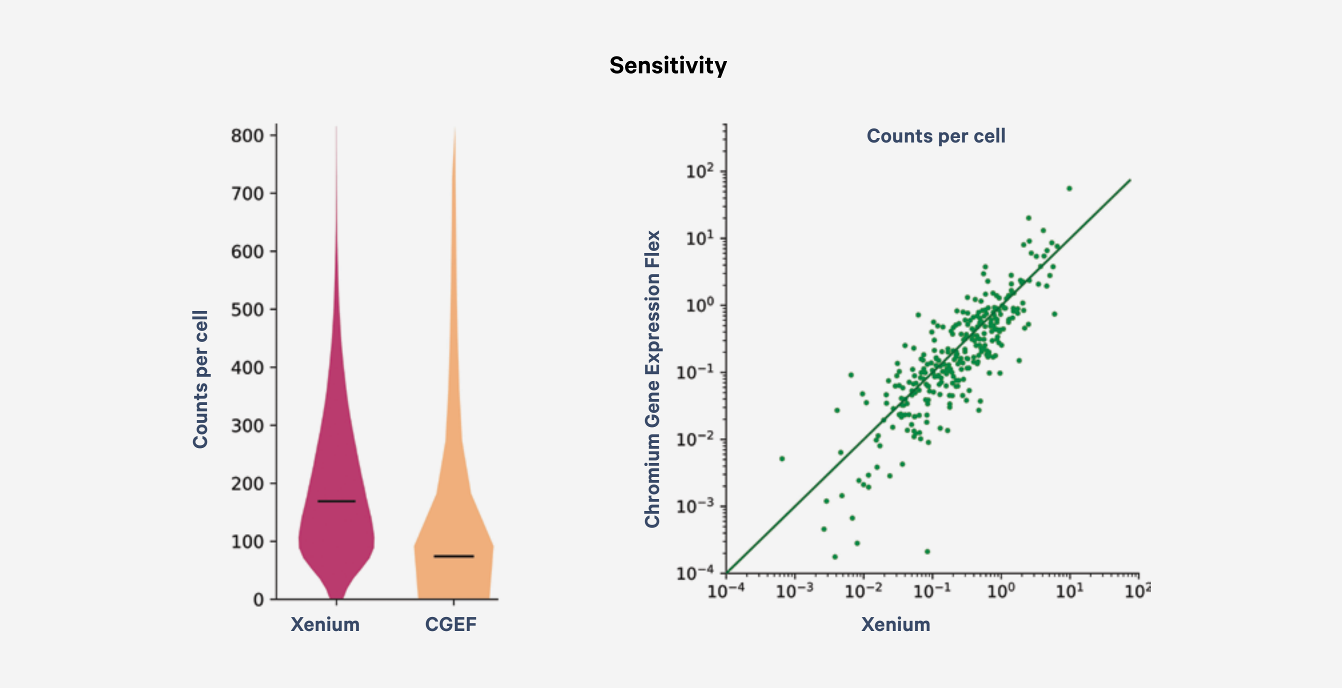


**Figure 8. Benchmarking Xenium specificity, sensitivity, and resolution against Chromium scFFPE-seq.** Chromium Gene Expression Flex data down-selected for only the 313 genes that appear on the Xenium gene panel. Violin and scatter plots showing the total number of transcript counts per cell detected in the Xenium data vs. UMIs per cell in the scFFPE-seq data.

#### Applications

The Xenium platform allows researchers to deepen the understanding of both healthy and diseased tissue through high-plex RNA and multiplexed protein detection at subcellular resolution. The utility of Xenium is demonstrated in numerous publications for:

- Identification of a clinically relevant mechanism that sensitizes diffuse midline glioma to radiation therapy (1)
- Revealing the stepwise molecular and cellular progression of lung adenocarcinoma (2)
- Providing new insights into the spatial niches that underlie pulmonary fibrosis pathology and progression (3)
- Identifying the role of Dkk2 in the Wnt pathway and its potential role in cleft palate (4)
- Characterization of the cellular composition and differentially expressed genes of distinct tumor microenvironments of interest in a ductal carcinoma in situ sample (5)

#### Justification for using the Xenium platform for your research

The Xenium platform offers many advantages, making it the ideal product for analyzing high-plexy spatial gene expression and multiplexed protein information with sub-100 nm resolution. At the core of the platform is the high-throughput Xenium Analyzer instrument, which performs fully automated labeling, imaging, and onboard data analysis.

- **Robust yet flexible core platform**—Xenium is a robust and flexible core platform that can be used for a variety of tissues, sample types, and applications. Additionally, the platform is compatible with many other capabilities, such as simultaneous detection of RNA and protein on the same section, additional third-party analysis features, and additional future capabilities.

- **Unified sample input compatibility**—The Xenium sample prep workflow is compatible with both fresh frozen and FFPE tissue, requires no optimization (even across diverse tissue types), and is non-destructive, which allows the same section to be H&E- or IF-stained post-workflow.

- **High plexity and subcellular resolution**—The Xenium platform currently enables the detection of up to 5,000 transcripts with sub-100 nm localization and high spatial fidelity.

- **Curated gene panels with custom capabilities**—The Xenium platform utilizes pre-designed panels for biologically relevant targets, which are built using a data-driven approach that combines extensive cell atlasing studies with manual curation and invaluable input from research area experts. Xenium panels can be combined and customized with tailored gene sets, including isoforms, exogenous sequences like gRNAs and barcodes, CAR-T transcripts, viruses, and more, delivered ready to use in the Xenium assay.

- **Same-cell spatial multomics**—Xenium Protein enables a first of its kind fully optimized and automated same-cell spatial multimics workflow. Combine ready-to-use Xenium Protein subpanels for immune, tumor, and checkpoint markers with <500-gene panels to correlate cell phenotypes with pathway activity for up to 27 proteins and 480 genes.

- **Highly specific probe chemistry**—The Xenium platform's unique probe chemistry, which leverages a dual hybridization and ligation stringency, enables highly specific binding and the potential to target expressed SNPs and isoforms.

- **Ease of use**—The Xenium platform is a complete and intuitive solution that includes reagents required to prepare samples for analysis as well as instrumentation for image acquisition and software for onboard processing on the Xenium Analyzer. The desktop Xenium Explorer software allows for visualization and interpretation of the data.

- **Best-in-class throughput**—The Xenium Analyzer performs high-plexy imaging at the whole-section scale. Each Xenium slide has a large 12 x 24 mm imageable area, and two slides can be loaded and analyzed simultaneously. Throughput for a specific experiment is determined by the panel plexity and the total tissue area you're analyzing in a single run. For a 480-plex panel on 472 $mm^2$ of tissue, using multimodal cell segmentation Xenium can provide data in less than 3 days. Our 5,000-plex panel using the same tissue area & segmentation can provide data in less than 6 days. You can reduce run time by choosing to analyze subsection(s) of your slide.

- **Software to accelerate data analysis and insight**—10x Genomics provides comprehensive onboard analysis via the Xenium Analyzer as well as state-of-the-art data visualization with the Xenium Explorer desktop software, which is provided free of charge. Onboard analysis output includes primary and secondary analysis—including cell segmentation, transcript and protein assignment to cells, and clustering results. This allows immediate visualization and exploration of gene and protein expression at subcellular and tissue scale in Xenium Explorer without requiring further off-instrument processing. Data is also provided in open standard file formats, allowing scientists the freedom to use other tools of their choice for custom analyses.

- **Broad support resources**—10x Genomics provides comprehensive support resources, ranging from our Technical Support Scientists, Field Application Scientists, Field Service Engineers, and Applied Bioinformatics Scientists, who are trained in the Xenium workflow, instrumentation, and analysis, to freely available videos and documents that guide new users through the Xenium workflow.

- **Certified product quality**—10x Genomics product development and manufacturing processes are ISO 9001:2015 certified.

Note: An internet connection is required for the installation and use of Xenium instruments. 10x Genomics collects certain system logs generated by Xenium instruments, which may be used by 10x Genomics for the purposes of monitoring and improving product performance. Such logs do not include any biological data regarding experimental samples. In addition, when you contact 10x Genomics for troubleshooting or other technical support for your Xenium instrument, 10x Genomics personnel may remotely access the instrument for the purposes of providing such support. Remote access is currently required for most forms of 10x Genomics technical support. For further information, please consult with your 10x Genomics sales representative.

#### References

1. Mangoli A, et al. Ataxia-telangiectasia mutated (Atm) disruption sensitizes spatially-directed H3.3K27M/TP53 diffuse midline gliomas to radiation therapy [Preprint]. *bioRxiv* (2023). doi: 10.1101/2023.10.18.562892
2. Haga, Y, et al. Whole-genome sequencing reveals the molecular implications of the stepwise progression of lung adenocarcinoma. *Nat Commun* 14: 8375 (2023). doi: 10.1038/s41467-023-43732-y
3. Vannan A, et al. Image-based spatial transcriptomics identifies molecular niche dysregulation associated with distal lung remodeling in pulmonary fibrosis [Preprint]. *bioRxiv* (2023). doi: 10.1101/2023.12.15.571954
4. Pina JO, et al. Spatial Multiomics Reveal the Role of Wnt Modulator, Dkk2, in Palatogenesis [Preprint]. *bioRxiv* (2023). doi: 10.1101/2023.05.16.541037
5. Janesick A, et al. High resolution mapping of the breast cancer tumor microenvironment using integrated single cell, spatial, and in situ analysis. *Nat Commun* 14: 8353 (2023). doi: 10.1038/s41467-023-43458-x

#### Resources

Technology overview page
10xgenomics.com/platforms/xenium

**Contact us**
**10xgenomics.com |**

© 2025 10x Genomics, Inc. FOR RESEARCH USE ONLY. NOT FOR USE IN DIAGNOSTIC PROCEDURES.
